# CONSORTiUM-IO: A Phase 1 Study Evaluating VE800 Plus Nivolumab in Treatment of Select Refractory or Metastatic Cancers

**DOI:** 10.64898/2026.11.13.26368051

**Authors:** Carolina Lyon De Ana, Rajita Menon, Shakti K. Bhattarai, Amanda L. Prince, Scott Buresh, Erica Darrah, Jason M. Norman, Rose Szabady, Dmitri Bobilev, Bruce Roberts, Judy S. Wang, Vanni Bucci, Jeremiah Faith, Jeffrey L. Silber, Kenya Honda, Bernat Olle, Diwakar Davar

## Abstract

Microbiome modulation can augment responses to immunotherapy in advanced cancer. Early studies of anti-PD-1 therapies in combination with fecal microbiota transplantation (FMT) reported efficacy in immunotherapy-naïve and -refractory settings. However, FMTs are variable in composition, difficult to scale, and risk the transfer of emerging pathogens. VE800 is a consortium of 11 strains of commensal bacteria derived from healthy donors and grown from clonal cell banks. VE800 increased anti-PD-1 efficacy in preclinical models of cancer. CONSORTiUM-IO was a first-in-human, open-label phase 1 study that evaluated safety and efficacy of VE800 with nivolumab in advanced PD-1-refractory melanoma, PD-1-naive gastric/gastroesophageal junction (G/GEJ) adenocarcinoma and PD-1-naive microsatellite-stable colorectal cancer (MSS CRC). VE800 plus nivolumab was well tolerated but offered no improvement over historic response rates. Exploratory analyses in patients with melanoma revealed that VE800 strain engraftment was associated with greater progression-free survival (PFS) and increased immune activation peripherally.

**STATEMENT OF SIGNIFICANCE:** The first-in-human study of the safety and efficacy of a defined bacterial consortium in combination with anti-PD-1 treatment in various solid tumors demonstrated the safety of the combination, albeit with low clinical efficacy. The results provide important knowledge and considerations about microbiome modulation in cancer, to help design future trials.

## INTRODUCTION

The use of immune checkpoint inhibitors (ICIs), including programmed death-1 (PD-1) and programmed death-ligand-1 (PD-L1) inhibitors, has fundamentally changed treatment and outcomes for many patients with cancer. Inhibiting PD-L1/PD-1 signaling relieves tumor-mediated immune suppression, reverses T cell exhaustion, and results in anti-tumor immune responses and improved survival in multiple solid tumors^1^. Unfortunately, not all patients respond, and many responses are not durable.

Multiple predictive biomarkers of ICI benefit have been described, including CD8+ tumor-infiltrating lymphocyte (TIL)-infiltrate^2^, PD-L1 expression^3^, and tumor mutation burden (TMB)^4^. More recently, multiple studies have examined the gut microbiota composition of ICI-treated patients with cutaneous melanoma^5–8^, non-small cell lung cancer (NSCLC)^9,10^, and renal cell carcinoma (RCC)^9,11^, to infer putative relationships between the presence of key bacterial species and ICI response. These studies identified specific members of each of the major bacterial phyla colonizing humans, namely, *Firmicutes*, *Actinobacteria*, *Proteobacteria*, *Bacteroidetes*, and *Verrucomicrobia*, in patients with durable ICI benefit. Preclinical studies have confirmed that supplementation with favorable commensal bacteria can restore the efficacy of PD-1 blockade and promote interferon gamma (IFN-γ)-secreting T cells and other antitumor responses^12^.

In humans, non-randomized and randomized studies of fecal microbiota transplantation (FMT) in combination with anti-PD-1 treatment have demonstrated preliminary efficacy in patients with ICI-naïve, non-small cell lung cancer, and ICI-refractory melanoma and metastatic renal cell carcinoma^13–18^. In these studies, FMT appeared to augment anti-PD-1 efficacy and reverse resistance. However, donor-derived fecal microbiota products, such as FMT, are highly variable in their composition, have low scalability, and carry a potential risk of pathogen transfer. Live biotherapeutic products (LBPs) composed of defined bacterial consortia are an attractive alternative to FMT, having demonstrated preliminary efficacy in preventing recurrent *Clostridioides difficile* infection (rCDI) in high-risk individuals^19,20^.

VE800 is an LBP consisting of 11 commensal bacterial strains, with evidence of efficacy in mouse models of melanoma and adenocarcinoma (**Supplementary Table S1**)^12^. Here, we examined the ability of VE800 to enhance the efficacy of an anti-PD-1 agent, nivolumab, in a first-in-human, open-label, multicenter, phase 1 study of patients with select advanced solid tumors including advanced melanoma refractory to anti-PD-1, PD-1-naïve microsatellite stable colorectal cancer (MSS-CRC), and PD-1-naïve gastric/gastroesophageal carcinoma (G/GEJ), after a pretreatment with vancomycin, to create a microbial niche for strain colonization.

## RESULTS

### Safety and Efficacy of VE800 and Nivolumab in PD-1-Resistant Melanoma, and PD-1-Naive G/GEJ or MSS-CRC

Seventy patients were screened, and 56 patients were enrolled between January 2020 and December 2020 across the three cohorts: melanoma (n=21), G/GEJ (n=21), and MSS-CRC (n=14) (**Fig. 1A**). Of these, two patients (1 melanoma, 1 G/GEJ) were excluded due to noncompliance with vancomycin pretreatment and not meeting the eligibility criteria prior to starting vancomycin pretreatment, respectively. Fifty-five patients who received at least 1 dose of vancomycin were evaluable for safety, and 54 patients who received vancomycin pretreatment plus at least 1 dose of VE800 and nivolumab were evaluable for efficacy (**Supplementary Fig. S1**). Baseline characteristics and demographics of patients enrolled are listed in **Supplementary Table S2**. The median age was 62 years, with a male predominance (56.4%) and Caucasian ethnicity (92.7%). The median number of prior systemic therapy was 2 (range, 1 to 3) in the melanoma, 2 (range 1 to 4) G/GEJ, and 3 (range 1 to 4) MSS-CRC cohorts.

**Figure 1:**
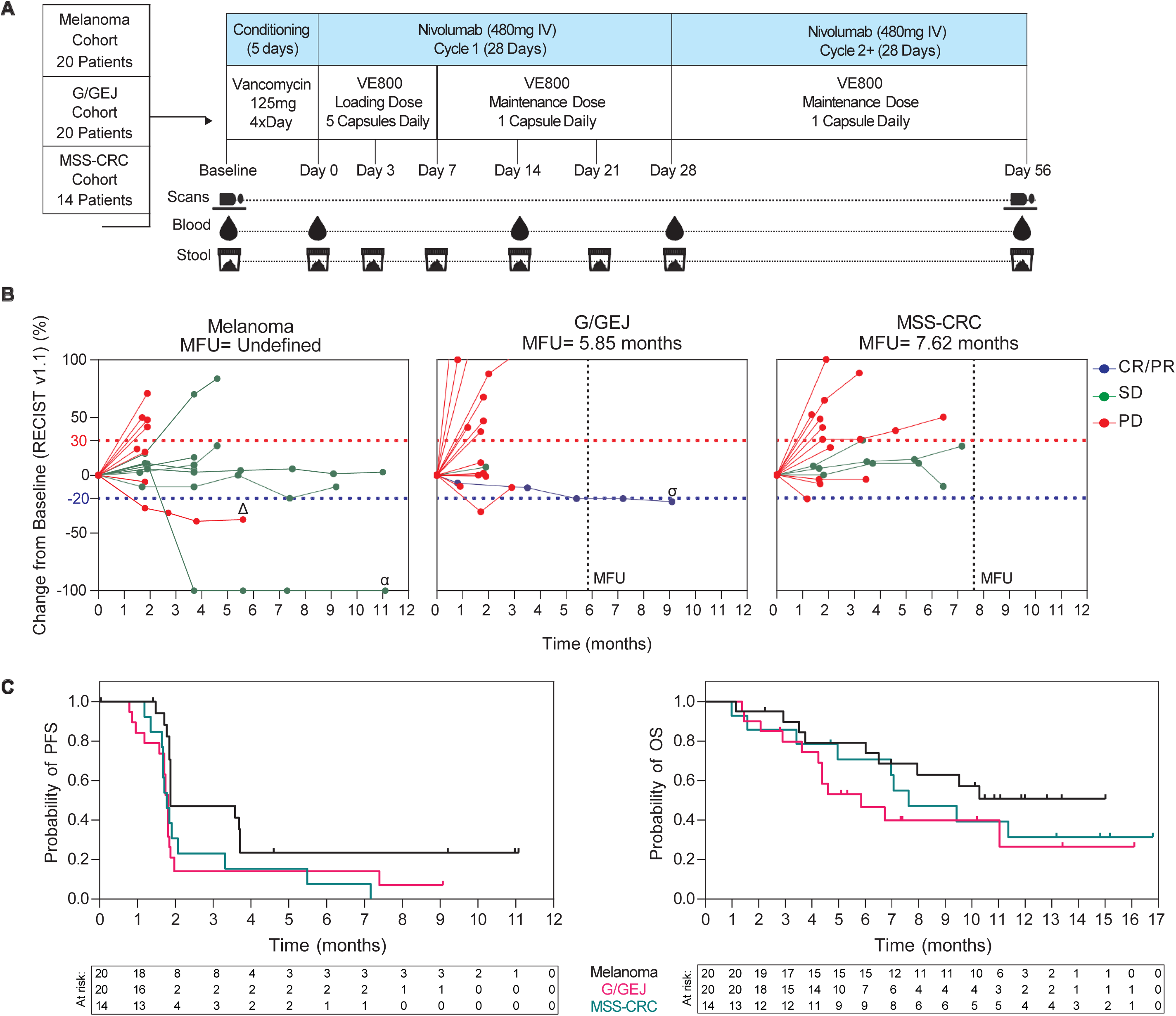
Study Design and Clinical Efficacy of VE800 with Anti-PD-1 in All Cohorts. A) CONSORTiUM-IO Stage 1 dosed 20 patients each in the melanoma and G/GEJ cohorts, and 14 patients in the CRC-MSS cohort. All patients received oral vancomycin pretreatment (125mg 4 times daily for 5 days), followed by a VE800 Loading Dose of 5 capsules (5.5x10^8^ CFU per capsule) daily for 7 days, for a total dose of 1.9x10^10^ CFU. Patients received a VE800 maintenance dose of 1 capsule daily from Day 7 until the end of study. Nivolumab was administered at 480mg IV every 28 days. Tumor scans, blood, and stool were collected at the indicated timepoints throughout the study. B) Percent tumor change from baseline over time, per cohort from the Efficacy Population using RECISTv1.1 criteria. Vertical dotted lines indicate the median follow-up (MFU) for each cohort, also defined under cohort title. Horizontal dotted lines indicate 30% increase from baseline (red) and 20% decrease from baseline (blue). Δ-patient exhibited a mixed response, target lesion was reduced but non-target lesion showed progression. α-target lesion was resected at cycle 3 and radiologic assessment indicated SD. σ-patient in gastric cohort with highest level of PD-L1 expression in tumor biopsy. C) Probability of PFS (Left) and Probability of OS (Right) in months. Tables indicate censored patients at each timepoint. Melanoma subjects are signified in black, G/GEJ subjects are signified in pink, and CRC-MSS are signified in green.

Among the 55 safety-evaluable patients, treatment-emergent adverse events (TEAEs) were reported in most (47 [87%]), with the highest percentage in the G/GEJ cohort. Treatment-related adverse events (TRAEs) occurred in 24 (44.4%) patients and were mostly related to nivolumab treatment (21 [38.9%]) (**Supplementary Table S3**). VE800 was generally well tolerated, with VE800-related TRAEs occurring in 24 patients (44.4%) and only 1 instance (1.9%) of Grade 3+ VE800-related TEAE (diarrhea) in a G/GEJ subject. No Grade 5 events were reported (**Supplementary Table S3**). Nivolumab-related TRAEs were expected and low-grade. TEAE-related dose interruption occurred in 14.8% of patients, the majority of which were related to nivolumab (9.3%) and summarized in **Supplementary Table S3**.

Investigator-assessed objective response rate (ORR) in the efficacy-evaluable patients (n=54) was low, at 1.9% (1/54), with 1 partial response (PR) in a G/GEJ patient that lasted 5.6 months (**Fig 1B, Supplementary Table S4**). The disease control rate (DCR) was 27.8% (15/54) overall, and 45% (9/20), 10% (2/20), and 14.3% (2/14) in the melanoma, G/GEJ and MSS-CRC cohorts, respectively (**Fig 1B, Supplementary Table S4**). Across the entire study population, median progression-free survival (PFS) was 1.8 months (range, 0.8-11 months) and median overall survival (OS) was 9.4 months (range, 1.0-16.8 months), highest in the melanoma cohort (**Fig 1C, Supplementary Table S4**). Landmark 6-month PFS and OS statistics are shown in **Supplementary Table S4**). The responding G/GEJ patient had high PD-L1 expression at baseline and given the established benefit of anti-PD-1 therapy in G/GEJ, the observed efficacy was likely related to nivolumab^21^. Given the low clinical activity observed at the end of Stage 1, the study did not meet the prespecified criteria to support continuing into Stage 2. Patients were permitted to continue treatment, including VE800, at the investigator’s discretion but no further patients were enrolled. For the purposes of secondary and exploratory analyses, we subsequently stratified patients based on their best response at week 8: stable disease (SD, PR) vs. progressive disease (no SD or PR) (in **Supplementary Table S4, S5**).

### VE800 Colonization and Alterations in the Gut Microbiome

Stool samples were collected longitudinally throughout the study and metagenomic sequencing was performed to quantify prevalence and relative abundance of VE800 strains (**Fig 1A, Supplementary Table S6**). By Day 14, the number of strains detected and total relative abundance were significantly greater than baseline across all cohorts (*p* < 0.0005 for detection, *p* < 0.005 for abundance), with sustained colonization up to 86 days after the start of VE800 administration in the melanoma and MSS-CRC cohorts (**Fig 2A, Supplementary Fig. S2, Supplementary Table S7**). Individual strains were detected at a median relative abundance of 0.1 to 1% over time. Eight of 11 strains were significantly increased by Day 14 (adjusted *p* < 0.05); however, strains VE800-05 and -06 did not colonize significantly above baseline in any cohort, and strain VE800-08 was only detectable in patients with melanoma, at low abundances (**Supplementary Fig. S2, Supplementary Table S7**). Altogether, colonization of most VE800 consortium strains was observed across the different cohorts.

**Figure 2:**
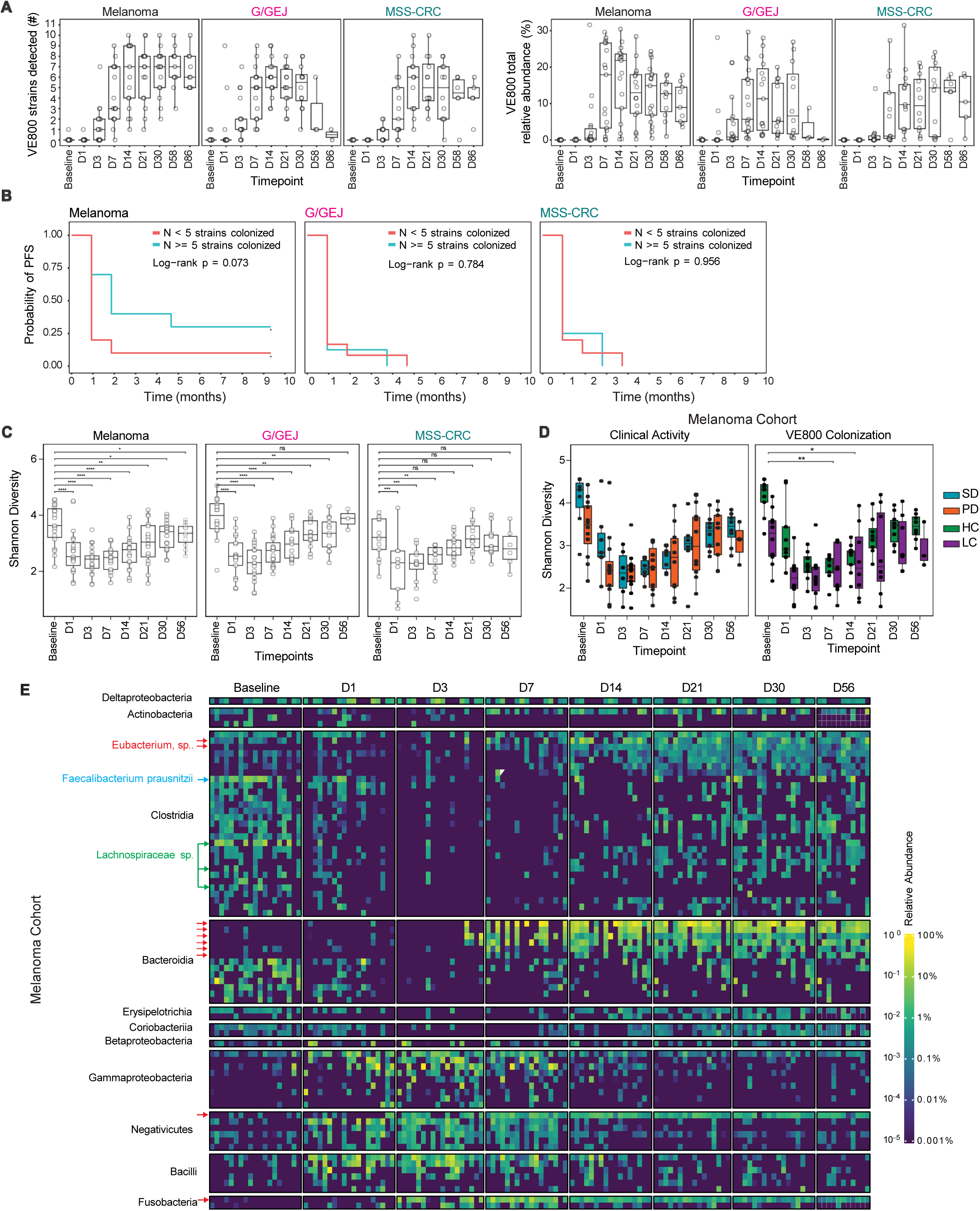
Colonization of VE800 and Microbiome Changes During Study. A) Number of VE800 strains detected (Right) and total relative abundance of the VE800 strains (Left) as a proportion of a subject’s total gut microbiota at the corresponding timepoint. The box-and-whisker plots depict the median, interquartile range, and reasonable extreme values at 1.5 × the inter-quartile range in the dataset. B) PFS in subjects dosed with VE800 as a function of strain colonization during the first month after the start of dosing in melanoma, G/GEJ, CRC-MSS. Patients were divided into high and low colonization with VE800 based on cox models described in Table S7. C) Box plot depicting Shannon diversity across multiple time points for all cohorts. Asterisks (*) indicate statistically significant differences between time points and baseline, as determined by LME (Shannon ∼ Time + (1|SubjectID)). D) Box plot depicting Shannon diversity by clinical outcome classification (left) and colonization of VE800 (right). Asterisks (*) indicate statistically significant differences between time points and baseline, as determined by LME (Shannon ∼ Time × Response + (1|SubjectID)). The number of asterisks corresponds to the level of significance: * p < 0.05, ** p < 0.01, *** p < 0.001. E) Heatmap showing the relative abundance of differentially abundant microbiome species (FDR < 0.05) in the melanoma cohort, as determined by a linear mixed-effects model (RA ∼ Time + (1|SubjectID)), comparing each time point to baseline. Red arrows indicate VE800 species relatives.

To assess the relationship between strain colonization and PFS, we first determined colonization based on the average number of VE800 strains detected in each patient during the first month after the start of dosing and used cutpoint analyses to test impact of colonization categories upon outcomes across the different cohorts. No significant relationship between colonization and PFS was observed in the G/GEJ and MSS-CRC cohorts at any colonization threshold (**Supplementary Table S8**). In the melanoma cohort, we generally observed higher VE800 strain colonization and higher relative abundance of VE800, compared to G/GEJ and MSS-CRC (**Supplementary Fig. S2, Supplementary Table S7**). In this cohort, higher colonization was associated with improved PFS in 3 models (≥5 strains, p=0.07; ≥6 strains, p = 0.048; ≥7 strains, p = 0.1), with all other thresholds non-significant. However, to maintain balanced sample numbers across groups, the cutoff of ≥5 strains was used to classify patients in the melanoma cohort as high (HC; ≥5 strains) or low (LC; <5 strains) colonizers (**Supplementary Table S9**). The probability of PFS was non-significantly increased in melanoma HC compared with LC (p = 0.073); the median PFS (the earliest timepoint at which the progression-free probability drops to or below 0.5) for HC and LC was 8 weeks and 4 weeks, respectively (**Fig. 2B**). Melanoma HC had higher abundance of strains VE800-03 and -04, two out of the four VE800 effector strains that were shown to promote IFN-γ induction by CD8+ T cells in preclinical models (**Supplementary Fig. S3, Supplementary Table S10**)^12^.

Vancomycin pretreatment resulted in reduced Shannon diversity, in line with previous observations (**Fig. 2C-E, Supplementary Fig. S4A-B, Supplementary Table S11, S12**)^20,22^. Species diversity recovered to baseline following administration of vancomycin and VE800 in the MSS-CRC and G-GEJ cohorts, whereas diversity in the melanoma cohort recovered over time but remained slightly lower than baseline levels by day 56 (Δ=0.41) (**Fig. 2C, Supplementary Table S12**). This reduced diversity was similar by clinical response (SD vs. PD) and colonization status (HC vs. LC) (**Fig. 2D, Supplementary Table S13**). In all cohorts, the endogenous microbiome at baseline was dominated by species belonging to the Clostridia and Bacteroidia classes (**Fig. 2E, Supplementary Fig. S4 A,B, Supplementary Table S13**). Vancomycin pretreatment resulted in increased relative abundance of Bacilli, Negativicutes, and Gammaproteobacteria species, and depletion of Bacteroidia and Clostridia species. The depleted species included members of Clostridia associated with responses to anti-PD-1, including *Faecalibacterium prausnitzii* and various *Lachnospiraceae* (**Fig. 2E-F, Supplementary Fig. S4A,B, Supplementary Table S13**)^6,9,23^, and members of Bacteroidia associated with non-response to anti-PD-1 (**Fig. 2E, Supplementary Fig. S4A-B, Supplementary Table S13**)^13,14,24^. VE800 enhanced colonization with members of the same classes, including *Eubacterium limosum* and other species of Bacteroidia, associated with non-response and response to anti-PD-1, respectively (**Fig. 2E, Supplementary Fig. S4A-B**)^6,9,13,14,24^. After the completion of the VE800 trial, the TOPOSCORE algorithm became an established tool to understand these collective microbiome changes in the context of immune checkpoint blockade outcomes^25^. In support of the TOPOSCORE as a predictive measure, we found baseline TOPOSCORE was higher in melanoma patients who achieved stable disease than in those who progressed (p=0.002). Consistent with the diversity outcomes, the microbiome community assessed by the TOPOSCORE shifted towards a less checkpoint-favorable composition after vancomycin preconditioning in all three cohorts (**Supplementary Fig. S5**) with a full recovery to at least the baseline level in MSS-CRC and G-GEJ but not in melanoma. These findings suggest that most VE800 strains colonize the host to some degree and that vancomycin pretreatment reduced microbiome diversity and worsened the TOPOSCORE with only partial recovery to baseline in the melanoma cohort.

### Endogenous Baseline Factors Impacting VE800 Colonization in Melanoma Cohort

To study impact of baseline host factors upon VE800 colonization, we used ordinal regression models to evaluate association between strain-level VE800 colonization and baseline fecal microbiome and serum metabolites, focusing on the melanoma cohort, which had the greatest degree of colonization. All strains were included in the analysis except VE800-06 (*Paraprevotella xylaniphila*), which had consistently poor colonization across patients (**Supplementary Table S1, S7**).

Endogenous taxa from Proteobacteria/Pseudomonadota, Verrucomicrobiota, and Coriobacteriia were negatively associated with colonization of several Bacteroidia/Bacteroidota VE800 strains, whereas Clostridia, Actinomycetota, and additional Bacteroidia species showed positive associations (**Fig. 3A, Supplementary Fig. S6, Supplementary Table S14**). The presence of *Oscillibacter* spp., and species from the Bifidobacteriales, Bacteroidiales and Erysipleotrichales orders were positively linked with colonization of 3 effector VE800 strains (-02, -03, -04) that induce IFN-γ production by CD8+ T cells in preclinical models^12^. Most serum short-chain fatty acids (SCFAs) and bile acids were negatively associated with colonization, with taurodeoxycholic acid being the only factor favorably associated with colonization of VE800-02 (**Fig. 3B, Supplementary Table S14**)^26^.

**Figure 3:**
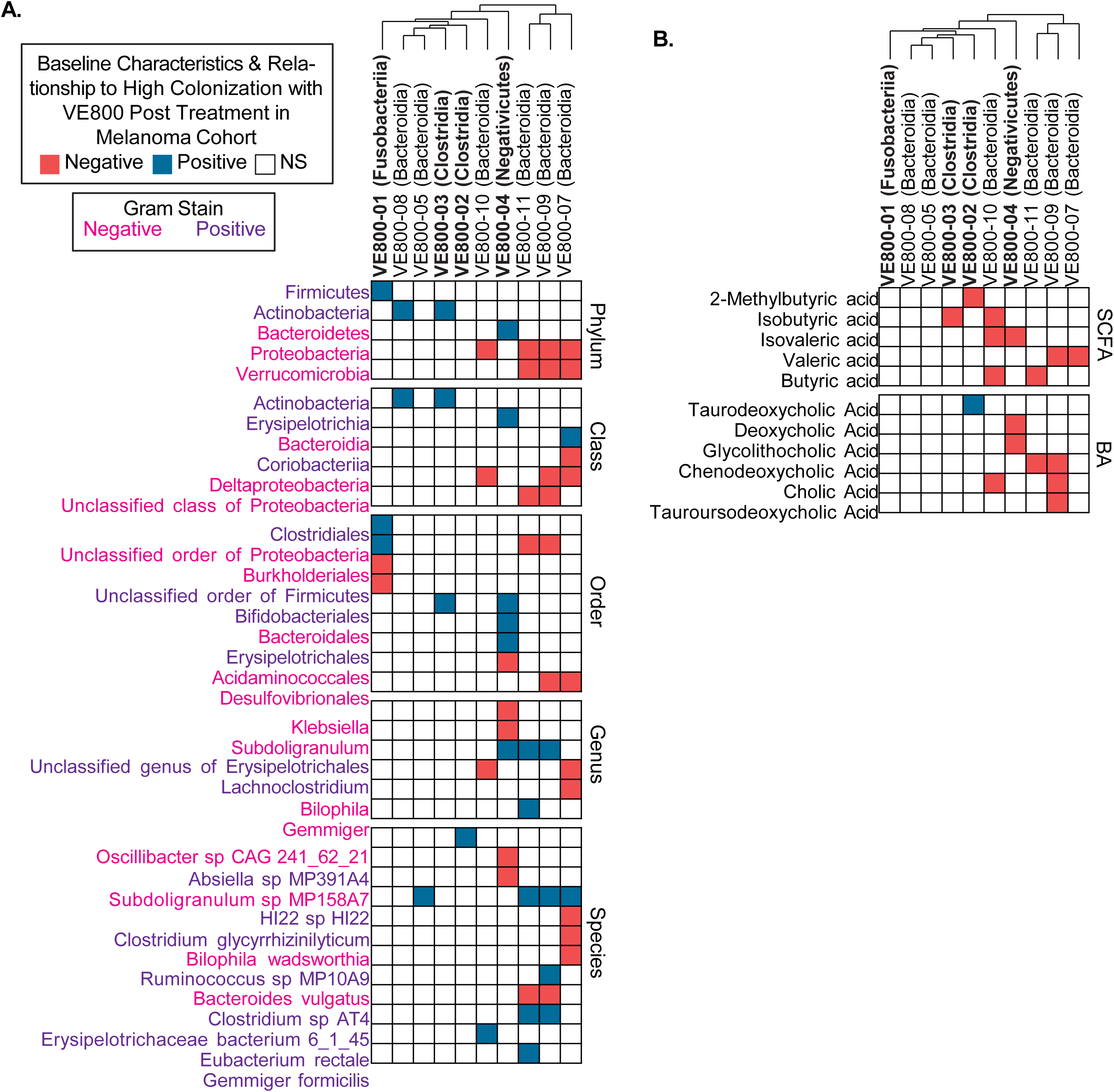
Baseline Characteristics and Relationship to High Colonization with VE800 Strains in Melanoma Cohort. Heatmap illustrating the association of multimodal biomarkers as predictors of strain colonization, as determined by an ordinal regression model (see Methods). A) Baseline endogenous microbial species from stool metagenomic sequencing. Microbial species color coded as Gram positive (purple) and Gram negative (pink). B) Baseline SCFA (Top) and BAs (Bottom) from serum analysis. Coloring of boxes depict a positive (blue) or negative (red) association between baseline biomarker and colonization of each strain. VE800 effector strains (IFN-γ producers in preclinical models) marked in bold.

### Metabolomic and Immune Changes in the Melanoma Cohort

To further evaluate the impact of VE800 strain colonization upon anti-tumor immunity, we evaluated serum metabolomics and circulating immunological parameters in the melanoma cohort by colonization status (HC vs. LC) and clinical activity (SD vs. PD). Response to FMT plus anti-PD-1 in refractory melanoma was linked to increased primary bile acids (PBAs)^13^, but longitudinal analyses revealed no significant changes in SCFAs with VE800 and nivolumab in the melanoma cohort by colonization status (HC vs. LC) and clinical activity (**Supplementary Fig. S7, Supplementary Table S15**). However, relatively little transformation of PBAs into secondary bile acids (SBAs) was observed, irrespective of clinical response. Low colonization with VE800 strains resulted in increases in both PBAs and conjugated SBAs (**Supplementary Fig. S8, Supplementary Table S16**). VE800 treatment did not result in any significant changes in serum metabolites.

Cytokine profiling revealed that baseline FLT-3L was elevated in patients who later achieved SD, whereas IL-17 was elevated in HC patients. By Day 14, patients with SD and HC showed increases in both pro-inflammatory/tumoricidal cytokines (e.g., CCL5, FLT-3L, IL-12p40, CXCL9, IL-21)^27^ and cytokines with pro-tumor or immunosuppressive associations (e.g., CCL22, Eotaxin)^27,28^. By Day 30, patients with SD exhibited increases in IL-21 and TRAIL, whereas patients with PD showed elevations in IL-17 family cytokines; those with HC demonstrated increases in both inflammatory cytokines (FLT-3L, TRAIL, IL-9, IL-21) and immunosuppressive cytokines (e.g., CCL22)^27^ (**Fig. 4A, Supplementary Tables S17**).

**Figure 4:**
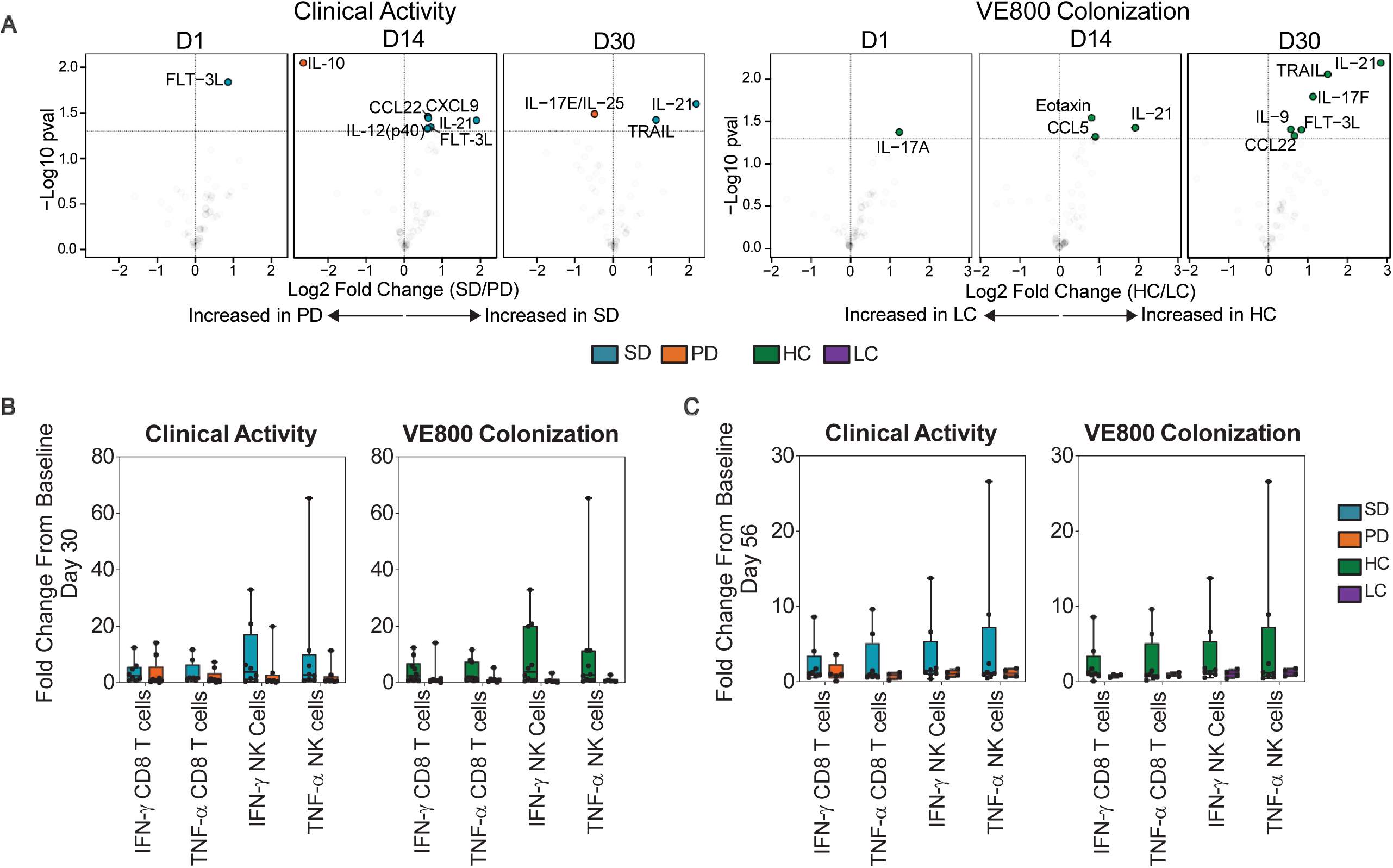
Systemic Immune Changes in Melanoma Cohort During Study. A) Volcano plots of serum cytokines significantly changed from baseline over time in patients stratified by Clinical Activity (Left) and VE800 Strain Colonization (Right). LME analysis, abundance∼timexResponse(SD/PD or HC/LC)+(1/subjectID), p<0.2. Fold change from baseline in IFN-γ CD8 T cells, TNF-α CD8 T cells, IFN-γ NK cells, and TNF-α NK cells in B) Day 30 and C) Day 56. Box plots with whiskers of minimum to maximum fold change showing all subjects.

To evaluate effects of VE800 and nivolumab upon systemic immunity, we used spectral flow cytometry (**Supplementary Fig. S9-13**). We observed that VE800 plus nivolumab resulted in increased IFN-γ-producing CD8 T cells in patients with HC by day 30 (**Fig. 4B**). Additionally, HC melanoma patients also exhibited increased pro-inflammatory TNF-α+ CD8 T cells, cytotoxic CD107a+ CD8 T cells, IFN-γ, and TNF-α+ NK cells by day 30, with responses attenuating by Day 56 (**Fig. 4B-C**). Regulatory T cells (Tregs) remained largely unchanged (**Supplementary Fig. S13A**). Immunosuppressive myeloid-derived suppressor cells (MDSCs) were greater at baseline in the melanoma cohort relative to healthy controls, although MDSCs decreased with treatment over time in patients with SD and those with HC (**Supplementary Fig. S13B**). Subsets of HLA-DR^hi^ MDSCs, previously associated with beneficial outcomes following FMT and anti-PD-1 in refractory melanoma^13^, were unchanged with treatment here (**Supplementary Fig. S13D**). Overall, VE800 plus nivolumab induced mixed pro-inflammatory and immunosuppressive immune signatures, without durable metabolic or systemic immune reprogramming.

## DISCUSSION

We have reported the results of the first-in-human trial of a rationally-designed LBP in combination with an anti-PD-1 therapy, nivolumab, in patients with three types of solid tumors: ICI-resistant advanced melanoma, ICI-naïve MSS-CRC and ICI-naïve G/GEJ. VE800 plus nivolumab was well tolerated, with most TRAEs attributed to anti-PD-1, and only 1 serious TEAE attributable to VE800, consistent with the tolerability profile of LBPs in other non-oncologic indications^19,20,29^. Intriguingly, some patients with ICI-resistant melanoma may have benefited from VE800 plus nivolumab, as the landmark 6-month PFS rate of 23.5% exceeded that observed with anti-PD1 monotherapy^30^. The clinical activity of VE800 plus nivolumab was low; the observed efficacy did not meet prespecified criteria for further development, and so the study was truncated.

Consistent with prior LBP studies, most VE800 strains engrafted efficiently across all 3 cohorts after vancomycin pretreatment, indicating that the lack of clinical benefit was not due to poor colonization^19,20,29^. In the melanoma cohort, HC correlated strongly with baseline enrichment in IFN-γ-promoting species (e.g., VE800-03 *Ruthenibacterium lactatiformans*, ordinal regression coefficients >0.1, p < 0.05) and inversely with most SCFAs, PBAs, and SBAs. However, the importance of strain-level engraftment in mediating efficacy of microbiome modulation in cancer is unclear.

In the absence of PFS in the G/GEJ and MSS-CRC cohorts, metabolomic and immune analyses were restricted to the melanoma cohort. Unlike prior FMT plus anti-PD-1 studies that reported increases in SCFAs and SBAs post-FMT^13^, no significant serum metabolite changes were detected after VE800 administration. VE800 plus nivolumab elicited transient elevations in inflammatory cytokines and chemokines (e.g., FLT-3L, IL-12p40, IL-9, CXCL9) by day 30, and modest increases in IFN-γ–producing CD8⁺ T cells in patients with HC or SD, patterns previously linked to ICI responsiveness^31^. However, immunosuppressive mediators such as CCL22 rose, and MDSCs and other suppressive populations^31^ remained unchanged. Given the non-randomized nature of the study, we are unable to state whether the elevations in inflammatory cytokines and chemokines are related to the combination of VE800 and anti-PD-1 or the use of anti-PD-1.

There were several limitations in this study. The small sample sizes of each cohort and lack of an anti-PD-1 monotherapy control limited our ability to draw conclusions. Patients with G/GEJ were enrolled prior to chemoimmunotherapy being an established modality, and hence the efficacy observed in the G/GEJ patient could be related to anti-PD-1 use. Although vancomycin pretreatment likely facilitated robust strain engraftment, antibiotics have been associated with impaired response to checkpoint-inhibitors^22,32^. In this study, the vancomycin pretreatment at least temporarily altered the microbiota to a less favorable state for check-point response as estimated by the TOPOSCORE^25^. However, without an antibiotic placebo, we cannot directly assess the vancomycin impact on outcomes. Moreover, the early discontinuation of progressing patients precluded collecting correlative samples at later timepoints and limited late analyses of pharmacodynamic and pharmacokinetic parameters. In addition, the lack of dietary data and enrollment of a predominantly Caucasian population limited the generalizability of these findings.

Collectively, although VE800 plus nivolumab was well tolerated, the efficacy in melanoma, G/GEJ, and MSS-CRC was low. Although this combination did not improve response to anti-PD-1 in these indications, our results suggest the ability of an LBP consisting of a defined bacterial consortium to elicit a relevant, albeit temporary, systemic response in some patients. It remains unclear whether the augmentation of beneficial bacteria, or depletion of deleterious bacteria, or both could lead to enhancing immunotherapy responses^17,18^. The strong context dependence of the gut microbiome across disease settings, ICI partners, and host factors, substantially complicates the rational development of LBPs. In conclusion, this study provides valuable insights into baseline microbiome and systemic profiles that may influence colonization with specific strains. More clinical studies are necessary to better understand the role of microbiome modulation in enhancing immunotherapy responses.

## METHODS

### Study Design

CONSORTiUM-IO (NCT04208958) was a first-in-human, open-label, phase 1 study of VE800 plus the anti-PD-1 agent, nivolumab, in selected tumors: advanced cutaneous melanoma with relapse following at least one line of anti-PD-1 therapy; recurrent locally advanced or metastatic G/GEJ adenocarcinoma with progression on or after treatment with at least one line of therapy; and advanced or metastatic MSS-CRC with progression after treatment with at least one line of therapy.

The study followed a Simon’s 2-stage design, to provide initial assessment of clinical activity in Stage 1 with potential advancement to Stage 2. Stage 1 aimed to enroll 21 patients in the melanoma and G/GEJ cohorts, and 14 in the CRC-MSS cohort. The prespecified ORR targets were 9.5% (2/21, melanoma), 9.5% (2/21, G/GEJ), and 7.1% (1/14, MSS-CRC), respectively. The study did not meet any of the prespecified ORR criteria for advancement to Stage 2.

The primary objective was to evaluate safety, tolerability, and clinical activity of VE800 in combination with nivolumab; secondary endpoints included measures of clinical benefit and metagenomic and translational results correlating strain engraftment with response.

This multi-center study was conducted in accordance with Good Clinical Practice guidelines. The protocol and informed consent documents were approved by the Institutional Review Board or ethics committee for each participating site, and all patients provided written informed consent before any study procedure. Approving boards and protocol numbers were: University of Utah IRB (IRB_00128490); University of Pittsburgh Medical Center IRB (MOD00655221, Pro00039903); Advarra Center for IRB Intelligence Platform (MOD00655221, MOD0146106, 00000971); WCG Institutional Review Board (study numbers 1282434, 1273387, 1275793, 1283587); New York University School of Medicine (i19-00845); Rhode Island Hospital & Miriam Hospital (204120); University of California Los Angeles (IRB#20000133-AM00006); Cedars-Sinai Samuel Oschin Comprehensive Cancer Institute (CR2019-1002-0); University of Chicago Medical Center (IRB19-1676); Sutter Health IRB (1549521-7); and Weill Cornell Medicine (20-02021418).

### Patient Eligibility

Eligible patients had to be at least 18 years of age, have advanced cancer with select histologies as noted above, had received no more than 3 lines of prior systemic therapy for advanced disease, and had to have adequate organ function. For enrollment in the melanoma cohort, patients must have had confirmed progressive disease (on at least two consecutive scans at least 4 weeks apart) on or within 3 months of discontinuation of prior PD-1/PD-L1 antibody as a single agent or combination with a standard or investigational therapy. Patients who progressed on or within 3 months of adjuvant therapy with ICI were eligible, and the adjuvant therapy counted as one prior line of therapy. An anti-PD-1/PD-L1 therapy must have been the most recent therapy received. For enrollment in the G/GEJ cohort, patients had to have recurrent locally advanced or metastatic gastric or GEJ adenocarcinoma, with disease progression on or after at least one line of therapy including fluoropyrimidine- and platinum-containing chemotherapy and, if appropriate, human epidermal growth factor receptor 2 (HER2)/neu-targeted. The MSS-CRC cohort enrolled patients with histologically confirmed advanced/metastatic MSS-CRC based on either an analysis of tissue from a prior biopsy or based on tissue from a new biopsy (patient’s microsatellite/mismatch repair status should have been known) who had progressed following at least one line of standard treatment for advanced/metastatic disease, which must have included fluoropyrimidine, oxaliplatin, or irinotecan, unless contraindicated. Patients who relapsed within 6 months of adjuvant chemotherapy composed of oxaliplatin and a fluoropyrimidine had their adjuvant therapy count as one prior regimen.

### Intervention

VE800 is a defined LBP composed of 11 commensal bacterial strains (**Supplementary Table S1**) that were isolated from a healthy human donor and manufactured from clonal cell banks^12^. The strains were selected based on their ability to promote IFN-γ-secreting CD8 T cells in mice, and to enhance the efficacy of anti-PD1 treatment in mouse models of melanoma and adenocarcinoma^12^. The dosing regimen of VE800 was selected based on previous LBP studies in healthy volunteers^20,29^.

Eligible patients received oral vancomycin pretreatment (125 mg, four times daily for 5 days), followed by a loading dose of VE800 (5 capsules of 5×10^8^ colony-forming units [CFU] per capsule, once daily) for 7 days, followed by a maintenance dose (1 capsule of 5×10^8^ CFU, once daily) in combination with nivolumab (480 mg intravenously [IV] every four weeks) for up to two years or discontinuation for toxicity, progression, or non-compliance (**Fig. 1A**). Anti-tumor response was evaluated with imaging at baseline, repeated every 8 weeks thereafter, and assessed by the investigator using RECIST v1.1. Peripheral blood and stool were collected throughout the study (**Fig. 1A**).

### Patient Enrollment

In total, 70 patients were screened, with 14 screen failures, resulting in 56 patients enrolled across three cohorts: melanoma (n=21), G/GEJ (n=21), and MSS-CRC (n=14) (**Fig. 1A**). A total of 54 patients were dosed with VE800 plus anti-PD1; two enrolled patients did not receive study drug therapy (**Supplementary Fig. S1**). Patients were enrolled between January 2020 and December 2020 and followed up to September 2022.

### Trial Procedures

The trial consisted of a screening period of 19 days that included baseline tumor assessment, tumor biopsy, and additional laboratory testing to assess eligibility. Eligible patients received the vancomycin pretreatment to create a microbial niche for VE800 strain colonization, followed by orally administered VE800 at 5 capsules per day (5.5 x 10^8^ CFU per capsule; daily dose, 2.79 x 10^9^ CFU per day) for the 7 first days (total loading dose, 1.93 x 10^10^ CFU), followed by 1 capsule per day (5.5 x 10^8^ CFU) continuously. Nivolumab (480mg IV) was administered beginning at cycle 1 day 1 (C1/D1), on the first day of VE800 administration. Intervention cycles of nivolumab occurred every 4 weeks. All patients continued study treatment until progression, unacceptable toxicity, withdrawal of consent, study discontinuation criteria met, completion of 2 years of treatment, or after 5 cycles of nivolumab were completed. Patients benefiting from treatment, as judged by the investigator, were allowed to continue treatment for up to 2 years.

Although we attempted to collect tumor tissue for biomarker analysis, insufficient tumor samples were collected to permit analyses. Stool samples were collected for VE800 strain detection, colonization duration, and other microbiome components such as overall diversity. Blood samples for PBMC isolation and serum cytokine analysis were collected throughout the study and at time of progression for exploratory analysis.

### Endpoints and Assessments

All patients who received at least 1 dose of VE800 plus nivolumab and had a valid baseline and post-baseline tumor assessment (death or clinical progression without imaging were considered post-baseline assessments) were included in the Efficacy Population for analysis. Tumor assessment was obtained at baseline and every 8 weeks thereafter, and confirmation of change was at least 4 weeks between previous tumor assessment with imaging scans. After 1 year, patients remaining on treatment were to have imaging studies repeated every 12 weeks. Primary endpoint evaluation was done following RECIST v1.1^33^.

ORR was used as the primary efficacy endpoint, and was defined as the proportion of patients achieving CR or PR, as determined by the investigator. ORRs were confirmed no less than 4 weeks after meeting the criteria for response. Duration of response (DOR) was assessed as a secondary endpoint and was defined as the time from first documentation of CR or PR until the time of first documentation of PD. PFS was assessed as a secondary endpoint and was defined as the time from start of treatment to the earlier date of assessment of progression or death by any cause in the absence of progression, at 2 months and every 2 months thereafter until 1 year. PFS at 6 months and 12 months was also measured. OS was assessed as a secondary endpoint and was defined as the time from the date of start of treatment to the date of death by any cause. DCR (CR+PR+SD) was assessed as a secondary endpoint and was defined as the percentage of patients achieving CR, PR, or SD. Radiographic scans were sent to a central vendor upon acquisition and archived for future evaluation.

Exploratory endpoints included measures of clinical benefit by changes in bacterial colonization and clinical benefit, immune kinetics, and global changes in microbiome and metabolomics.

### Safety Analysis

All enrolled patients who received at least one dose of either VE800 or nivolumab were included in the Safety Analysis Set. Safety data were overseen by a committee comprised of investigators and the study sponsor. Patients were evaluated for dose-limiting toxicity (DLT) for a period of 28 days after starting VE800 plus nivolumab. The first 12 patients to complete the first 28 days of combination treatment or who experienced a DLT were included in the DLT-evaluable population. Safety information was collected throughout the study treatment period. Safety information for analysis included listed symptoms, clinical laboratory parameters (hematology and chemistry), vital signs, echocardiogram parameters, Eastern Cooperative Oncology Group (ECOG) performance status, and physical examinations.

### Microbiome Analysis

Stool samples were collected fresh, transferred to an OMNIgene-GUT (DNA Genotek OMR-200 tube, and resuspended in the preservation buffer according to the manufacturer’s instructions. Stool suspensions were extracted using (MoBio) PowerSoil Pro DNA isolation kit (QIAGEN) at Diversigen, Houston, Texas. Libraries were prepared using the Illumina Nextera XT DNA Library Prep kit (Illumina Inc.) and libraries were sequenced on the Illumina NovaSeq platform at Diversigen using 1×100 base pairs chemistry, with a target read depth of 5 million reads per sample. Stool metagenomic sequences (or reads) were assigned to taxonomy using the *k*-mer-based One Codex platform (https://www.onecodex.com/) with a reference database comprised of the National Center for Biotechnology Information (NCBI), and proprietary Vedanta strain genomes^34^. In total, 54 subjects were included in the analysis (efficacy analysis group, **Supplemental Fig. S1**). The colonization of VE800 strains was evaluated using stool samples collected over the duration of the study from each subject (**Fig. 1A**). Stool sample availability for sequencing is summarized in **Supplementary Table S18**.

VE800 strains were detected in subject stool samples using a specialized bioinformatics assay^20,35^. A set of unique genomic markers spanning the entire length of the strain genome were identified for each of the VE800 organisms. The presence or absence of each VE800 strain in each subject stool sample was determined by the depth and coverage of the unique genomic markers in the metagenomic reads. The relative abundance of individual strains was calculated as the total marker depth per strain (the ratio of metagenomic reads matching any strain marker to the total number of strain markers) divided by the total bases and multiplied by the genome size: RA = (total marker depth/total bases per sample) × genome size. The estimated RA of endogenous bacterial species in the microbial community was determined using the standard One Codex algorithm from quality-filtered metagenomic reads after removal of unclassified reads and reads that map to the human host. At higher taxonomic levels (e.g., Phylum or Class level), the RA was calculated for each sample as the ratio of sequence reads assigned at the desired taxonomic level (plus all reads assigned below) to the total number of assigned reads.

For the TOPOSCORE analysis^25^, VE800-001 stool metagenomic raw reads were profiled using MetaPhlAn4 (version 4.1.1) against the standard ChocoPhlAn SGB database (vJan26)^36^. The TOPOSCORE was calculated as previously described using the MetaPhlAn4 output to quantify the difference of proportions between present SIG1 (37 species) and SIG2 (45 species) bacterial species in each sample plus Akkermansia sp presence within the normal range, as defined in Derosa et al.

### Serum Analyses

Peripheral blood (4 mL) was collected into Serum Separator Clot activator tubes (Greiner Bio-One) and processed as per manufacturer recommendations for serum separation. The separated serum was cryopreserved until processing. Samples from the melanoma cohort from Baseline and Days 1, 14, 30, 56, and 86 were thawed and spun to clarify samples prior to Luminex analysis (ThermoFisher Scientific). Clarified serum samples were analyzed for cytokine concentrations with MILLIPLEX MAP Human Cytokine/Chemokine/Growth Factor Panel A, 48-Plex Premixed Magnetic Bead Panel (Millipore Sigma), MILLIPLEX Human Cytokine Panel Magnetic Bead Panel II (IL-21 and TRAIL only) (Millipore Sigma), and MILLIPLEX MAP TGF-β Magnetic Bead 3-Plex Kit (Millipore Sigma) according to the manufacturer’s instructions on Luminex MAGPIX (ThermoFisher Scientific). The timing and availability of serum samples from each patient is described in **Supplementary Table S18**.

Samples from subjects in the melanoma cohort were transferred to Metabolon, Inc, Morrisville, North Carolina, for analysis. Samples were split into equal parts for analysis on the liquid chromatography with tandem mass spectrometry (LC/MS/MS) and Polar Liquid Chromatography platforms at the following timepoints: Baseline and Days 1, 14, 30, 56, and 86. For bile acid analysis, the Human Serum and Feces Bile Acids Panel (Metabolon, Inc.) was used to measure all major PBAs and SBAs: cholic acid, chenodeoxycholic acid (CDCA), deoxycholic acid (DCA), lithocholic acid (LCA), ursodeoxycholic acid (UDCA), glycocholic acid (GCA), glycochenodeoxycholic acid (GCDCA), glycodeoxycholic acid (GDCA), glycoursodeoxycholic acid (GUDCA), taurocholic acid (TCA), taurochenodeoxycholic acid (TCDCA), taurodeoxycholic acid (TDCA), taurolithocholic acid (TLCA), tauroursodeoxycholic acid (TUDCA), and glycolithocholic acid (GLCA). Eight SCFAs were measured: acetic acid, isobutyric acid, butyric acid, 2-methyl-butyric acid, isovaleric acid, valeric acid, caproic acid, and succinic acid.

### PBMC Analyses

Whole blood was collected in sodium citrate cell preparation tubes (CPT) (BD Biosciences) and inverted 8 to 10 times to disperse anticoagulant. Tubes were centrifuged within 24 hours of collection, following manufacturer’s instructions. Isolated PBMCs were cryopreserved until analysis. PBMC samples from subjects in the melanoma cohort were transferred to Vedanta Biosciences for flow cytometry analysis. Sample availability from subjects for analysis is delineated in **Supplementary Table S18**.

Subject PBMCs were thawed, washed with complete Roswell Park Memorial Institute Medium (cRPMI) made by combining RPMI (Gibco), with fetal bovine serum (FBS) (Gibco), penicillin/streptomycin (Gibco), Glutamax (Gibco) and 2-(4-[2-hydroxyethyl]piperazin-1-yl)ethanesulfonic acid (HEPES) (Gibco), supplemented with extra FBS (total 20%) and Pierce™ Nuclease for Cell Lysis (Thermo Fisher Scientific). Cells were counted using a ViCell Blue cell counter (Beckman Coulter) and resuspended at 1.0 × 10^7^ cells/mL with cRPMI. Then, 1.0 × 10^6^ cells were plated with cRPMI prepared with 2 µL/mL phorbol 12-myristate 13-acetate (PMA) and ionomycin-based Activation Cocktail (Biolegend), 1 µL/mL of protein transport inhibitors Brefeldin A and Monensin (Biolegend), and 25 µL/mL anti-human CD107a (Biolegend). Cells were incubated for 5 hours. After incubation, cells were transferred to a v-bottom plate for flow cytometry staining with cytokine markers. Aliquots of unstimulated 5.0 × 10^5^ cells were used for staining with transcription factor markers. Viability staining was done using Zombie NIR fixable viability kit (Biolegend) at 0.5 µL/mL in phosphate-buffered saline (PBS) (Gibco) for 20 minutes at approximately 4°C with a cooling plate and protected from light. After incubation, cells were washed with PBS supplemented with FBS and ethylenediaminetetraacetic acid (EDTA) (Invitrogen). Cells were stained with surface markers for 35 minutes over a cold plate after blocking with 200 µL/mL Human TruStain FcX (Biolegend). Cells were washed and fixed using the eBioscience forkhead box protein P3 (Foxp3)/Transcription Factor Staining Buffer Set (Thermo Fisher Scientific) overnight at 4°C. After fixing, cells were pelleted and incubated with permeabilization buffer from FOXP3/Transcription Factor Staining Buffer Set for 5 minutes. After permeabilization, cells were stained with intracellular markers for 60 minutes at room temperature and washed prior to acquisition. Gating strategy for the cytokine panel is outlined in **Supplemental Fig. S9** and for the transcription factor panel in **Supplemental Fig. S10**. Flow cytometry data were acquired with the 3 Laser Violet/Blue/Red Aurora Spectral Flow Cytometer by Cytek Biosciences and analyzed using FlowJo™ Software version 10.10 (BD Biosciences). Reagents and antibodies for flow cytometry staining are delineated in **Supplementary Table S19**.

### Statistical Analyses

The original sample size planned for the study was 111 patients: 42 for melanoma, 42 for G/GEJ, and 27 for CRC-MSS, separated into two stages. Due to the ORRs observed across the 3 cohorts in Stage 1, the study did not meet the criteria for continuation and enrollment did not continue for Stage 2.

To model the probability of PFS as a function of VE800 strain colonization, we calculated Kaplan-Meier (KM) curves for subjects with “High Colonization” and “Low Colonization”. Colonization status was determined based on the average number of VE800 strains detected in each patient during the first month of dosing (averaged over the number of strains detected at Days 3, 7, 14, 21, and 30). A cutpoint analysis was conducted to determine meaningful colonization categories, calculating time-to-progression curves and log-rank p-values for all possible colonization cutpoints across the cancer types. In each cohort, progression-free probability was calculated using a Cox proportional hazard model using Colonization as a covariate to draw inferences between PFS and VE800 colonization.

Statistical analysis of microbial diversity, microbial species, serum metabolites, and serum cytokines was performed as follows: Comparisons across different time points were performed using a linear mixed-effects (LME) model, specified as:

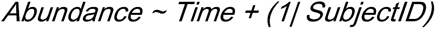

In this model, Abundance represents the RA, concentration, or proportion of various analytes, and the term *(1∣ SubjectID)* accounts for the random effect due to repeated samples.

To compare the responses and engraftment between SD/PD and HC/LC across all analytes, we employed LME models:

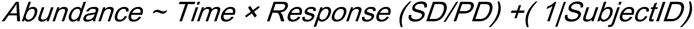

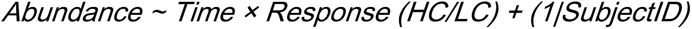

For cross-sectional comparisons at different time points, we used post-hoc contrasts via emmeans (https://github.com/rvlenth/emmeans) to assess differences between any two groups (SD/PD or HC/LC). We report p-values along with adjusted p-values using the Benjamini-Hochberg method for all comparisons.

For ordinal regression analysis, we defined strain colonization in an individual based on detection across samples collected at multiple time points (Days 7, 14, 21, and 30) within a month of VE800 administration. Colonization was treated as an ordinal variable, quantified as the proportion of days the strain was detected. For example, a strain was considered fully colonized in an individual if detected in all four samples within the month, yielding a colonization value of 1. If a strain was detected in three out of four samples, the colonization value was calculated as the fraction of days it was present (e.g., 0.75). In R, we used the ordinalNet package to build ordinal regression models, applying Lasso penalization to improve feature selection and reduce the risk of overfitting. We optimized the regularization parameter (λ) through cross-validation using the *ordinalNetCV* function, selecting the optimal λ based on the average across folds. For each strain, we fitted the model using the cumulative link function and the optimal λ. The cumulative link function models the probability of an observation falling into a given category or higher. The model coefficients indicate how changes in the predictor variables affect these probabilities. Positive coefficients imply that as the predictor value increases, the probability of being in a higher category also increases. Conversely, negative coefficients suggest that as the predictor value increases, the probability of being in a higher category decreases.

Statistical analysis of flow cytometry data obtained from Flowjo software was performed using GraphPad Prism 10 Software (LaJolla, CA, USA) version 10.2.0. Data were grouped by clinical outcome (SD/PD) or colonization of VE800 strains (HC/LC) and organized by timepoints. An LME model was used, including interactions of clinical efficacy and timepoint, with sphericity assumptions of equal variability, without corrections for multiple comparisons. Corrections for multiple comparisons were omitted because changes from baseline were assessed from clinical outcome or colonization groups from a single population at a time. Subsets of populations changed were identified from preclinical data and literature.

## Data Availability

VE800 strain genomes are available under BioProject PRJEB29940. Metagenomic sequencing data are available under BioProject PRJNA1460837. VE800 strain assignments were assessed using the Genome Taxonomy Database Version 207. All other raw data for analyses are available in supplementary tables. Other materials generated in the study will be made available under a material transfer agreement with Vedanta Biosciences. Correspondence should be addressed to and the corresponding authors.

## ACKNOWLEDGEMENTS

We thank all the patients and their families for their participation in this study. We also acknowledge the clinical investigators, and staff members involved from all participating sites for their efforts in conducting this trial. We also thank the members of the independent Data and Safety Monitoring Board for their guidance. We acknowledge the support from Bristol Myers Squibb for their supply of nivolumab and for supporting this clinical trial. The clinical study was sponsored and funded by Vedanta Biosciences, Inc. As the trial sponsor, Vedanta Biosciences designed the study, collected and analyzed the data, and wrote the manuscript. S.K.B. is supported by a sponsored research agreement with Vedanta Biosciences.

## AUTHOR CONTRIBUTIONS

D.B., R.S., K.H. and B.O. conceptualized and designed the study. C.L.D.A., R.M., S.K.B., A.P. and V.B. developed the methodology for statistical and computational analysis. C.L.D.A., R.M., S.K.B., A.P., S.B., E.D., R.S., B.R., D.B., J.L.S., obtained, curated, and analyzed the data. C.L.D.A., R.M., S.K.B., A.P., S.B., E.D., J.M.N., R.S., D.B., B.R., V.B., J.S.W., J.F., J.L.S. and D.D. contributed to the investigation and interpretation of the data. C.L.D.A., R.M. and S.K.B. drafted the manuscript and figures. All authors contributed to substantial revisions of the manuscript. All authors reviewed the manuscript for intellectual content. All authors approved the final version of the submitted manuscript, agreed to be accountable for all aspects, and vouch for data accuracy and completeness.

**Supplementary Figure S1: Patient Disposition.** CONSORT diagram of study patients enrolled.

**Supplementary Figure S2: Relative Abundance of Individual VE800 Strains Overtime**. Line plots indicate the median (solid line) and mean (dashed line) log relative abundance per strain across subjects at the indicated collection timepoints across cohorts. Whiskers represent the standard error in the mean computed across subjects at each timepoint. The grey lines show the abundance of each strain over time within individual subjects.

**Supplementary Figure S3: Relative Abundance of Individual VE800 Strains Overtime in Melanoma Cohort.** Line plots indicate the median (solid line) and mean (dashed line) log relative abundance per strain at the indicated collection timepoints in melanoma subjects characterized as having HC versus LC of VE800 strains. Whiskers represent the standard error in the mean computed across subjects at each timepoint. The grey lines show the abundance of each strain over time within individual subjects

**Supplementary Figure S4: Relative Abundance of Microbiome Species in G/GEJ and CRC-MSS Cohorts.** Heatmap showing the relative abundance of differentially abundant microbiome species (FDR < 0.05) in G/GEJ (Top) and CRC-MSS (Bottom) cohort, as determined by an LME model (Relative Abundance ∼ Time + (1|SubjectID)), comparing each time point to baseline. Red arrows indicate VE800 species relatives.

**Supplementary Figure S5: TOPOSCORE analysis over time by cohort and overall response:** TOPOSCORE analysis of microbiome species over time by cohort, CRC (left), Gastric (middle) and Melanoma (right) and overall responses, SD (orange) and PD (blue). Thin lines represent individual subjects and bold lines represent mean ± standard error. Antibiotics commenced at the baseline sample and were given for five days.

**Supplementary Figure S6: Phylogenetic Tree of Species Associated with Colonization of VE800:** Phylogenetic tree of baseline endogenous species with significant associations to colonization with VE800 strains. Phylogenetic associations identified by color, Firmicutes (now referred to as Bacillota) in purple, Proteobacteria (now referred to as Pseudomonadota) in orange, Actinobacteria (now referred to as Actinomycetota) in green, Bacteroidetes (now referred to as Bacteriodota) in blue, and Verrucomicrobia (now referred to as Verrucomicrobiota) in pink.

**Supplementary Figure S7: SCFA Overtime in Melanoma Cohort.** Heatmaps showing SCFA levels in serum from baseline to Day 30 in subjects based on Clinical Activity (Top) or VE800 Strain Colonization (Bottom). To identify significant changes (FDR<0.05) over time for each analyte, an LME model: Abundance ∼ time **×** Response +(1|SubjectID) was applied. Z-score represents log-transformed abundances over time. The annotation bar on the far right showing the coefficients (effect size) from the model, comparing Days 1, 14, and 30 to baseline, respectively. Coefficient depicts changes from baseline and * indicate statistically significant changes (FDR < 0.05).

**Supplementary Figure S8: Bile Acid Overtime in Melanoma Cohort.** Heatmaps showing BA levels in serum from baseline to Day 30 in subjects based on Clinical Activity (Top) or VE800 Strain Colonization (Bottom). To identify significant changes (FDR<0.05) over time for each analyte, an LME model: Abundance ∼ time **×** Response +(1|SubjectID) was applied. Z-score represents log-transformed abundances over time. The annotation bars on the far right showing the coefficients (effect size) from the model, comparing Days 1, 14, and 30 to baseline, respectively. Coefficient depicts changes from baseline and * indicate statistically significant changes (FDR < 0.05).

**Supplementary Figure S9: Flow Cytometry Gating Strategy for Cytokine Panel.** A) Gating strategy of circulating PBMCs processed with the flow cytometry spectral cytokine panel. First, all cells were gated (side scatter height [SSC-H] vs. forward scatter height [FSC-H]), then through doublet exclusion (SSC-H vs. side scatter area [SSC-A] and FSC-H vs. forward scatter area [FSC-A]), then live cells (SSC-A vs. viability). CD8 and CD4 T cells were identified by gating CD3+ cells (SSC-A vs CD3) and CD8+ or CD4+, respectively (CD8 vs. CD4). Cytokine-secreting NK cells were identified by gating CD3-cells and CD56+ cells that were CD94+. From each population, cytokine-secreting cells were identified by gating cells positive for interferon gamma (IFN-γ), tumor necrosis factor alpha (TNF-α), interleukin (IL)-17A, IL-4, IL-10 and CD107a. B) Representative plots of unstimulated controls used for gating.

**Supplementary Figure S10: Flow Cytometry Gating Strategy for Transcription Factor Panel.** A) Major subsets of immune cells were identified by the following gating strategy: first gating all cells (SSC-H vs FSC-H), then through doublet exclusion (SSC-H vs SSC-A and FSC-H vs FSC-A), then live cells (SSC-A area vs viability). T cells were identified as CD3+ (CD14 vs CD3) and CD4+ or CD8+, respectively. Classical and Intermediate monocytes were identified by gating CD3-cells (CD14 vs CD3) and CD14+CD16- and CD14+/-CD16+, respectively. Myeloid suppressor cells were identified from classical monocytes by gating CD33+CD11b+ cells. B and NK cells were identified by gating CD3-cells and CD19+ CD56-for B cells and CD19-CD56+ for NK cells. B) T cell subsets could be further characterized based on activation/exhaustion. Fluorescence minus one (FMO) controls were used for gating controls for all activation/exhaustion markers.

**Supplementary Figure S11: Flow Cytometry Gating Strategy for T cell Subsets.** T cell subsets were further characterized into cytotoxic T cells (Tc) or helper T cells (Th) and Tregs based on expression of indicated transcription factors. FMO controls were used for gating controls for all transcription factor markers.

**Supplementary Figure 12: Circulating T cell Changes Overtime with Flow Cytometry in Melanoma Cohort.** Frequency of A) IFN-γ CD8 T cells (left) and TNF-α CD8 T cells (right), B)) IFN-γ NK cells (left) and TNF-α NK cells (right), C) CD107a+ CD8 T cells (left) and TNF-α+ IFN-γ+ CD8 T cells (right), D) IL-17a+ CD8 T cells (Left), in subsets with SD (blue) and PD (orange) or HC (green) and LC (purple) across study timepoints. Box plots of baseline subtracted frequencies with minimum to maximum whiskers, LME, p<0.05

**Supplementary Figure 13: Circulating Immune Cells Overtime with Flow Cytometry in Melanoma Cohort.** Frequency of A) NK cells (top right) and CD107a+ NK cells (top left), MAIT cells (bottom left), and Tregs (bottom right) and B) MDSCs, in subsets with SD (blue) and PD (orange) or HC (green) and LC (purple) across study timepoints. Box plots of baseline subtracted frequencies with minimum to maximum whiskers, LME, p<0.05 C) Representative flow cytometry plots of HLA-DR expression in MDSCs. D) Frequency of HLA-DRl^ow^ (left) and HLA-DR^high^ (right) MDSCs in PBMCs from subsets with SD (blue) and PD (orange) or HC (green) and LC (purple) across study timepoints. Box plots with minimum-to-maximum whiskers, LME, p<0.05

