## supplemental figures and tables for "CONSORTiUM-IO: A Phase 1 Study Evaluating VE800 Plus Nivolumab in Treatment of Select Refractory or Metastatic Cancers"

Supplementary Figure S1: Consort Diagram

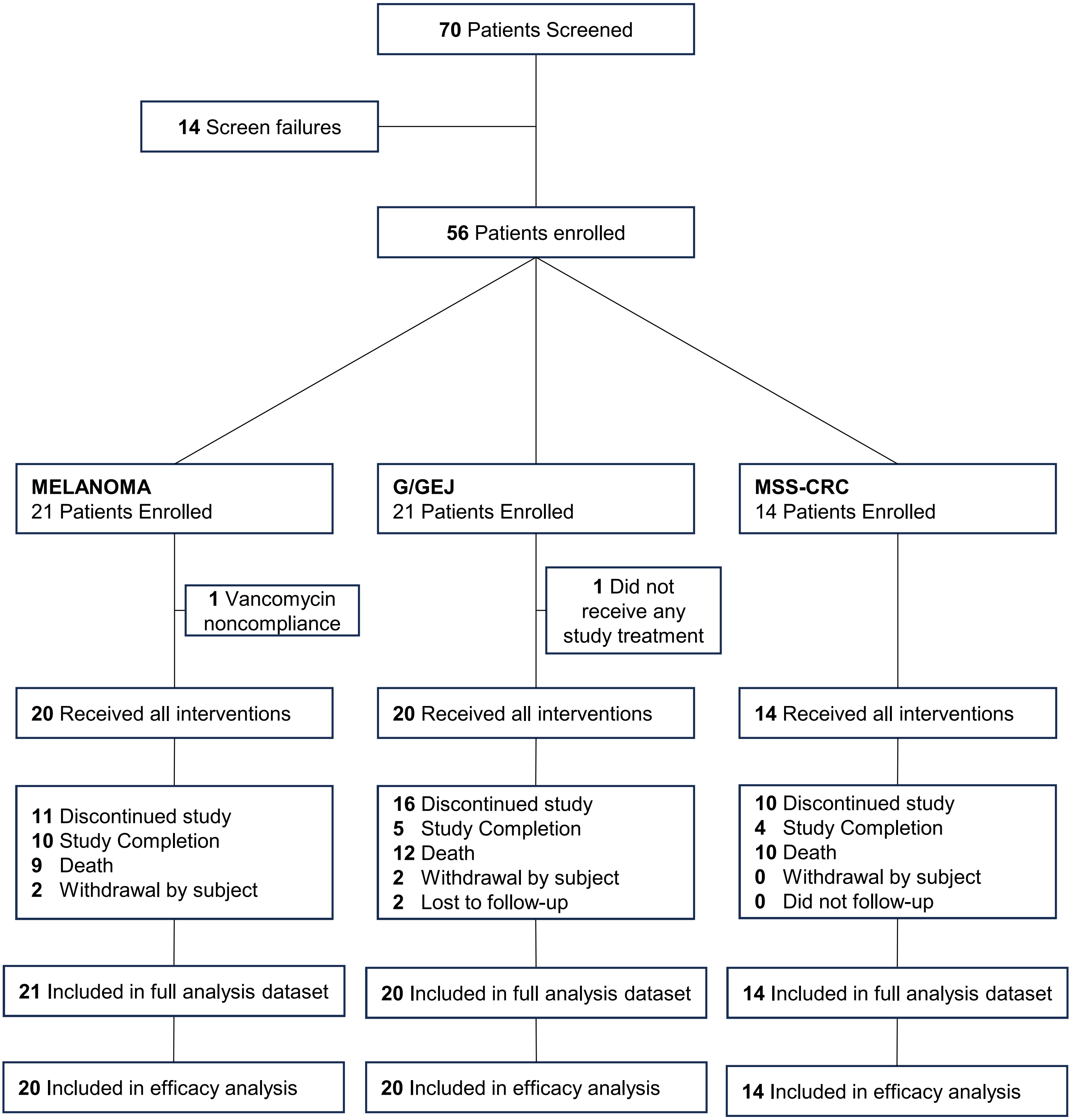

Supplementary Figure S2:
Relative Abundance of Individual VE800 Strains Changes Over Time Across Treatment Groups

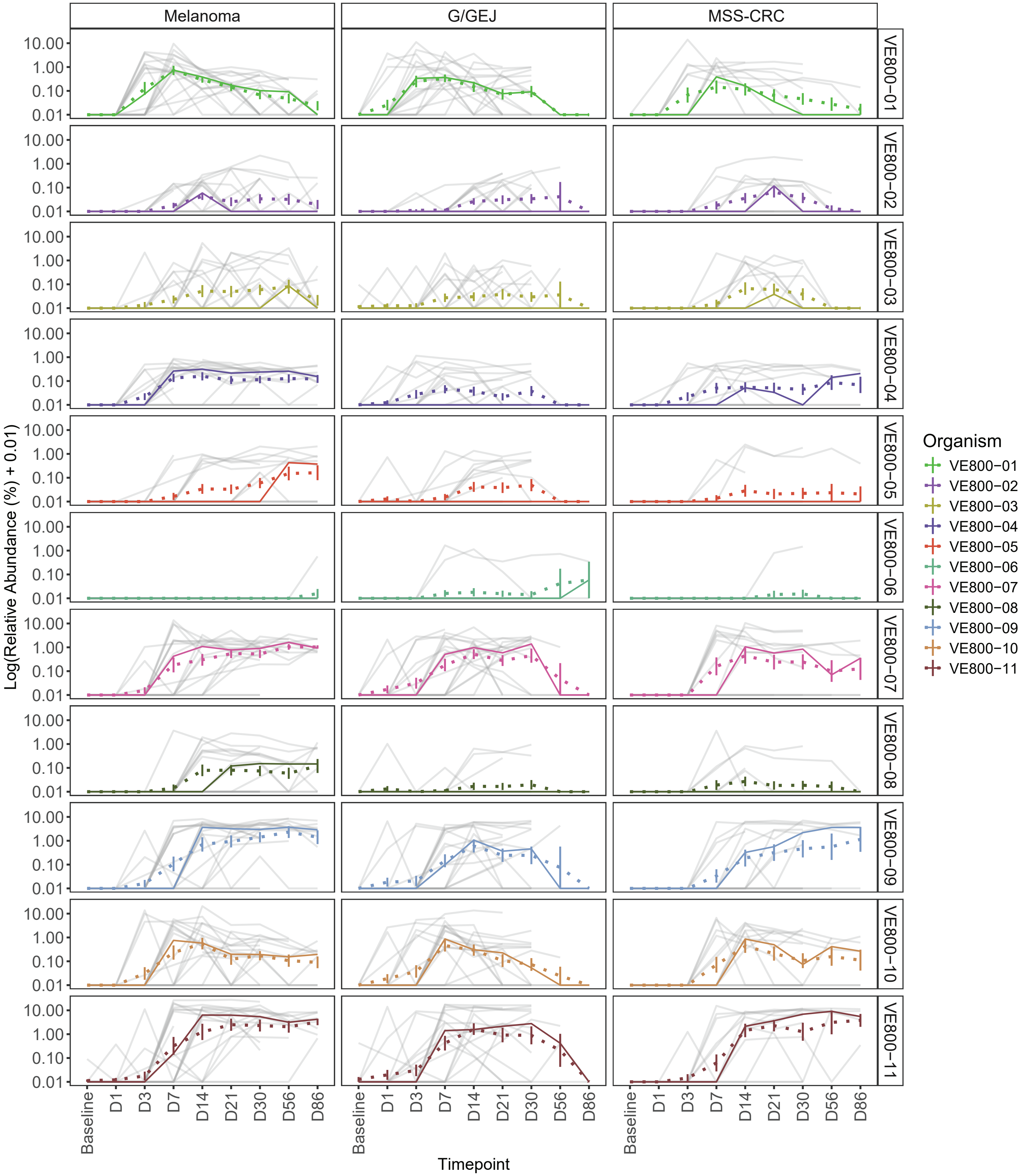

Supplementary Figure S3:
Relative Abundance of Individual VE800 Strains Changes Over Time Across High and Low Colonized Subjects in Melanoma Cohort

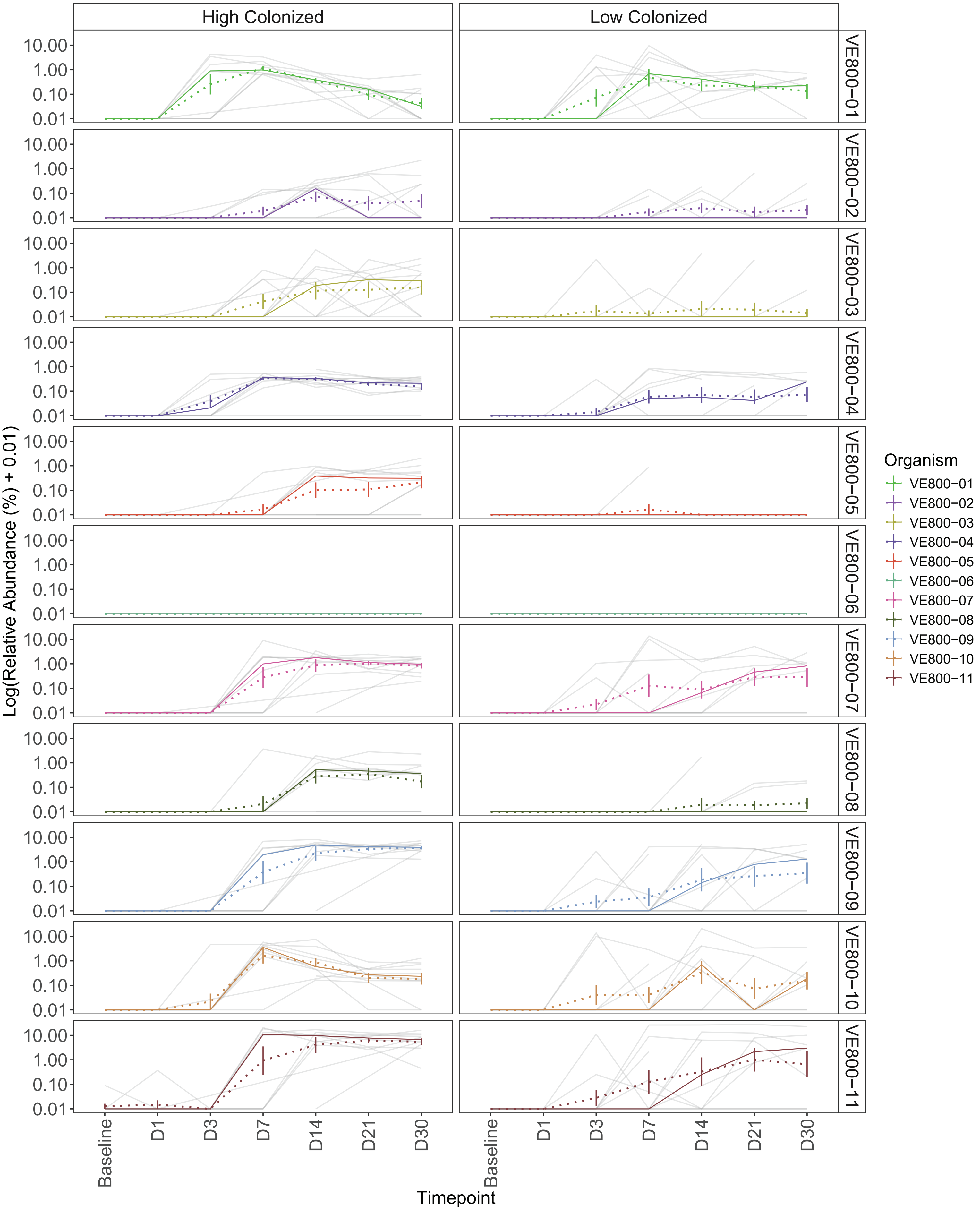

Supplementary Figure S4:
Relative Abundance of Microbiome Species in G/GEJ and MSS-CRC Cohorts Across Timepoints

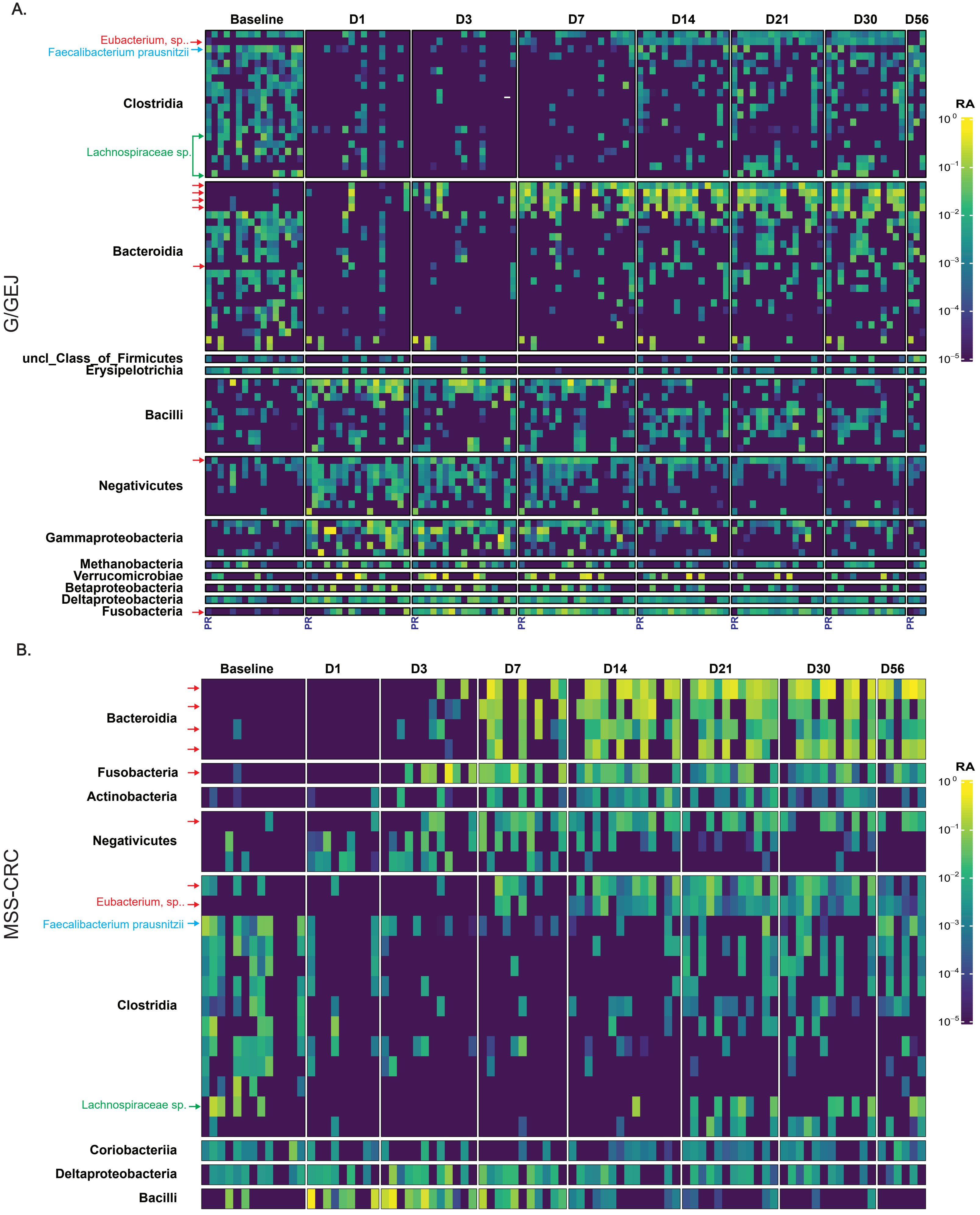

### Supplementary Figure S5: TOPOSCORE over time by cohort and overall response

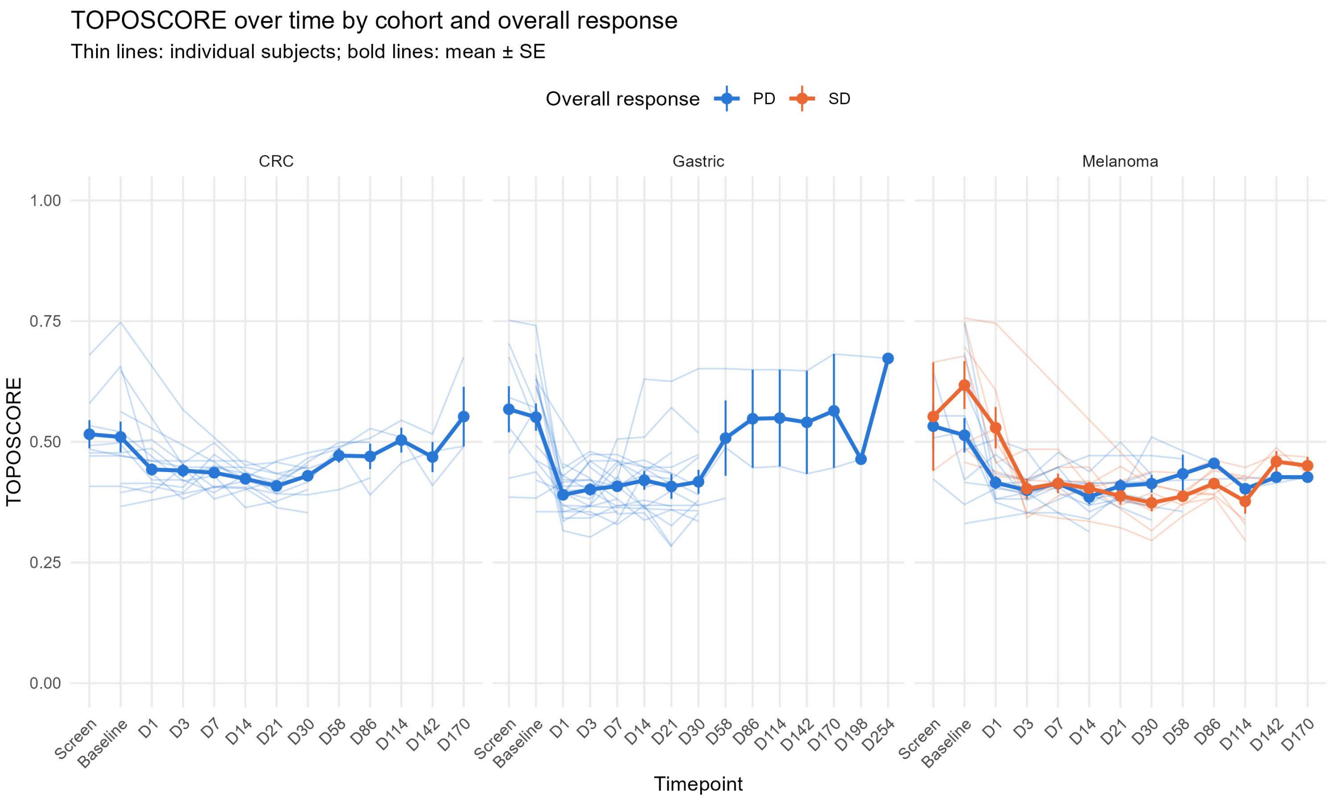

Supplementary Figure S6: Phylogenetic Tree of Species Associated with Colonization of VE800

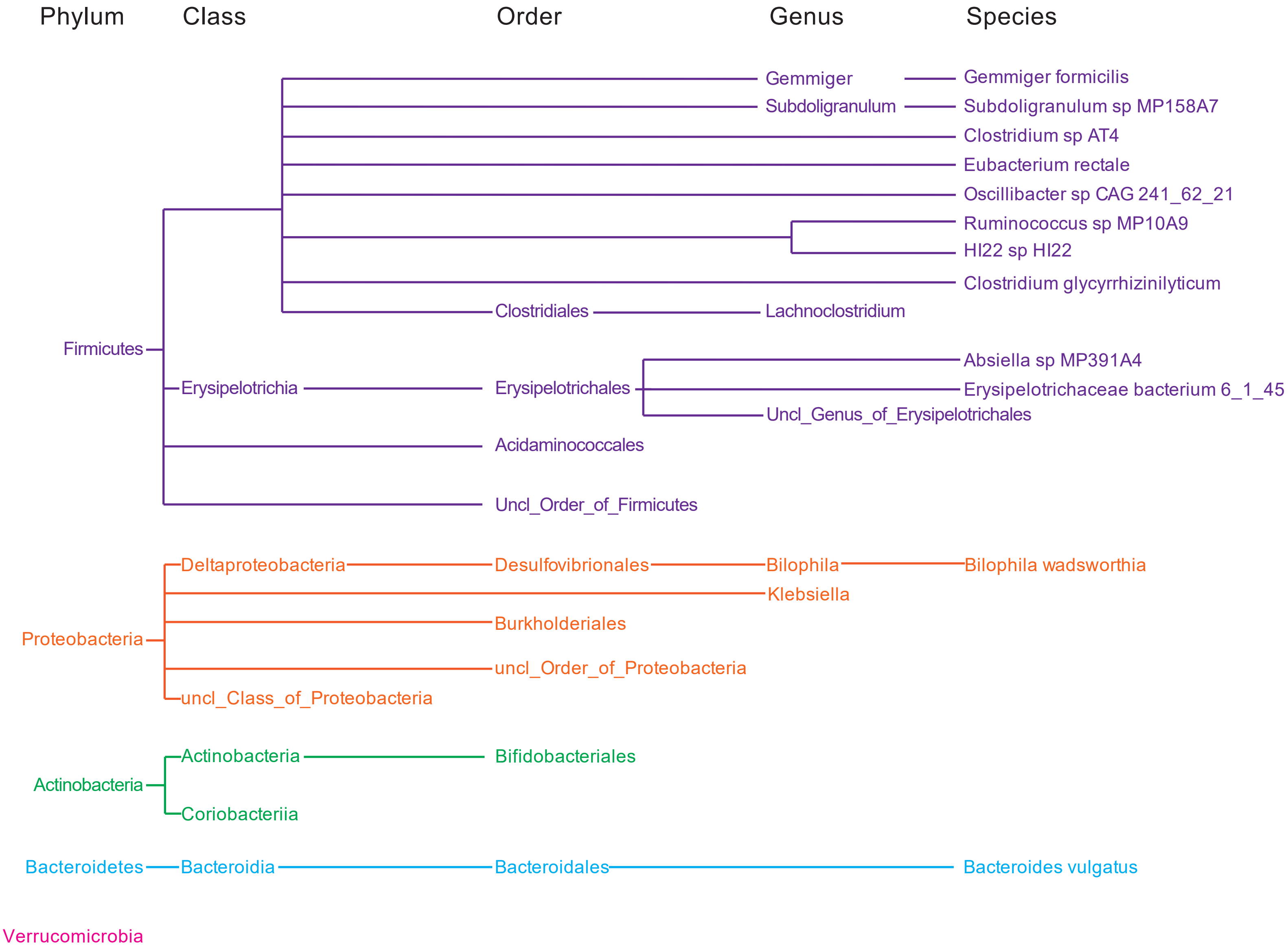

Supplementary Figure S7: SCFA Overtime in Melanoma Cohort

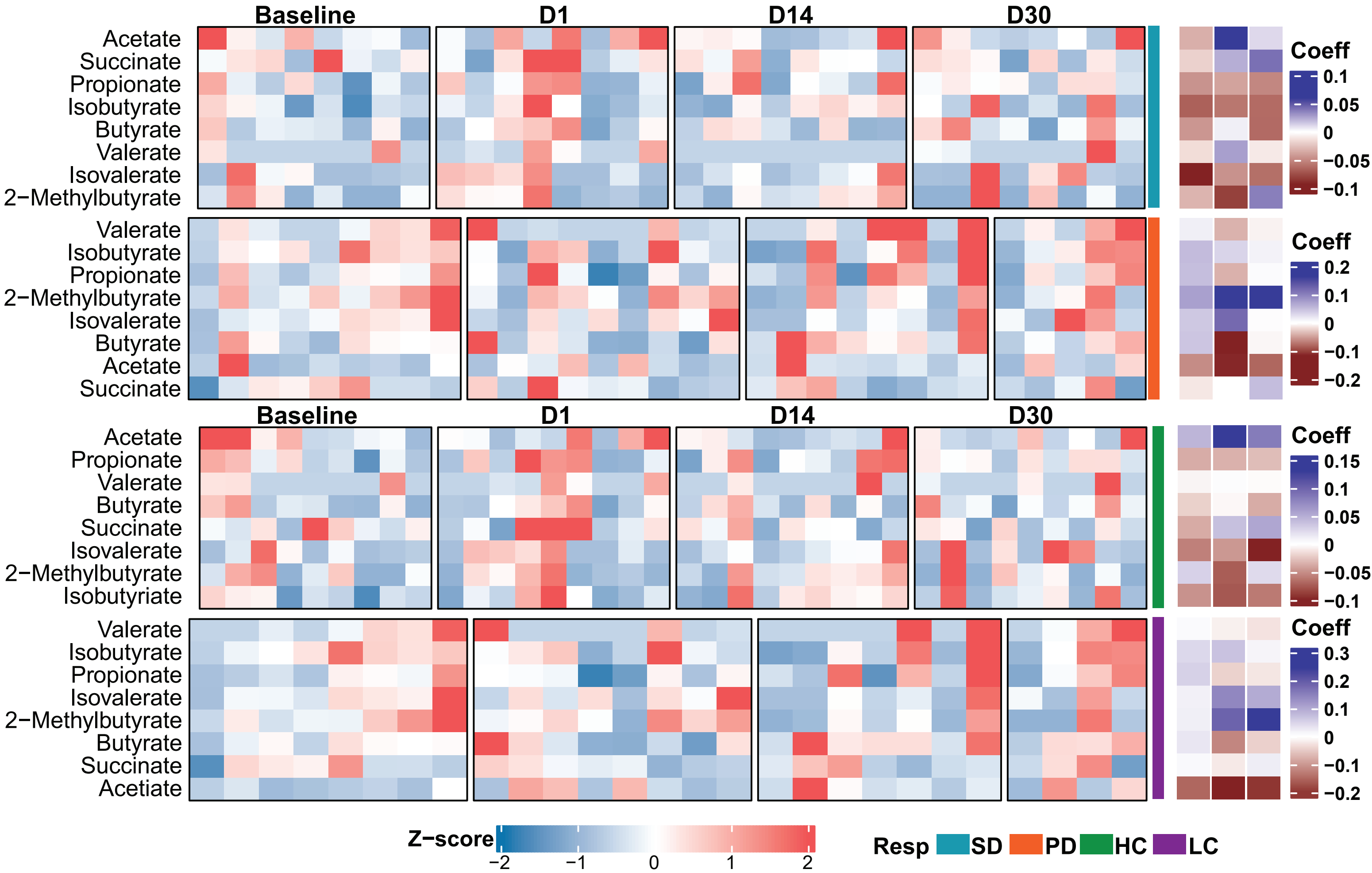

Supplementary Figure S8: Bile Acids Overtime in Melanoma Cohort

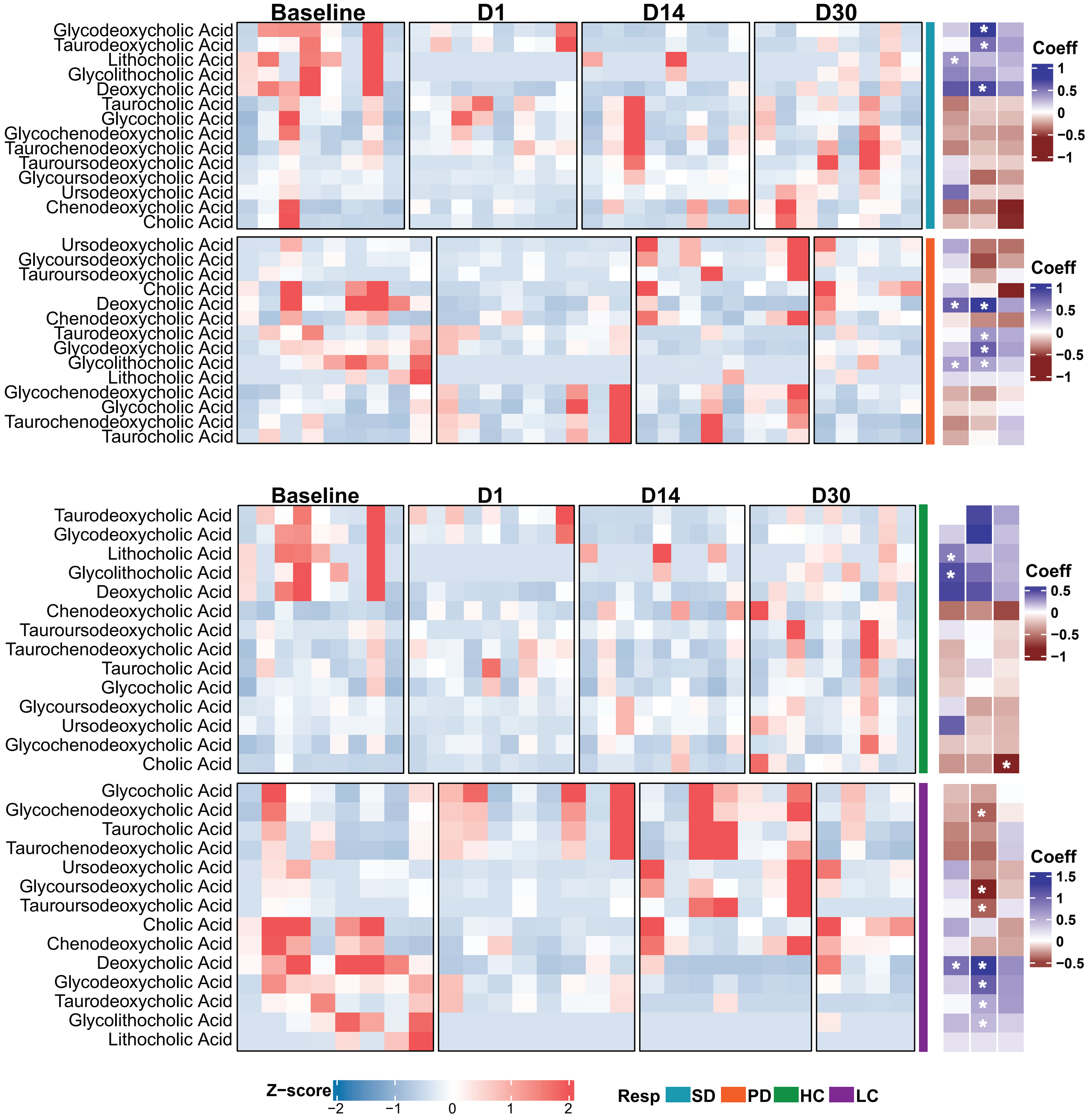

Supplementary Figure S9: Flow Cytometry Gating Strategy for Cytokine Panel

#### Cytokine Panel Gating Strategy

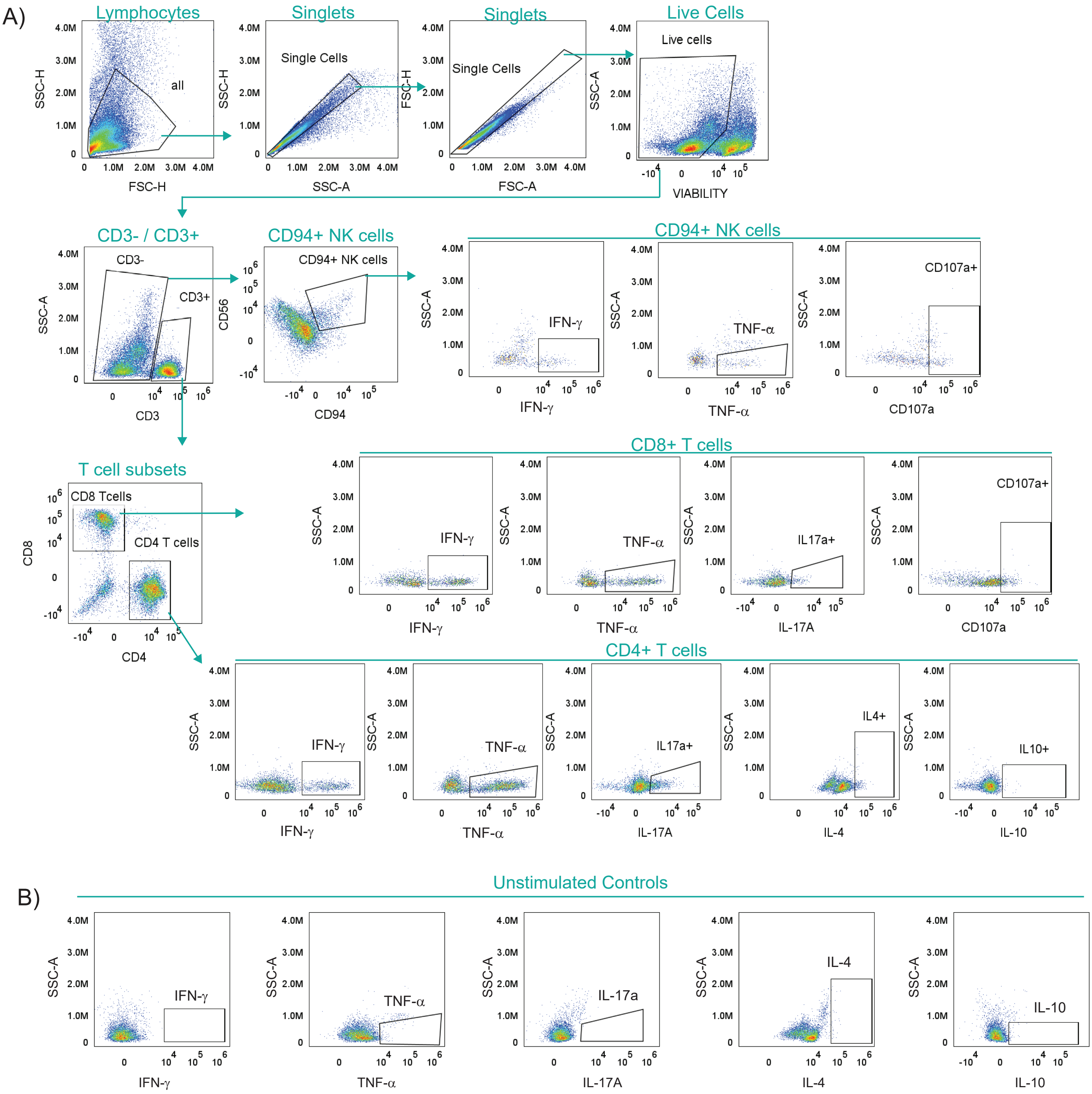

Supplementary Figure S10: Flow Cytometry Gating Strategy for Transcription Factor

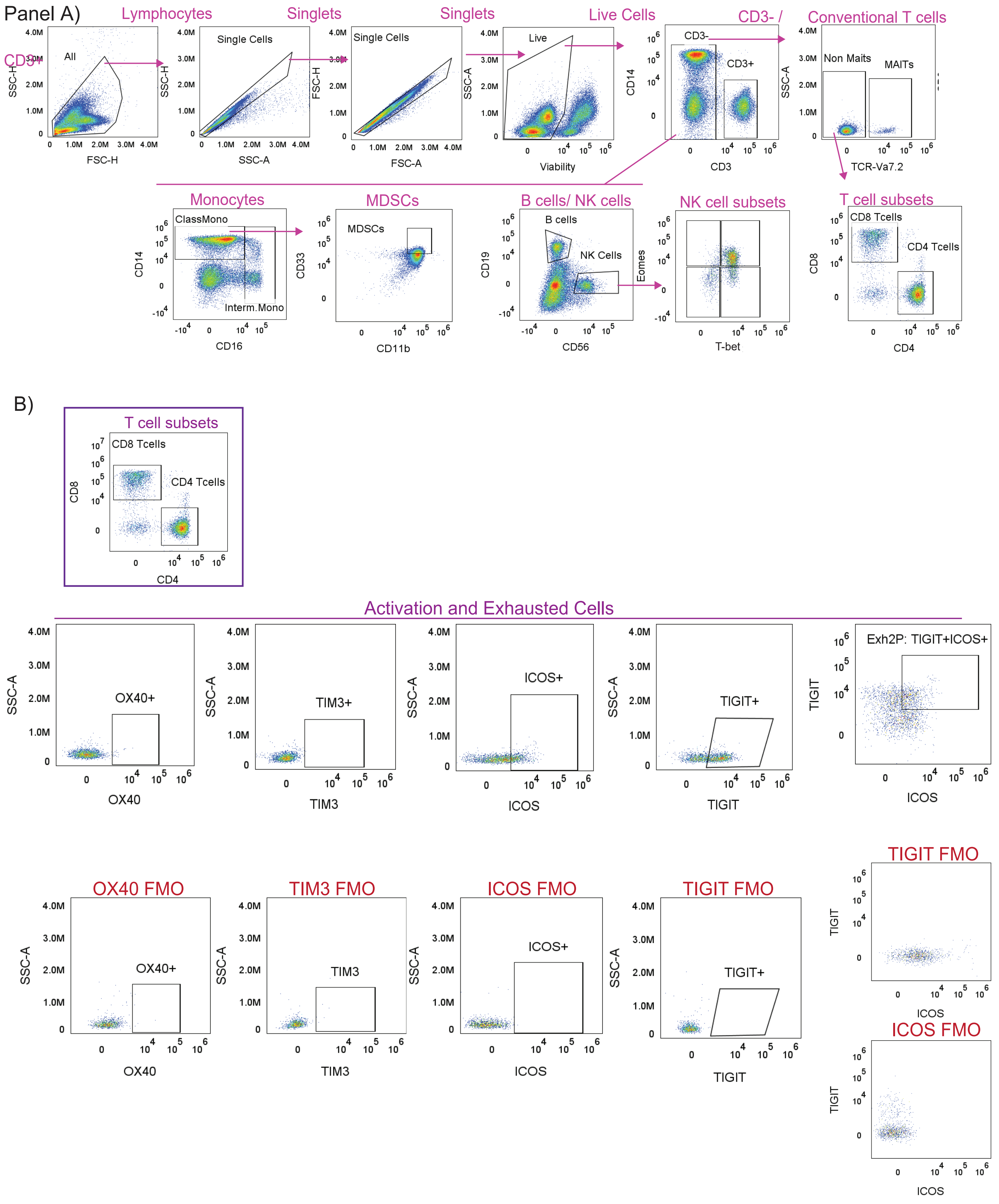

Supplementary Figure S11: Flow Cytometry Gating Strategy for T cell Subsets

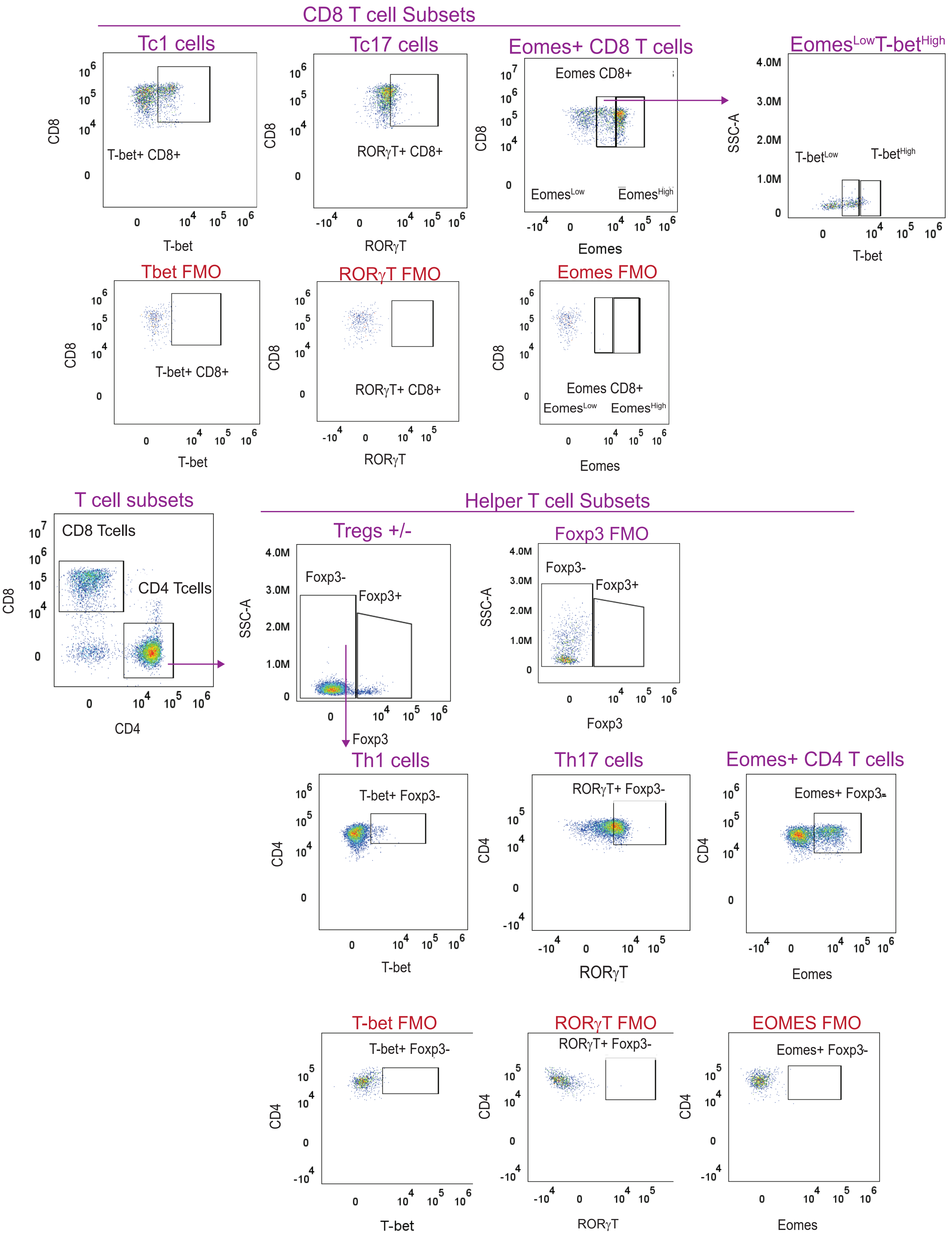

Supplementary Figure S12: Circulating T cell Changes Overtime with Flow Cytometry.

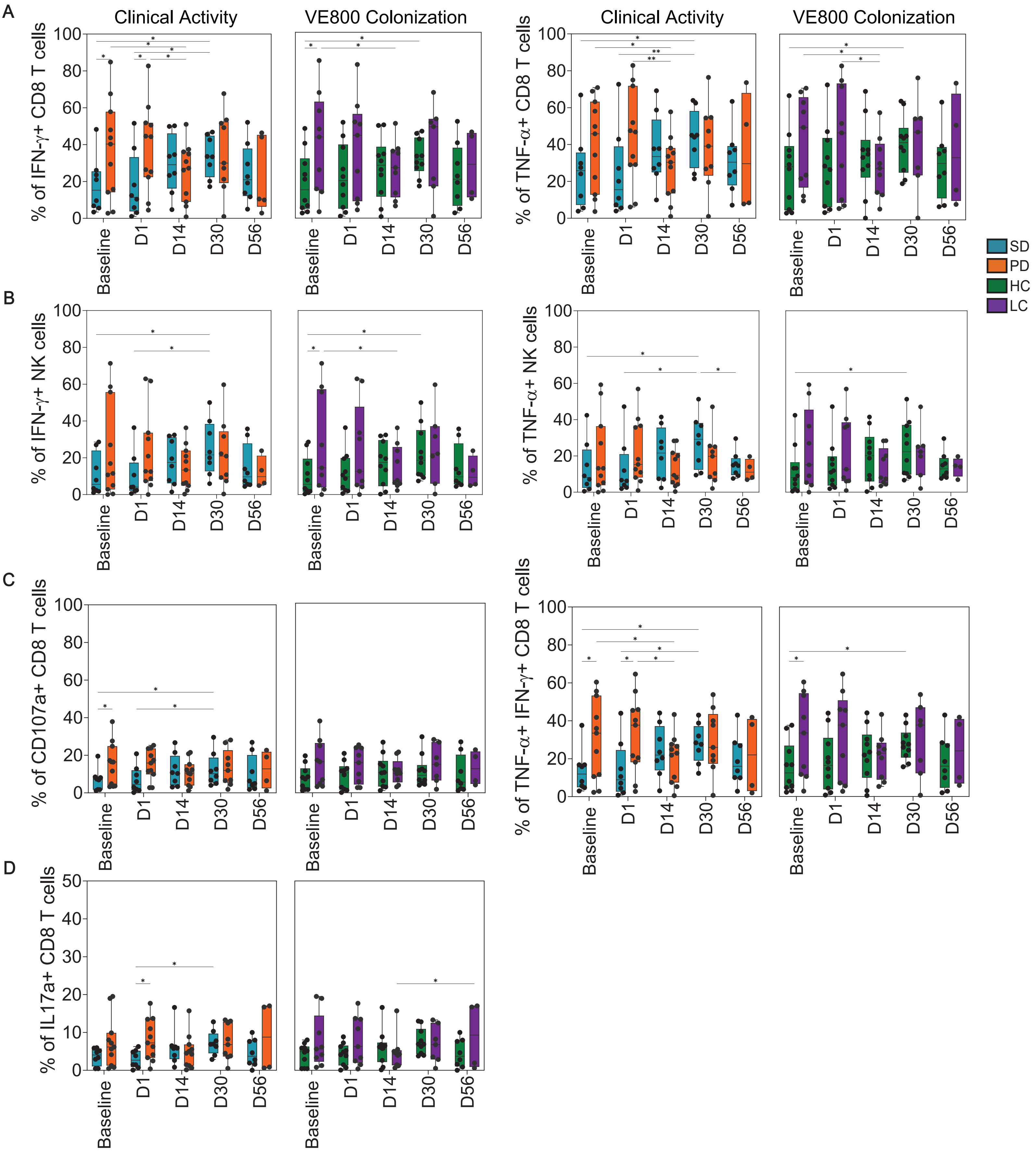

Supplementary Figure S13: Circulating Immune Cells Identified in Melanoma Cohort

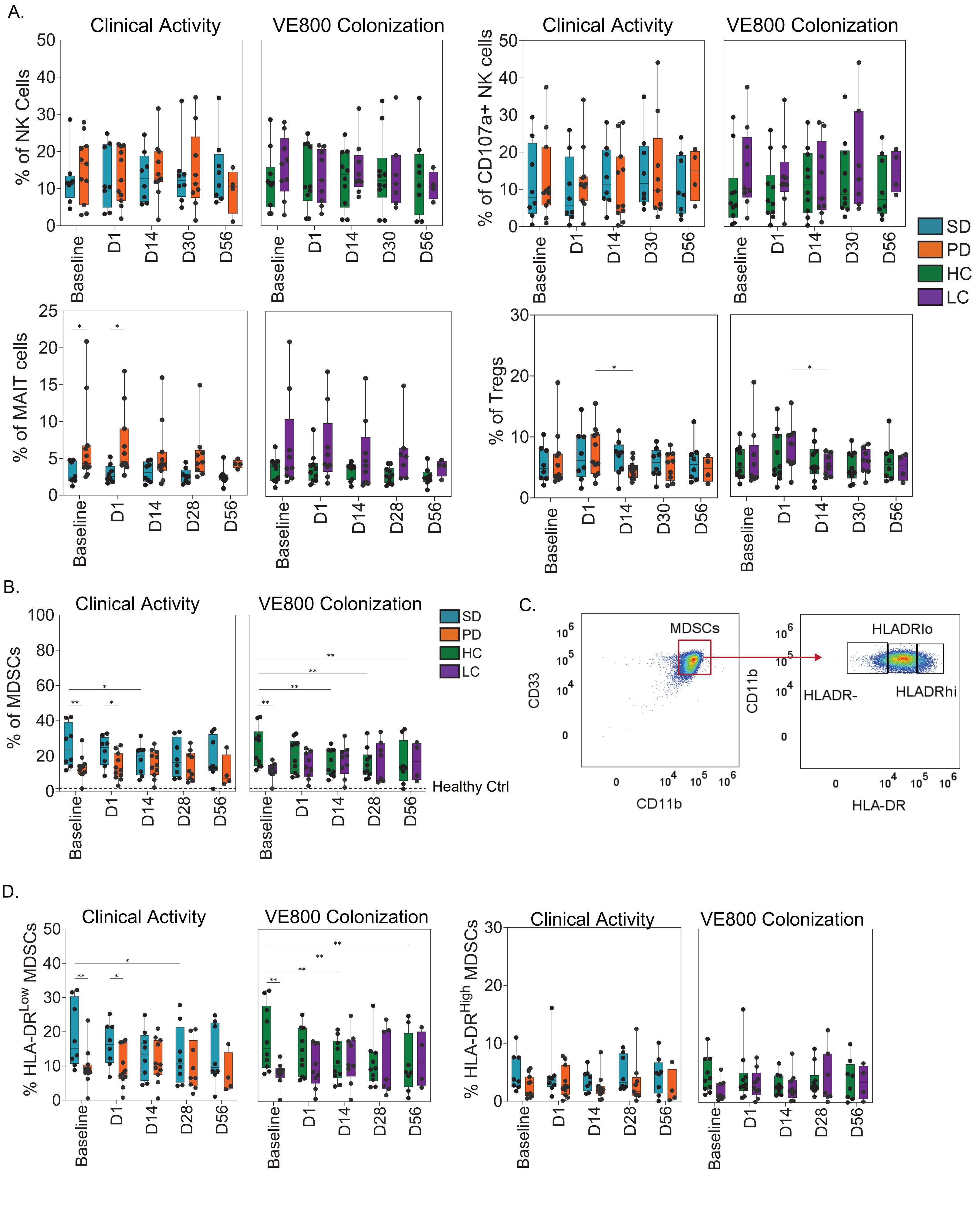

#### SUPPLEMENTARY TABLES

**Supplementary Table S1: Strains in VE800**

| Strain ID | Effector Strain | Nearest Species[1] |
|---|---|---|
| VE800-01 | Yes | *Fusobacterium_A ulcerans_A* |
| VE800-02 | Yes | *Eubacterium limosum* |
| VE800-03 | Yes | *Ruthenibacterium lactatiformans* |
| VE800-04 | Yes | *Phascolarctobacterium faecium* |
| VE800-05 | No | *Parabacteroides johnsonii* |
| VE800-06 | No | *Paraprevotella xylaniphila* |
| VE800-07 | No | *Parabacteroides distasonis* |
| VE800-08 | No | *Alistipes senegalensis* |
| VE800-09 | No | *Phocaeicola dorei* |
| VE800-10 | No | *Parabacteroides timonensis* |
| VE800-11 | No | *Bacteroides uniformis* |

[1] Genome Taxonomy DataBase, release 207 (https://gtdb.ecogenomic.org/); April 2022

**Supplementary Table S2: Patient demographics and characteristics (Full Analysis Dataset)**

| Characteristic | | Melanoma (N=21) | Gastric/GEJ (N=20) | CRC-MSS (N=14) | Total (N=55) |
|---|---|---|---|---|---|
| Age at consent | | | | | |
| | Median | 62 | 61.5 | 59.5 | 62 |
| | Range | 22-87 | 45-79 | 31-75 | 22-87 |
| Sex [ n (%)] | | | | | |
| | Female | 11 (52.4) | 7 (35.0) | 6 (42.9) | 24 (43.6) |
| | Male | 10 (47.6) | 13 (65.0) | 8 (57.1) | 31 (56.4) |
| Race [ n (%)] | | | | | |
| | American Indian or Alaska Native | 0 | 0 | 0 | 0 |
| | Asian | 0 | 1 (5.0) | 0 | 1 (1.8) |
| | Black or African American | 0 | 1 (5.0) | 1 (7.1) | 2 (3.6) |
| | White | 21 (100) | 17 (85.0) | 13 (92.9) | 51 (92.7) |
| | Native Hawaiian or other Pacific Islander | 0 | 0 | 0 | 0 |
| | Other | 0 | 0 | 0 | 0 |
| | Unknown | 0 | 1 (5.0) | 0 | 1 (1.8) |
| Ethnicity [ n (%)] | | | | | |
| | Hispanic or Latino | 1 (4.8) | 0 | 0 | 1 (1.8) |
| | Not Hispanic or Latino | 20 (95.2) | 19 (95.0) | 13 (92.9) | 52 (94.5) |
| | Not Reported | 0 | 1 (5.0) | 0 | 1 (1.8) |
| | Unknown | 0 | 0 | 1 (7.1) | 1 (1.8) |
| Disease Stage [ n (%)][a] | | | | | |
| | Metastatic | 21 (100) | 15 (75.0) | 14 (100) | 50 (90.9) |
| | Locally Advanced | 0 | 5 (25.0) | 0 | 5 (9.1) |
| Prior Systemic Therapy Regimens | | | | | |
| | Median | 2.0 | 2.0 | 3.0 | 2.0 |
| Prior Immune Checkpoint Inhibitor [n (%)] | | 21 (100) | 0 | 0 | 21 (38.2) |
| | Nivolumab | 16 (76.2) | NA | NA | 16 (29.1) |
| | Ipilimumab | 14 (66.7) | NA | NA | 14 (25.5) |
| | Pembrolizumab | 8 (38.1) | NA | NA | 8 (14.5) |

[a] As determined by local investigator

**Supplementary Table S3. Summary of Treatment-Emergent Adverse Events (TEAEs) and Treatment-Related Adverse Events (TRAEs) by Preferred Term (Efficacy Analysis Dataset)**

| **Adverse event[a] [ n (%)]** | **Melanoma (N=20)** | **G/GEJ (N=20)** | **CRC-MSS (N=14)** | **Total (N=54)** |
|---|---|---|---|---|
| Any TEAEs[b] | 16 (80.0) | 19 (95.0) | 12 (85.7) | 47 (87.0) |
| Any TRAEs | 9 (45.0) | 9 (45.0) | 6 (42.9) | 24 (44.4) |
| Vancomycin-related | 1 (5.0) | 3 (15.0) | 1 (7.1) | 5 (9.3) |
| Nivolumab-related | 8 (40.0) | 8 (40.0) | 5 (35.7) | 21 (38.9) |
| VE800-related | 3 (15.0) | 3 (15.0) | 3 (21.4) | 9 (16.7) |
| Any Grade 3 or Greater Adverse Event | 6 (30.0) | 8 (40.0) | 2 (14.3) | 16 (29.6) |
| Vancomycin-related | 0 | 1 (5.0) | 0 | 1 (1.9) |
| Nivolumab-related | 0 | 3 (15.0) | 0 | 3 (5.6) |
| VE800-related | 0 | 1 (5.0) | 0 | 1 (1.9) |
| TRAEs reported in ≥2 patients | | | | |
| Diarrhea | 4 (20.0) | 1 (5.0) | 1 (7.1) | 6 (11.1) |
| Nausea | 1 (5.0) | 3 (15.0) | 0 | 4 (7.1) |
| Vomiting | 0 | 3 (15.0) | 0 | 3 (5.6) |
| Fatigue | 4 (20.0) | 1 (5.0) | 1 (7.1) | 6 (11.1) |
| Pruritus | 2 (10.0) | 2 (10.0) | 0 | 4 (7.4) |
| Rash | 1 (5.0) | 1 (5.0) | 0 | 2 (3.7) |
| Infusion-related reaction | 0 | 0 | 2.0 (14.3) | 2 (3.7) |
| Myalgia | 0 | 1 (5.0) | 1 (7.1) | 2 (3.7) |
| Pleural effusion | 0 | 1 (5.0) | 1 (7.1) | 2 (3.7) |
| Alanine aminotransferase increased | 0 | 1 (5.0) | 1 (7.1) | 2 (3.7) |
| Aspartate aminotransferase increased | 0 | 1 (5.0) | 1 (7.1) | 2 (3.7) |

[a] For each category, patients were included only once, even if they experienced multiple events in that category.

[b] Treatment-emergent is defined as beginning after the first dose of any study treatment, prior to 28 days after the last dose of any study treatment, and prior to the initiation of follow-up systemic anti-cancer therapy.

**Supplementary Table S4. Summary of Clinical Efficacy (Efficacy Analysis Dataset)**

| | **Melanoma (N=20)** | **G/GEJ (N=20)** | **CRC-MSS (N=14)** | **Total (N=54)** |
|---|---|---|---|---|
| Best Overall Response (BOR) (n [%]) | | | | |
| Complete Response (CR) | 0 | 0 | 0 | 0 |
| Partial Response (PR) | 0 | 1 (5.0) | 0 | 1 (1.9) |
| Stable Disease (SD) | 9 (45.0) | 2 (10.0) | 3 (21.4) | 14 (25.9) |
| Progressive Disease (PD) | 9 (45.0) | 16 (80.0) | 10 (71.4) | 35 (64.8) |
| Non evaluable (NE) | 0 | 0 | 0 | 0 |
| Unknown[a] | 2 (10.0) | 1 (5.0) | 1 (7.1) | 4 (7.4) |
| Objective Response Rate (ORR)[b] | 0 | 1 (5.0) | 0 | 1 (1.9) |
| Disease Control Rate (DCR)[c] | 9 (45.0) | 3 (15.0) | 3 (21.4) | 15 (27.8) |
| Duration of Response (DOR) (months)[d] | NA | 5.6 | NA | 5.6 |
| Median Progression-Free Survival (PFS) (months)[e] | 1.9 | 1.8 | 1.8 | 1.8 |
| Min, Max (months) | 0.0, 11.1 | 0.0, 9.1 | 0.0, 7.2 | 0.0, 11.1 |
| 6-month PFS (%) | 23.5 | 14.0 | 7.7 | 14.9 |
| Median Overall survival (OS) (months)[f] | NA | 5.8 | 7.6 | 9.4 |
| Min, Max (months) | 1.1, 15.0 | 1.4, 16.1 | 1.0, 16.8 | 1.0, 16.8 |
| 6-month OS (%) | 79.2 | 46.5 | 70.7 | 65.5 |

[a] Patients in the Efficacy Set who did not have a postbaseline tumor assessment were classified as “Unknown” and “Non-Responders” for ORR, and DCR.

[b] ORR is defined as a BOR of CR or PR from C1/D1 until disease progression or start of new anti-cancer therapy, whichever comes first. ORR will be confirmed no less than 4 weeks after criteria of response are met.

[c] DCR is defined as a BOR of CR, PR or SD from C1/D1 until disease progression or start of new anti-cancer therapy, whichever comes first. CR and PR will be confirmed no less than 4 week after criteria of response are met.

[d] DOR is defined for patients with confirmed CR or PR at the time from date of the first occurrence of a documented confirmed objective tumor response to PD or death due to any cause, whichever comes first. Patients who have not progressed or died at time of analysis were censored at the last date of response assessment. NA indicates no patients exhibited CR or PR in the cohort.

[e] PFS is defined for as time from C1/D1 to progression or death due to any cause, whichever comes first. Patients were censored from PFS analysis if: not progressed or died at time of analysis, progressed or died after two or more consecutive missed scheduled visits, do not have evaluable post-baseline assessment. Obtained via Kaplan Meier estimation, Brookmeyer and Crowley 1982 method[35].

[f] OS defined as time from Cycle1/Day 1 to death due to any cause or loss to follow up. Patients who were alive at the time of analysis were censored on the last date they were reported to be alive. Obtained via Kaplan Meier estimation, Brookmeyer and Crowley 1982 method[35]. NA indicates >50% of the patients in a cohort remain alive.

**Supplementary Table 5: Summary of Subjects (n) Included Per Treatment Group for Pharmacodynamics Analysis Based on Week 8 Clinical Assessments**

| Treatment Group | Subjects (n) | Stable Disease (SD) | Progressive Disease (PD) |
|---|---|---|---|
| CRC-MSS | 14 | 3 | 11 |
| G/GEJ | 20 | 3 | 17 |
| Melanoma | 20 | 8 | 12 |

**Supplementary Table S6: Summary of Sample Availability for Pharmacodynamic Analyses**

| | STOOL | | | SERUM | PBMC |
|---|---|---|---|---|---|
| **Time-point** | **CRC-MSS (n samples)** | **G/GEJ (n samples)** | **Melanoma (n samples)** | **Melanoma (n samples)** | **Melanoma (n samples)** |
| Baseline | 13 | 16 | 19 | 16 | 22 |
| D1 | 9 | 17 | 19 | 16 | 19 |
| D3 | 13 | 17 | 18 | na | na |
| D7 | 13 | 19 | 17 | na | na |
| D14 | 13 | 15 | 17 | 15 | 18 |
| D21 | 12 | 14 | 16 | Na | na |
| D30 | 12 | 12 | 17 | 13 | 18 |
| D56 | 6 | 3 | 11 | 7 | 12 |
| D86 | 5 | 2 | 9 | 6 | 9 |
| D114 | 3 | 2 | 7 | na | na |
| D142 | 3 | 2 | 4 | na | na |
| D170 | 3 | 2 | 3 | na | na |
| EOT | 6 | 6 | 4 | na | na |

**Supplementary Table S7: Comparison of VE800 Strain Relative Abundance and Detection from Baseline to Day 14 Across Cohorts**

| Cohort | Species | p-value | Adjusted p-value | Statistic |
|---|---|---|---|---|
| CRC-MSS | VE800-01 | 0.000320 | 0.00088 | -5.15 |
| | VE800-02 | 0.000036 | 0.00013 | -6.65 |
| | VE800-03 | 0.00096 | 0.0021 | -4.46 |
| | VE800-04 | 0.0015 | 0.0028 | -4.18 |
| | VE800-05 | 0.20 | 0.22 | -1.36 |
| | VE800-06 | 0.34 | 0.34 | 1.00 |
| | VE800-07 | 0.0044 | 0.0070 | -3.5641 |
| | VE800-08 | 0.10 | 0.12 | -1.81 |
| | VE800-09 | 0.03 | 0.036 | -2.57 |
| | VE800-10 | 0.000003 | 0.000038 | -8.54 |
| | VE800-11 | 0.000008 | 0.000046 | -7.79 |
| | total VE800 abundance | 7.9E-05 | no p-adjustment | -6.09 |
| | total VE800 detection | 0.0025 | no p-adjustment | W=0 |
| G/GEJ | VE800-01 | 1.03E-10 | 6.54E-10 | -20.53 |
| | VE800-02 | 3.91E-09 | 1.43E-08 | -14.99 |
| | VE800-03 | 0.0092 | 0.013 | -3.10 |
| | VE800-04 | 0.0029 | 0.0046 | -3.72 |
| | VE800-05 | 0.22 | 0.22 | -1.31 |
| | VE800-06 | 0.17 | 0.18 | -1.48 |
| | VE800-07 | 1.37E-05 | 3.77E-05 | -7.03 |
| | VE800-08 | 0.12 | 0.14 | -1.68 |
| | VE800-09 | 0.00063 | 0.0014 | -4.59 |
| | VE800-10 | 1.19E-10 | 6.54E-10 | -20.28 |
| | VE800-11 | 0.0023 | 0.0042 | -3.85 |
| | total VE800 abundance | 2.14E-06 | no p-adjustment | -8.45 |
| | total VE800 detection | 0.0016 | no p-adjustment | W=0 |
| Melanoma | VE800-01 | 2.96E-16 | 3.26E-15 | -37.576 |
| | VE800-02 | 1.13E-05 | 3.12E-05 | -6.430 |
| | VE800-03 | 0.0013 | 0.0018 | -3.949 |
| | VE800-04 | 4.88E-05 | 8.95E-05 | -5.620 |
| | VE800-05 | 0.074 | 0.081 | -1.922 |
| | VE800-06 | 0.33 | 0.33 | 1.000 |
| | VE800-07 | 7.98E-06 | 2.93E-05 | -6.632 |
| | VE800-08 | 0.011 | 0.014 | -2.897 |
| | VE800-09 | 0.00075 | 0.0012 | -4.218 |
| | VE800-10 | 1.43E-06 | 7.89E-06 | -7.673 |
| | VE800-11 | 4.35E-05 | 8.95E-05 | -5.682 |
| | total VE800 abundance | 2.87E-07 | no p-adjustment | -8.736 |
| | total VE800 detection | 0.00042 | no p-adjustment | W=0 |

Pairwise tests comparing VE800 colonization at Baseline and Day 14 across cohorts. Statistical analysis shown for individual VE800 strain abundance (paired t-test), total abundance (paired t-test), and number of strains detected (Wilcox signed rank test). A negative t-statistic value indicates an increase at Day 14 compared with Baseline, while a W-statistic W=0 indicates a strong and consistent directional difference with all observations increased at Day 14 compared to Baseline.

**Supplementary Table S8: Progression-free Survival Probability in Subjects with High vs Low Colonization**

| Cancer type | Term (compared to High Colonized) | Effect Size | standard error | statistic | *p*-value | Hazard ratio |
|---|---|---|---|---|---|---|
| All cancers | Low Colonized | 0.510 | 0.296 | 1.724 | 0.085 | 1.67 |
| Colorectal | Low Colonized | 0.033 | 0.605 | 0.055 | 0.956 | 1.03 |
| G/GEJ | Low Colonized | -0.128 | 0.466 | -0.274 | 0.784 | 0.88 |
| Melanoma | Low Colonized | 0.932 | 0.519 | 1.794 | 0.073 | 2.54 |

Results of Cox models for PFS as a function of VE800 strain colonization. A positive value for the Effect Size indicates increased probability of PFS in subjects in High Colonized group.

**Supplementary Table S9: Cox Model Summary: Cut-point Threshold Analysis for High vs. Low Colonization in Melanoma Subjects with High vs Low Colonization**

| Cut-point | n (High Colonized) | n (Low Colonized) | Effect Size | standard error | statistic | *p*-value | Hazard ratio |
|---|---|---|---|---|---|---|---|
| Low = N colonized < 1 | 20 | 0 | NA, model does not converge due to unbalanced groups | | | | |
| Low = N colonized < 2 | 19 | 1 | NA, model does not converge due to unbalanced groups | | | | |
| Low = N colonized < 3 | 18 | 2 | -0.699 | 1.035 | -0.675 | 0.499 | 0.497 |
| Low = N colonized < 4 | 16 | 4 | -0.135 | 0.641 | -0.211 | 0.833 | 0.874 |
| Low = N colonized < 5 | 10 | 10 | 0.932 | 0.519 | 1.794 | 0.073 | 2.540 |
| Low = N colonized < 6 | 6 | 14 | 1.285 | 0.650 | 1.978 | 0.048 | 3.615 |
| Low = N colonized < 7 | 4 | 16 | 1.246 | 0.762 | 1.636 | 0.102 | 3.478 |
| Low = N colonized < 8 | 1 | 19 | NA, model does not converge due to unbalanced groups | | | | |
| Low = N colonized < 9 | 0 | 20 | NA, model does not converge due to unbalanced groups | | | | |
| Low = N colonized < 10 | 0 | 20 | NA, model does not converge due to unbalanced groups | | | | |
| Low = N colonized < 11 | 0 | 20 | NA, model does not converge due to unbalanced groups | | | | |

Summary of Cox models evaluating PFS as a function of colonization for different colonization thresholds. A hazard ratio > 1 indicates higher recurrence probability in low colonized vs high colonized subjects.

**Supplementary Table S10: VE800 Strains With Differential Abundance In Melanoma Subjects With High Vs Low Colonization**

| Species | Estimate | Std. Error | p-value | Adjusted p-value |
|---|---|---|---|---|
| VE800-08 | 1.15 | 0.27 | 0.00041 | 0.0030 |
| VE800-03 | 0.94 | 0.26 | 0.00055 | 0.0030 |
| VE800-05 | 1.10 | 0.27 | 0.00092 | 0.0034 |
| VE800-09 | 1.17 | 0.35 | 0.0013 | 0.0037 |
| VE800-04 | 0.98 | 0.32 | 0.0062 | 0.014 |
| VE800-11 | 0.97 | 0.41 | 0.026 | 0.048 |
| VE800-10 | 0.80 | 0.36 | 0.039 | 0.061 |
| VE800-02 | 0.44 | 0.22 | 0.052 | 0.071 |
| VE800-07 | 0.69 | 0.38 | 0.084 | 0.10 |
| VE800-01 | 0.09 | 0.27 | 0.75 | 0.82 |
| VE800-06 | 0 | 3.87E-17 | 1 | 1 |

#### Supplementary Table S11: Shannon Diversity of All Cohorts

| | | Value | Std.Error | DF | t.value | p.value | Var | padj | Visit. Name. [illegible] |
|---|---|---|---|---|---|---|---|---|---|
| Melanoma | Visit.Name. NormD1 | -0.943112 | 0.167577 | 107 | -5.628 | 1.48E-07 | Visit.Name. NormD1 | 3.449E-07 | D1 |
| | Visit.Name. NormD3 | -1.249087 | 0.170264 | 107 | -7.336 | 4.46E-11 | Visit.Name. NormD3 | 3.122E-10 | D3 |
| | Visit.Name. NormD7 | -1.158045 | 0.173118 | 107 | -6.689 | 1.06E-09 | Visit.Name. NormD7 | 3.702E-09 | D7 |
| | Visit.Name. NormD14 | -0.948338 | 0.174001 | 107 | -5.450 | 3.25E-07 | Visit.Name. NormD14 | 5.689E-07 | D14 |
| | Visit.Name. NormD21 | -0.613557 | 0.176384 | 107 | -3.479 | 7.30E-04 | Visit.Name. NormD21 | 1.022E-03 | D21 |
| | Visit.Name. NormD30 | -0.401474 | 0.173962 | 107 | -2.308 | 2.29E-02 | Visit.Name. NormD30 | 2.675E-02 | D30 |
| | Visit.Name. NormD56 | -0.411970 | 0.200147 | 107 | -2.058 | 4.20E-02 | Visit.Name. NormD56 | 4.199E-02 | D56 |
| Gastric | Visit.Name. NormD1 | -1.319537 | 0.192850 | 88 | -6.842 | 9.95E-10 | Visit.Name. NormD1 | 3.483E-09 | D1 |
| | Visit.Name. NormD3 | -1.572422 | 0.192995 | 88 | -8.147 | 2.32E-12 | Visit.Name. NormD3 | 1.621E-11 | D3 |
| | Visit.Name. NormD7 | -1.175069 | 0.187229 | 88 | -6.276 | 1.27E-08 | Visit.Name. NormD7 | 2.968E-08 | D7 |
| | Visit.Name. NormD14 | -0.919434 | 0.198391 | 88 | -4.634 | 1.23E-05 | Visit.Name. NormD14 | 2.158E-05 | D14 |
| | Visit.Name. NormD21 | -0.560295 | 0.199138 | 88 | -2.814 | 6.04E-03 | Visit.Name. NormD21 | 8.459E-03 | D21 |
| | Visit.Name. NormD30 | -0.558978 | 0.206990 | 88 | -2.701 | 8.30E-03 | Visit.Name. NormD30 | 9.687E-03 | D30 |
| | Visit.Name. NormD56 | -0.028193 | 0.358232 | 88 | -0.079 | 9.37E-01 | Visit.Name. NormD56 | 9.374E-01 | D56 |
| MSS-CRC | Visit.Name. NormD1 | -1.077304 | 0.243794 | 68 | -4.419 | 3.66E-05 | Visit.Name. NormD1 | 1.279E-04 | D1 |
| | Visit.Name. NormD3 | -1.027645 | 0.223202 | 68 | -4.604 | 1.87E-05 | Visit.Name. NormD3 | 1.279E-04 | D3 |
| | Visit.Name. NormD7 | -0.786646 | 0.228848 | 68 | -3.437 | 1.01E-03 | Visit.Name. NormD7 | 2.348E-03 | D7 |
| | Visit.Name. NormD14 | -0.359514 | 0.213467 | 68 | -1.684 | 9.67E-02 | Visit.Name. NormD14 | 1.354E-01 | D14 |
| | Visit.Name. NormD21 | -0.028928 | 0.222973 | 68 | -0.130 | 8.97E-01 | Visit.Name. NormD21 | 8.972E-01 | D21 |
| | Visit.Name. NormD30 | -0.210266 | 0.223109 | 68 | -0.942 | 3.49E-01 | Visit.Name. NormD30 | 4.075E-01 | D30 |
| | Visit.Name. NormD56 | -0.492029 | 0.280279 | 68 | -1.755 | 8.37E-02 | Visit.Name. NormD56 | 1.354E-01 | D56 |

**Supplementary Table S12: Shannon Diversity of Melanoma Cohort Stratified by Clinical Activity and VE800 Engraftment**

| CLINICAL ACTIVITY STRATIFICATION | | | | | | |
|---|---|---|---|---|---|---|
| | **Value** | **Std.Error** | **DF** | **t.value** | **p.value** | **Var** |
| (Intercept) | 3.41274 | 0.17844 | 100 | 19.12509 | 0.0000 | (Intercept) |
| Visit.Name.NormD1 | -0.91307 | 0.21073 | 100 | -4.33288 | 0.0000 | Visit.Name. NormD1 |
| Visit.Name.NormD3 | -1.03101 | 0.21073 | 100 | -4.89252 | 0.0000 | Visit.Name. NormD3 |
| Visit.Name.NormD7 | -0.93584 | 0.21606 | 100 | -4.33150 | 0.0000 | Visit.Name. NormD7 |
| Visit.Name.NormD14 | -0.73314 | 0.22240 | 100 | -3.29644 | 0.0014 | Visit.Name. NormD14 |
| Visit.Name.NormD21 | -0.37108 | 0.22249 | 100 | -1.66782 | 0.0985 | Visit.Name. NormD21 |
| Visit.Name.NormD30 | -0.18004 | 0.22982 | 100 | -0.78342 | 0.4352 | Visit.Name. NormD30 |
| Visit.Name.NormD58 | -0.39727 | 0.34481 | 100 | -1.15213 | 0.2520 | Visit.Name. NormD58 |
| Response_Wk8SD | 0.63748 | 0.29200 | 18 | 2.18316 | 0.0425 | Response_Wk8 SD |
| Visit.Name.NormD1: Response_Wk8SD | -0.08153 | 0.34718 | 100 | -0.23484 | 0.8148 | Visit.Name. NormD1: Response_Wk8 SD |
| Visit.Name.NormD3: Response_Wk8SD | -0.61455 | 0.35761 | 100 | -1.71851 | 0.0888 | Visit.Name. NormD3: Response_Wk8 SD |
| Visit.Name.NormD7: Response_Wk8SD | -0.61531 | 0.36077 | 100 | -1.70555 | 0.0912 | Visit.Name. NormD7: Response_Wk8 SD |
| Visit.Name.NormD14: Response_Wk8SD | -0.58787 | 0.35714 | 100 | -1.64603 | 0.1029 | Visit.Name. NormD14: Response_Wk8 SD |
| Visit.Name.NormD21: Response_Wk8SD | -0.66108 | 0.36466 | 100 | -1.81286 | 0.0729 | Visit.Name. NormD21: Response_Wk8 SD |
| Visit.Name.NormD30: Response_Wk8SD | -0.57489 | 0.35368 | 100 | -1.62546 | 0.1072 | Visit.Name. NormD30: Response_Wk8 SD |
| Visit.Name.NormD58: Response_Wk8SD | -0.26358 | 0.43723 | 100 | -0.60284 | 0.5480 | Visit.Name. NormD58: Response_Wk8 SD |

| VE800 ENGRAFTMENT STRATIFICATION | | | | | | |
|---|---|---|---|---|---|---|
| | **Value** | **Std.Error** | **DF** | **t.value** | **p.value** | **Var** |
| (Intercept) | 4.14057 | 0.18862 | 100 | 21.95203 | 0.0000 | (Intercept) |
| Visit.Name.NormD1 | -0.90895 | 0.23410 | 100 | -3.88274 | 0.0002 | Visit.Name. NormD1 |
| Visit.Name.NormD3 | -1.58713 | 0.24229 | 100 | -6.55043 | 0.0000 | Visit.Name. NormD3 |
| Visit.Name.NormD7 | -1.62489 | 0.24229 | 100 | -6.70626 | 0.0000 | Visit.Name. NormD7 |
| Visit.Name.NormD14 | -1.39101 | 0.23610 | 100 | -5.89169 | 0.0000 | Visit.Name. NormD14 |
| Visit.Name.NormD21 | -0.90016 | 0.24229 | 100 | -3.71516 | 0.0003 | Visit.Name. NormD21 |
| Visit.Name.NormD30 | -0.74101 | 0.22915 | 100 | -3.23376 | 0.0017 | Visit.Name. NormD30 |
| Visit.Name.NormD58 | -0.74701 | 0.24402 | 100 | -3.06125 | 0.0028 | Visit.Name. NormD58 |

| | | | | | | |
|---|---|---|---|---|---|---|
| EngraftmentLC | -0.93837 | 0.26071 | 18 | -3.59930 | 0.0021 | EngraftmentLC |
| Visit.Name.NormD1: EngraftmentLC | -0.06490 | 0.32269 | 100 | -0.20114 | 0.8410 | Visit.Name. NormD1: EngraftmentLC |
| Visit.Name.NormD3: EngraftmentLC | 0.63129 | 0.32868 | 100 | 1.92071 | 0.0576 | Visit.Name. NormD3: EngraftmentLC |
| Visit.Name.NormD7: EngraftmentLC | 0.87518 | 0.33329 | 100 | 2.62592 | 0.0100 | Visit.Name. NormD7: EngraftmentLC |
| Visit.Name.NormD14: EngraftmentLC | 0.84457 | 0.33465 | 100 | 2.52376 | 0.0132 | Visit.Name. NormD14: EngraftmentLC |
| Visit.Name.NormD21: EngraftmentLC | 0.53126 | 0.33911 | 100 | 1.56661 | 0.1204 | Visit.Name. NormD21: EngraftmentLC |
| Visit.Name.NormD30: EngraftmentLC | 0.64144 | 0.33716 | 100 | 1.90250 | 0.0600 | Visit.Name. NormD30: EngraftmentLC |
| Visit.Name.NormD58: EngraftmentLC | 0.64151 | 0.41548 | 100 | 1.54402 | 0.1257 | Visit.Name. NormD58: EngraftmentLC |

#### Supplementary Table S13: Endogenous Microbiome Species In All Cohorts

| | Value | Std.Error | DF | t.value | pval | Bac | Var | p_adj |
|---|---|---|---|---|---|---|---|---|
| 1 | 0.49615 | 0.08072 | 68 | 6.14638 | 0.00000 | Bacteroides.sp..VE800.11 | Visit.Name.NormD58 | 0.00001 |
| 2 | 0.37025 | 0.06416 | 68 | 5.77079 | 0.00000 | Bacteroides.sp..VE800.11 | Visit.Name.NormD30 | 0.00003 |
| 3 | 0.34151 | 0.06411 | 68 | 5.32664 | 0.00000 | Bacteroides.sp..VE800.11 | Visit.Name.NormD21 | 0.00016 |
| 4 | 0.19967 | 0.03913 | 68 | 5.10257 | 0.00000 | Parabacteroides.sp..VE800.07 | Visit.Name.NormD7 | 0.00038 |
| 5 | 0.09135 | 0.01863 | 68 | 4.90392 | 0.00001 | Phascolarctobacterium.sp..VE800.04 | Visit.Name.NormD7 | 0.00040 |
| 6 | 0.18440 | 0.03643 | 68 | 5.06119 | 0.00000 | Parabacteroides.sp..VE800.07 | Visit.Name.NormD14 | 0.00044 |
| 7 | 0.29610 | 0.06135 | 68 | 4.82636 | 0.00001 | Bacteroides.sp..VE800.11 | Visit.Name.NormD14 | 0.00054 |
| 8 | -0.09920 | 0.02108 | 68 | -4.70641 | 0.00001 | Faecalibacterium.prausnitzii | Visit.Name.NormD14 | 0.00056 |
| 9 | 0.06403 | 0.01357 | 68 | 4.71747 | 0.00001 | Eubacterium.sp..VE800.02 | Visit.Name.NormD21 | 0.00081 |
| 10 | 0.13102 | 0.02935 | 68 | 4.46461 | 0.00003 | Parabacteroides.sp..VE800.10 | Visit.Name.NormD14 | 0.00102 |
| 11 | -0.10187 | 0.02258 | 68 | -4.51258 | 0.00003 | Faecalibacterium.prausnitzii | Visit.Name.NormD7 | 0.00114 |
| 12 | 0.20743 | 0.04486 | 68 | 4.62355 | 0.00002 | Bacteroides.sp..VE800.09 | Visit.Name.NormD30 | 0.00114 |
| 13 | -0.06299 | 0.01438 | 68 | -4.38119 | 0.00004 | Blautia.sp..MP512E3 | Visit.Name.NormD7 | 0.00137 |
| 14 | 0.15838 | 0.03911 | 68 | 4.05002 | 0.00013 | Fusobacterium.sp..VE800.01 | Visit.Name.NormD7 | 0.00350 |
| 15 | 0.06804 | 0.01734 | 68 | 3.92449 | 0.00021 | Phascolarctobacterium.sp..VE800.04 | Visit.Name.NormD14 | 0.00538 |
| 16 | 0.05005 | 0.01298 | 68 | 3.85581 | 0.00026 | Eubacterium.sp..VE800.02 | Visit.Name.NormD14 | 0.00565 |
| 17 | -0.04089 | 0.01089 | 68 | -3.75348 | 0.00036 | Blautia.sp..MP341G7 | Visit.Name.NormD14 | 0.00595 |
| 18 | -0.04804 | 0.01269 | 68 | -3.78536 | 0.00033 | X.Eubacterium..hallii | Visit.Name.NormD14 | 0.00595 |
| 19 | -0.08880 | 0.02201 | 68 | -4.03369 | 0.00014 | Faecalibacterium.prausnitzii | Visit.Name.NormD30 | 0.00618 |
| 20 | 0.19135 | 0.04766 | 68 | 4.01460 | 0.00015 | Lactobacillus.rhamnosus | Visit.Name.NormD3 | 0.00848 |
| 21 | -0.08678 | 0.02202 | 68 | -3.94075 | 0.00019 | Faecalibacterium.prausnitzii | Visit.Name.NormD3 | 0.00848 |
| 22 | -0.05893 | 0.01402 | 68 | -4.20187 | 0.00008 | Blautia.sp..MP512E3 | Visit.Name.NormD3 | 0.00848 |
| 23 | -0.04288 | 0.01132 | 68 | -3.78728 | 0.00032 | Blautia.sp..MP341G7 | Visit.Name.NormD21 | 0.00883 |
| 24 | -0.08309 | 0.02200 | 68 | -3.77628 | 0.00034 | Faecalibacterium.prausnitzii | Visit.Name.NormD21 | 0.00883 |
| 25 | -0.05013 | 0.01325 | 68 | -3.78466 | 0.00033 | X.Eubacterium..hallii | Visit.Name.NormD21 | 0.00883 |
| 26 | -0.04227 | 0.01132 | 68 | -3.73376 | 0.00039 | Blautia.sp..MP341G7 | Visit.Name.NormD30 | 0.01270 |
| 27 | -0.04126 | 0.01159 | 68 | -3.56110 | 0.00068 | Blautia.sp..MP341G7 | Visit.Name.NormD7 | 0.01272 |
| 28 | -0.04127 | 0.01154 | 68 | -3.57539 | 0.00065 | Blautia.sp..MP78G6 | Visit.Name.NormD7 | 0.01272 |
| 29 | 0.04953 | 0.01359 | 68 | 3.64569 | 0.00052 | Eubacterium.sp..VE800.02 | Visit.Name.NormD30 | 0.01355 |
| 30 | 0.13944 | 0.03814 | 68 | 3.65603 | 0.00050 | Fusobacterium.sp..VE800.01 | Visit.Name.NormD3 | 0.01638 |
| 31 | 0.04815 | 0.01438 | 68 | 3.34867 | 0.00133 | Bifidobacterium.dentium | Visit.Name.NormD14 | 0.01738 |
| 32 | -0.04493 | 0.01342 | 68 | -3.34894 | 0.00133 | Blautia.sp..MP512E3 | Visit.Name.NormD14 | 0.01738 |
| 33 | 0.07081 | 0.02167 | 68 | 3.26763 | 0.00170 | Ruthenibacterium.sp..VE800.03 | Visit.Name.NormD14 | 0.01756 |
| 34 | -0.04362 | 0.01338 | 68 | -3.25986 | 0.00174 | Blautia.sp..MP40F7 | Visit.Name.NormD14 | 0.01756 |
| 35 | 0.14023 | 0.04288 | 68 | 3.26999 | 0.00169 | Bacteroides.sp..VE800.09 | Visit.Name.NormD14 | 0.01756 |
| 36 | -0.03483 | 0.01079 | 68 | -3.22776 | 0.00192 | Blautia.sp..MP78G6 | Visit.Name.NormD14 | 0.01798 |
| 37 | -0.04311 | 0.01267 | 68 | -3.40312 | 0.00112 | Eggerthella.lenta | Visit.Name.NormD7 | 0.01835 |
| 38 | 0.07716 | 0.02264 | 68 | 3.40785 | 0.00110 | Ruthenibacterium.sp..VE800.03 | Visit.Name.NormD21 | 0.01958 |
| 39 | 0.03852 | 0.01139 | 68 | 3.38233 | 0.00120 | Dorea.sp..MP50E3 | Visit.Name.NormD21 | 0.01958 |
| 40 | 0.15561 | 0.04483 | 68 | 3.47132 | 0.00090 | Bacteroides.sp..VE800.09 | Visit.Name.NormD21 | 0.01958 |
| 41 | 0.10090 | 0.03059 | 68 | 3.29795 | 0.00155 | Parabacteroides.sp..VE800.10 | Visit.Name.NormD21 | 0.02032 |
| 42 | 0.12683 | 0.03810 | 68 | 3.32897 | 0.00141 | Parabacteroides.sp..VE800.07 | Visit.Name.NormD21 | 0.02032 |
| 43 | 0.05312 | 0.01560 | 68 | 3.40443 | 0.00112 | Bilophila.wadsworthia | Visit.Name.NormD3 | 0.02462 |
| 44 | 0.03789 | 0.01114 | 68 | 3.40109 | 0.00113 | Veillonella.sp..MP19D1_02.09 | Visit.Name.NormD3 | 0.02462 |
| 45 | -0.06807 | 0.02100 | 68 | -3.24220 | 0.00184 | Absiella.sp..MP391A4 | Visit.Name.NormD7 | 0.02677 |
| 46 | 0.05735 | 0.01813 | 68 | 3.16252 | 0.00234 | Phascolarctobacterium.sp..VE800.04 | Visit.Name.NormD21 | 0.02784 |
| 47 | 0.09965 | 0.03136 | 68 | 3.17710 | 0.00224 | Parabacteroides.sp..VE800.10 | Visit.Name.NormD7 | 0.02932 |
| 48 | -0.06716 | 0.02050 | 68 | -3.27679 | 0.00166 | Absiella.sp..MP391A4 | Visit.Name.NormD3 | 0.03098 |
| 49 | -0.04342 | 0.01473 | 68 | -2.94807 | 0.00438 | Blautia.sp..MP237F5b | Visit.Name.NormD14 | 0.03585 |
| 50 | -0.05817 | 0.01968 | 68 | -2.95541 | 0.00429 | Absiella.sp..MP391A4 | Visit.Name.NormD14 | 0.03585 |
| 51 | -0.04442 | 0.01396 | 68 | -3.18269 | 0.00220 | Blautia.sp..MP40F7 | Visit.Name.NormD3 | 0.03604 |
| 52 | -0.04304 | 0.01325 | 68 | -3.24778 | 0.00181 | X.Eubacterium..hallii | Visit.Name.NormD30 | 0.03948 |
| 53 | -0.03451 | 0.01135 | 68 | -3.04105 | 0.00335 | Ruthenibacterium.lactatiformans | Visit.Name.NormD7 | 0.03962 |
| 54 | 0.05274 | 0.01767 | 68 | 2.98552 | 0.00393 | Colibacter.massiliensis | Visit.Name.NormD7 | 0.03962 |
| 55 | -0.09841 | 0.03274 | 68 | -3.00585 | 0.00371 | Lachnospiraceae.bacterium.1_1_57FAA | Visit.Name.NormD7 | 0.03962 |
| 56 | -0.05299 | 0.01857 | 68 | -2.85388 | 0.00572 | Blautia.sp..MP588D7 | Visit.Name.NormD7 | 0.05351 |
| 57 | -0.09604 | 0.03194 | 68 | -3.00676 | 0.00370 | Lachnospiraceae.bacterium.1_1_57FAA | Visit.Name.NormD3 | 0.05382 |
| 58 | 0.11783 | 0.03813 | 68 | 3.08982 | 0.00290 | Parabacteroides.sp..VE800.07 | Visit.Name.NormD30 | 0.05429 |
| 59 | -0.03256 | 0.01132 | 68 | -2.87616 | 0.00537 | Blautia.sp..MP341G7 | Visit.Name.NormD3 | 0.05540 |
| 60 | -0.03734 | 0.01326 | 68 | -2.81681 | 0.00634 | X.Eubacterium..hallii | Visit.Name.NormD3 | 0.05540 |
| 61 | -0.04224 | 0.01482 | 68 | -2.85037 | 0.00578 | Roseburia.intestinalis | Visit.Name.NormD3 | 0.05540 |
| 62 | -0.05128 | 0.01811 | 68 | -2.83117 | 0.00609 | Blautia.sp..MP588D7 | Visit.Name.NormD3 | 0.05540 |
| 63 | -0.04345 | 0.01531 | 68 | -2.83790 | 0.00598 | Blautia.sp..MP237F5b | Visit.Name.NormD3 | 0.05540 |
| 64 | -0.03145 | 0.01107 | 68 | -2.84110 | 0.00593 | Ruthenibacterium.lactatiformans | Visit.Name.NormD3 | 0.05540 |
| 65 | 0.18265 | 0.06582 | 68 | 2.77488 | 0.00712 | Bacteroides.sp..VE800.11 | Visit.Name.NormD7 | 0.05551 |
| 66 | -0.03076 | 0.01110 | 68 | -2.77083 | 0.00720 | Oscillospiraceae.bacterium.VE202.24 | Visit.Name.NormD7 | 0.05551 |
| 67 | -0.04347 | 0.01567 | 68 | -2.77387 | 0.00714 | Blautia.sp..MP237F5b | Visit.Name.NormD7 | 0.05551 |
| 68 | -0.03127 | 0.01126 | 68 | -2.77691 | 0.00708 | Blautia.sp..MP78G6 | Visit.Name.NormD3 | 0.05800 |
| 69 | -0.03249 | 0.01184 | 68 | -2.74530 | 0.00773 | Eggerthella.lenta | Visit.Name.NormD14 | 0.05955 |
| 70 | 0.01162 | 0.00424 | 68 | 2.73904 | 0.00786 | Phascolarctobacterium.succinatutens | Visit.Name.NormD3 | 0.06057 |
| 71 | -0.03422 | 0.01265 | 68 | -2.70510 | 0.00862 | Alistipes.sp..MP237E6a | Visit.Name.NormD3 | 0.06275 |
| 72 | -0.02876 | 0.01083 | 68 | -2.65668 | 0.00982 | Oscillospiraceae.bacterium.VE202.24 | Visit.Name.NormD3 | 0.06358 |
| 73 | -0.03414 | 0.01287 | 68 | -2.65245 | 0.00994 | Tyzzerella.sp..MP10F1re | Visit.Name.NormD3 | 0.06358 |

| | | | | | | | | |
|---|---|---|---|---|---|---|---|---|
| 74 | 0.06380 | 0.02414 | 68 | 2.64297 | 0.01019 | Citrobacter.freundii | Visit.Name.NormD3 | 0.06358 |
| 75 | 0.04222 | 0.01600 | 68 | 2.63842 | 0.01032 | Bilophila.wadsworthia | Visit.Name.NormD7 | 0.06436 |
| 76 | -0.03486 | 0.01295 | 68 | -2.69137 | 0.00895 | HI22.sp..HI22 | Visit.Name.NormD7 | 0.06436 |
| 77 | -0.03457 | 0.01296 | 68 | -2.66676 | 0.00956 | Alistipes.sp..MP237E6a | Visit.Name.NormD7 | 0.06436 |
| 78 | 0.10317 | 0.03892 | 68 | 2.65050 | 0.00999 | Akkermansia.muciniphila | Visit.Name.NormD7 | 0.06436 |
| 79 | -0.08797 | 0.02403 | 68 | -3.66052 | 0.00049 | Faecalibacterium.prausnitzii | Visit.Name.NormD1 | 0.06457 |
| 80 | -0.04345 | 0.01531 | 68 | -2.83805 | 0.00598 | Blautia.sp..MP237F5b | Visit.Name.NormD21 | 0.06526 |
| 81 | -0.03447 | 0.01319 | 68 | -2.61293 | 0.01104 | Tyzzerella.sp..MP10F1re | Visit.Name.NormD7 | 0.06574 |
| 82 | -0.04218 | 0.01615 | 68 | -2.61265 | 0.01105 | Blautia.sp..MP245E9 | Visit.Name.NormD3 | 0.06579 |
| 83 | -0.03694 | 0.01430 | 68 | -2.58340 | 0.01194 | Blautia.sp..MP40F7 | Visit.Name.NormD7 | 0.06799 |
| 84 | -0.04218 | 0.01652 | 68 | -2.55301 | 0.01293 | Blautia.sp..MP245E9 | Visit.Name.NormD7 | 0.07007 |
| 85 | 0.02967 | 0.01168 | 68 | 2.54009 | 0.01337 | Pseudomonas.aeruginosa | Visit.Name.NormD7 | 0.07007 |
| 86 | 0.03795 | 0.01531 | 68 | 2.47819 | 0.01569 | Bifidobacterium.dentium | Visit.Name.NormD7 | 0.07907 |
| 87 | -0.03456 | 0.01263 | 68 | -2.73540 | 0.00794 | HI22.sp..HI22 | Visit.Name.NormD21 | 0.08000 |
| 88 | 0.19004 | 0.05650 | 68 | 3.36367 | 0.00127 | Bacteroides.sp..VE800.09 | Visit.Name.NormD58 | 0.08298 |
| 89 | 0.17447 | 0.05205 | 68 | 3.35184 | 0.00131 | Lactobacillus.rhamnosus | Visit.Name.NormD1 | 0.08608 |
| 90 | 0.03271 | 0.01345 | 68 | 2.43278 | 0.01762 | Klebsiella.oxytoca | Visit.Name.NormD3 | 0.09703 |
| 91 | 0.04347 | 0.01790 | 68 | 2.42928 | 0.01778 | Veillonella.atypica | Visit.Name.NormD3 | 0.09703 |
| 92 | -0.04344 | 0.01531 | 68 | -2.83729 | 0.00599 | Blautia.sp..MP237F5b | Visit.Name.NormD30 | 0.09810 |
| 93 | 0.03305 | 0.01394 | 68 | 2.37049 | 0.02061 | Eubacterium.sp..VE800.02 | Visit.Name.NormD7 | 0.09998 |
| 94 | 0.09203 | 0.03646 | 68 | 2.52408 | 0.01394 | Fusobacterium.sp..VE800.01 | Visit.Name.NormD14 | 0.10145 |
| 95 | 0.03221 | 0.01378 | 68 | 2.33805 | 0.02233 | Klebsiella.oxytoca | Visit.Name.NormD7 | 0.10449 |
| 96 | -0.03762 | 0.01226 | 68 | -3.06746 | 0.00310 | Blautia.sp..MP341G7 | Visit.Name.NormD1 | 0.10497 |
| 97 | -0.04718 | 0.01618 | 68 | -2.91534 | 0.00481 | Roseburia.intestinalis | Visit.Name.NormD1 | 0.10497 |
| 98 | -0.03596 | 0.01209 | 68 | -2.97442 | 0.00406 | Ruthenibacterium.lactatiformans | Visit.Name.NormD1 | 0.10497 |
| 99 | 0.20039 | 0.06615 | 68 | 3.02927 | 0.00346 | Klebsiella.michiganensis | Visit.Name.NormD1 | 0.10497 |
| 100 | -0.03528 | 0.01417 | 68 | -2.48894 | 0.01527 | Roseburia.intestinalis | Visit.Name.NormD14 | 0.10525 |
| 101 | -0.03808 | 0.01553 | 68 | -2.45139 | 0.01681 | Blautia.sp..MP245E9 | Visit.Name.NormD14 | 0.11008 |
| 102 | 0.04945 | 0.01815 | 68 | 2.72424 | 0.00818 | Phascolarctobacterium.sp..VE800.04 | Visit.Name.NormD30 | 0.11383 |
| 103 | 0.07998 | 0.03060 | 68 | 2.61369 | 0.01102 | Parabacteroides.sp..VE800.10 | Visit.Name.NormD30 | 0.11383 |
| 104 | 0.02979 | 0.01139 | 68 | 2.61544 | 0.01097 | Dorea.sp..MP50E3 | Visit.Name.NormD30 | 0.11383 |
| 105 | -0.03340 | 0.01265 | 68 | -2.64039 | 0.01026 | Alistipes.sp..MP237E6a | Visit.Name.NormD30 | 0.11383 |
| 106 | -0.05338 | 0.02050 | 68 | -2.60430 | 0.01130 | Absiella.sp..MP391A4 | Visit.Name.NormD30 | 0.11383 |
| 107 | -0.02758 | 0.01206 | 68 | -2.28749 | 0.02528 | Alistipes.putredinis | Visit.Name.NormD7 | 0.11421 |
| 108 | -0.04333 | 0.01531 | 68 | -2.82986 | 0.00612 | Blautia.sp..MP512E3 | Visit.Name.NormD1 | 0.11447 |
| 109 | 0.02390 | 0.01023 | 68 | 2.33656 | 0.02242 | Streptococcus.thermophilus | Visit.Name.NormD3 | 0.11746 |
| 110 | 0.04088 | 0.01769 | 68 | 2.31107 | 0.02387 | Veillonella.parvula | Visit.Name.NormD3 | 0.12025 |
| 111 | 0.03311 | 0.01445 | 68 | 2.29139 | 0.02504 | X.Clostridium..symbiosum | Visit.Name.NormD3 | 0.12151 |
| 112 | -0.02725 | 0.01236 | 68 | -2.20540 | 0.03081 | Eggerthella.lenta | Visit.Name.NormD3 | 0.13088 |
| 113 | 0.02591 | 0.01178 | 68 | 2.19954 | 0.03124 | Parasutterella.excrementihominis | Visit.Name.NormD3 | 0.13088 |
| 114 | -0.02823 | 0.01264 | 68 | -2.23361 | 0.02880 | HI22.sp..HI22 | Visit.Name.NormD3 | 0.13088 |
| 115 | 0.01915 | 0.00878 | 68 | 2.18114 | 0.03263 | Megasphaera.sp..MP182G6 | Visit.Name.NormD3 | 0.13088 |
| 116 | -0.03271 | 0.01470 | 68 | -2.22580 | 0.02935 | Blautia.sp..MP87G3 | Visit.Name.NormD3 | 0.13088 |
| 117 | 0.05010 | 0.02302 | 68 | 2.17682 | 0.03297 | Lactobacillus.salivarius | Visit.Name.NormD3 | 0.13088 |
| 118 | -0.09496 | 0.03481 | 68 | -2.72783 | 0.00810 | Lachnospiraceae.bacterium.1_1_57FAA | Visit.Name.NormD1 | 0.13272 |
| 119 | 0.03941 | 0.01693 | 68 | 2.32873 | 0.02285 | Parabacteroides.sp..VE800.05 | Visit.Name.NormD14 | 0.13375 |
| 120 | -0.02462 | 0.01059 | 68 | -2.32518 | 0.02305 | Ruthenibacterium.lactatiformans | Visit.Name.NormD14 | 0.13375 |
| 121 | -0.07094 | 0.03061 | 68 | -2.31768 | 0.02348 | Lachnospiraceae.bacterium.1_1_57FAA | Visit.Name.NormD14 | 0.13375 |
| 122 | 0.02754 | 0.01100 | 68 | 2.50365 | 0.01470 | X.Clostridium..leptum | Visit.Name.NormD21 | 0.13753 |
| 123 | 0.03888 | 0.01817 | 68 | 2.14052 | 0.03590 | Phascolarctobacterium.sp..VE800.04 | Visit.Name.NormD3 | 0.13833 |
| 124 | -0.02975 | 0.01359 | 68 | -2.18920 | 0.03202 | X.Eubacterium..hallii | Visit.Name.NormD7 | 0.13981 |
| 125 | 0.05876 | 0.02724 | 68 | 2.15735 | 0.03452 | Raoultella.ornithinolytica | Visit.Name.NormD7 | 0.14034 |
| 126 | -0.03134 | 0.01449 | 68 | -2.16223 | 0.03412 | Bacteroides.dorei | Visit.Name.NormD7 | 0.14034 |
| 127 | 0.05309 | 0.02473 | 68 | 2.14713 | 0.03535 | Citrobacter.freundii | Visit.Name.NormD7 | 0.14034 |
| 128 | 0.03544 | 0.01674 | 68 | 2.11718 | 0.03790 | Sutterella.wadsworthensis | Visit.Name.NormD3 | 0.14187 |
| 129 | 0.03142 | 0.01212 | 68 | 2.59289 | 0.01164 | Veillonella.sp..MP19D1_02.09 | Visit.Name.NormD1 | 0.15251 |
| 130 | -0.04328 | 0.01659 | 68 | -2.60856 | 0.01117 | Blautia.sp..MP237F5b | Visit.Name.NormD1 | 0.15251 |
| 131 | 0.15333 | 0.06169 | 68 | 2.48536 | 0.01541 | Escherichia.coli | Visit.Name.NormD1 | 0.15296 |
| 132 | -0.03522 | 0.01446 | 68 | -2.43512 | 0.01751 | X.Eubacterium..hallii | Visit.Name.NormD1 | 0.15296 |
| 133 | -0.03379 | 0.01377 | 68 | -2.45411 | 0.01669 | HI22.sp..HI22 | Visit.Name.NormD1 | 0.15296 |
| 134 | -0.03480 | 0.01377 | 68 | -2.52744 | 0.01382 | Alistipes.sp..MP237E6a | Visit.Name.NormD1 | 0.15296 |
| 135 | -0.03520 | 0.01402 | 68 | -2.51099 | 0.01442 | Tyzzerella.sp..MP10F1re | Visit.Name.NormD1 | 0.15296 |
| 136 | -0.02332 | 0.01126 | 68 | -2.07162 | 0.04210 | Roseburia.faecis | Visit.Name.NormD3 | 0.15319 |
| 137 | 0.15920 | 0.06611 | 68 | 2.40813 | 0.01875 | Klebsiella.pneumoniae | Visit.Name.NormD1 | 0.15353 |
| 138 | 0.02329 | 0.01111 | 68 | 2.09549 | 0.03985 | Desulfovibrio.fairfieldensis | Visit.Name.NormD7 | 0.15355 |
| 139 | -0.07321 | 0.03561 | 68 | -2.05625 | 0.04360 | Bacteroides.thetaiotaomicron | Visit.Name.NormD3 | 0.15436 |
| 140 | -0.03152 | 0.01519 | 68 | -2.07443 | 0.04183 | Roseburia.intestinalis | Visit.Name.NormD7 | 0.15656 |
| 141 | 0.03967 | 0.01924 | 68 | 2.06208 | 0.04302 | Alistipes.sp..VE800.08 | Visit.Name.NormD7 | 0.15656 |
| 142 | -0.05004 | 0.02484 | 68 | -2.01403 | 0.04797 | Alistipes.finegoldii | Visit.Name.NormD3 | 0.16119 |
| 143 | -0.02846 | 0.01413 | 68 | -2.01382 | 0.04799 | Bacteroides.dorei | Visit.Name.NormD3 | 0.16119 |
| 144 | 0.05442 | 0.02266 | 68 | 2.40193 | 0.01905 | Ruthenibacterium.sp..VE800.03 | Visit.Name.NormD30 | 0.16182 |
| 145 | 0.02695 | 0.01126 | 68 | 2.39390 | 0.01943 | Blautia.sp..VE303.05 | Visit.Name.NormD30 | 0.16182 |
| 146 | 0.03161 | 0.01324 | 68 | 2.38718 | 0.01976 | Collinsella.tanakaei | Visit.Name.NormD30 | 0.16182 |
| 147 | -0.02899 | 0.01227 | 68 | -2.36227 | 0.02103 | Blautia.sp..MP78G6 | Visit.Name.NormD1 | 0.16207 |
| 148 | -0.02347 | 0.01154 | 68 | -2.03443 | 0.04581 | Roseburia.faecis | Visit.Name.NormD7 | 0.16219 |

| | | | | | | | | |
|---|---|---|---|---|---|---|---|---|
| 149 | 0.12066 | 0.06056 | 68 | 1.99226 | 0.05036 | Klebsiella.michiganensis | Visit.Name.NormD3 | 0.16493 |
| 150 | -0.02328 | 0.01176 | 68 | -1.97971 | 0.05179 | Alistipes.putredinis | Visit.Name.NormD3 | 0.16546 |
| 151 | 0.01755 | 0.00756 | 68 | 2.32189 | 0.02324 | Megasphaera.sp..UBA1696 | Visit.Name.NormD1 | 0.16915 |
| 152 | 0.02765 | 0.01265 | 68 | 2.18502 | 0.03234 | Blautia.coccoides | Visit.Name.NormD14 | 0.17382 |
| 153 | 0.00878 | 0.00404 | 68 | 2.17423 | 0.03317 | Phascolarctobacterium.succinatutens | Visit.Name.NormD14 | 0.17382 |
| 154 | -0.03420 | 0.01764 | 68 | -1.93925 | 0.05662 | Solobacterium.moorei | Visit.Name.NormD3 | 0.17660 |
| 155 | -0.01774 | 0.00928 | 68 | -1.91105 | 0.06021 | X.Clostridium..aldenense | Visit.Name.NormD3 | 0.17938 |
| 156 | -0.03351 | 0.01755 | 68 | -1.90905 | 0.06048 | Bacteroides.xylanisolvens | Visit.Name.NormD3 | 0.17938 |
| 157 | -0.04173 | 0.02196 | 68 | -1.90042 | 0.06162 | Bacteroides.vulgatus | Visit.Name.NormD3 | 0.17938 |
| 158 | -0.03826 | 0.01936 | 68 | -1.97631 | 0.05218 | Alistipes.sp..HK66 | Visit.Name.NormD7 | 0.17988 |
| 159 | -0.03952 | 0.01749 | 68 | -2.25991 | 0.02703 | Blautia.sp..MP245E9 | Visit.Name.NormD1 | 0.18639 |
| 160 | -0.03483 | 0.01800 | 68 | -1.93536 | 0.05711 | Bacteroides.xylanisolvens | Visit.Name.NormD7 | 0.18764 |
| 161 | -0.02912 | 0.01506 | 68 | -1.93385 | 0.05729 | Blautia.sp..MP87G3 | Visit.Name.NormD7 | 0.18764 |
| 162 | -0.03388 | 0.01481 | 68 | -2.28730 | 0.02530 | Roseburia.intestinalis | Visit.Name.NormD30 | 0.19234 |
| 163 | -0.02921 | 0.01287 | 68 | -2.26927 | 0.02643 | Tyzzerella.sp..MP10F1re | Visit.Name.NormD30 | 0.19234 |
| 164 | -0.04293 | 0.02247 | 68 | -1.91036 | 0.06031 | Bacteroides.vulgatus | Visit.Name.NormD7 | 0.19268 |
| 165 | -0.02979 | 0.01348 | 68 | -2.21095 | 0.03041 | Eggerthella.lenta | Visit.Name.NormD1 | 0.19916 |
| 166 | -0.03379 | 0.01808 | 68 | -1.86862 | 0.06598 | Solobacterium.moorei | Visit.Name.NormD7 | 0.20581 |
| 167 | 0.01251 | 0.00689 | 68 | 1.81414 | 0.07407 | Megasphaera.sp..UBA1696 | Visit.Name.NormD3 | 0.20644 |
| 168 | -0.03484 | 0.01914 | 68 | -1.82031 | 0.07311 | Bacteroides.sp..MP264A7 | Visit.Name.NormD3 | 0.20644 |
| 169 | -0.03492 | 0.01683 | 68 | -2.07433 | 0.04184 | Solobacterium.moorei | Visit.Name.NormD14 | 0.21079 |
| 170 | 0.02397 | 0.01309 | 68 | 1.83144 | 0.07142 | Prevotella.copri | Visit.Name.NormD7 | 0.21757 |
| 171 | 0.08812 | 0.04887 | 68 | 1.80313 | 0.07580 | Lactobacillus.rhamnosus | Visit.Name.NormD7 | 0.22206 |
| 172 | -0.04087 | 0.02271 | 68 | -1.80011 | 0.07628 | Eubacterium.sp..CAG.180 | Visit.Name.NormD7 | 0.22206 |
| 173 | -0.03518 | 0.01733 | 68 | -2.02942 | 0.04633 | Blautia.sp..MP588D7 | Visit.Name.NormD14 | 0.22479 |
| 174 | -0.02678 | 0.01235 | 68 | -2.16800 | 0.03366 | Eggerthella.lenta | Visit.Name.NormD30 | 0.23209 |
| 175 | -0.02684 | 0.01519 | 68 | -1.76677 | 0.08175 | Blautia.sp..HF17 | Visit.Name.NormD7 | 0.23282 |
| 176 | 0.02750 | 0.01332 | 68 | 2.06418 | 0.04282 | Methanobrevibacter.smithii | Visit.Name.NormD1 | 0.23652 |
| 177 | 0.04060 | 0.01951 | 68 | 2.08103 | 0.04120 | Veillonella.atypica | Visit.Name.NormD1 | 0.23652 |
| 178 | 0.01811 | 0.00880 | 68 | 2.05895 | 0.04333 | Megasphaera.massiliensis | Visit.Name.NormD1 | 0.23652 |
| 179 | 0.05405 | 0.02624 | 68 | 2.05946 | 0.04328 | Citrobacter.freundii | Visit.Name.NormD1 | 0.23652 |
| 180 | -0.03253 | 0.01600 | 68 | -2.03327 | 0.04593 | Blautia.sp..MP87G3 | Visit.Name.NormD1 | 0.24068 |
| 181 | 0.03557 | 0.01795 | 68 | 1.98170 | 0.05156 | Alistipes.sp..VE800.08 | Visit.Name.NormD14 | 0.24121 |
| 182 | -0.03046 | 0.01520 | 68 | -2.00437 | 0.04902 | Blautia.sp..MP40F7 | Visit.Name.NormD1 | 0.24696 |
| 183 | 0.03640 | 0.01624 | 68 | 2.24076 | 0.02831 | X.Ruminococcus..torques.CAG.61 | Visit.Name.NormD21 | 0.24725 |
| 184 | 0.03835 | 0.01931 | 68 | 1.98608 | 0.05106 | Veillonella.parvula | Visit.Name.NormD1 | 0.24772 |
| 185 | 0.03309 | 0.01496 | 68 | 2.21213 | 0.03032 | Bifidobacterium.dentium | Visit.Name.NormD21 | 0.24825 |
| 186 | -0.03538 | 0.01833 | 68 | -1.92989 | 0.05779 | Bifidobacterium.longum | Visit.Name.NormD14 | 0.26106 |
| 187 | 0.02083 | 0.00971 | 68 | 2.14444 | 0.03557 | Neglecta.timonensis | Visit.Name.NormD21 | 0.26242 |
| 188 | -0.02977 | 0.01401 | 68 | -2.12503 | 0.03722 | Blautia.sp..MP512E3 | Visit.Name.NormD21 | 0.26242 |
| 189 | -0.02315 | 0.01106 | 68 | -2.09320 | 0.04006 | Ruthenibacterium.lactatiformans | Visit.Name.NormD21 | 0.26242 |
| 190 | 0.02372 | 0.01123 | 68 | 2.11231 | 0.03834 | Absiella.sp..MP355B5 | Visit.Name.NormD21 | 0.26242 |
| 191 | 0.01826 | 0.00965 | 68 | 1.89262 | 0.06267 | Succinatimonas.hippei | Visit.Name.NormD14 | 0.27364 |
| 192 | 0.01372 | 0.00823 | 68 | 1.66672 | 0.10017 | Megasphaera.massiliensis | Visit.Name.NormD7 | 0.27920 |
| 193 | -0.02645 | 0.01416 | 68 | -1.86776 | 0.06611 | Blautia.sp..HF17 | Visit.Name.NormD14 | 0.27935 |
| 194 | 0.02618 | 0.01323 | 68 | 1.97934 | 0.05183 | Blautia.coccoides | Visit.Name.NormD21 | 0.28180 |
| 195 | 0.05449 | 0.02722 | 68 | 2.00190 | 0.04929 | Alistipes.sp..AF14.19 | Visit.Name.NormD21 | 0.28180 |
| 196 | -0.02901 | 0.01480 | 68 | -1.95937 | 0.05417 | Roseburia.intestinalis | Visit.Name.NormD21 | 0.28180 |
| 197 | -0.02474 | 0.01265 | 68 | -1.95580 | 0.05460 | Alistipes.sp..MP237E6a | Visit.Name.NormD21 | 0.28180 |
| 198 | -0.04180 | 0.02049 | 68 | -2.03956 | 0.04528 | Absiella.sp..MP391A4 | Visit.Name.NormD21 | 0.28180 |
| 199 | -0.09242 | 0.04752 | 68 | -1.94484 | 0.05593 | Lactobacillus.plantarum | Visit.Name.NormD21 | 0.28180 |
| 200 | -0.03648 | 0.01396 | 68 | -2.61364 | 0.01102 | Blautia.sp..MP341G7 | Visit.Name.NormD58 | 0.28873 |
| 201 | -0.06898 | 0.02546 | 68 | -2.70905 | 0.00853 | Absiella.sp..MP391A4 | Visit.Name.NormD58 | 0.28873 |
| 202 | 0.05712 | 0.02143 | 68 | 2.66593 | 0.00958 | Bacteroides.sp..MP227D2 | Visit.Name.NormD58 | 0.28873 |
| 203 | -0.03981 | 0.02096 | 68 | -1.89929 | 0.06177 | Bacteroides.sp..MP264A7 | Visit.Name.NormD1 | 0.28899 |
| 204 | 0.07540 | 0.04603 | 68 | 1.63806 | 0.10603 | Bacteroides.sp..VE800.09 | Visit.Name.NormD7 | 0.28937 |
| 205 | 0.01963 | 0.00972 | 68 | 2.01902 | 0.04743 | Neglecta.timonensis | Visit.Name.NormD30 | 0.29967 |
| 206 | 0.00841 | 0.00424 | 68 | 1.98449 | 0.05124 | Phascolarctobacterium.succinatutens | Visit.Name.NormD30 | 0.29967 |
| 207 | 0.02999 | 0.01520 | 68 | 1.97256 | 0.05261 | Bacteroides.sp..MP244D10 | Visit.Name.NormD30 | 0.29967 |
| 208 | 0.02405 | 0.01205 | 68 | 1.99550 | 0.05000 | Gemella.bergeri | Visit.Name.NormD30 | 0.29967 |
| 209 | -0.03694 | 0.01906 | 68 | -1.93764 | 0.05682 | Bifidobacterium.longum | Visit.Name.NormD30 | 0.29994 |
| 210 | -0.02178 | 0.01126 | 68 | -1.93429 | 0.05724 | Blautia.sp..MP78G6 | Visit.Name.NormD30 | 0.29994 |
| 211 | -0.01777 | 0.01101 | 68 | -1.61343 | 0.11128 | X.Clostridium..leptum | Visit.Name.NormD3 | 0.30371 |
| 212 | 0.07209 | 0.03810 | 68 | 1.89234 | 0.06270 | Fusobacterium.sp..VE800.01 | Visit.Name.NormD21 | 0.30423 |
| 213 | -0.03559 | 0.01906 | 68 | -1.86710 | 0.06620 | Bifidobacterium.longum | Visit.Name.NormD21 | 0.30972 |
| 214 | 0.09473 | 0.06046 | 68 | 1.56700 | 0.12176 | Klebsiella.pneumoniae | Visit.Name.NormD3 | 0.31485 |
| 215 | 0.01683 | 0.01084 | 68 | 1.55337 | 0.12498 | Desulfovibrio.fairfieldensis | Visit.Name.NormD3 | 0.31485 |
| 216 | 0.01255 | 0.00803 | 68 | 1.56302 | 0.12269 | Megasphaera.massiliensis | Visit.Name.NormD3 | 0.31485 |
| 217 | -0.01891 | 0.01194 | 68 | -1.58333 | 0.11799 | Ruminococcus.sp..MP226F3 | Visit.Name.NormD3 | 0.31485 |
| 218 | -0.01824 | 0.01155 | 68 | -1.58000 | 0.11875 | Lachnospiraceae.bacterium.5_1_57FAA | Visit.Name.NormD7 | 0.31747 |
| 219 | 0.01704 | 0.00927 | 68 | 1.83865 | 0.07033 | X.Clostridium..aldenense | Visit.Name.NormD21 | 0.31771 |
| 220 | -0.03407 | 0.02215 | 68 | -1.53838 | 0.12860 | Eubacterium.sp..CAG.180 | Visit.Name.NormD3 | 0.31785 |
| 221 | 0.02025 | 0.01083 | 68 | 1.87054 | 0.06571 | Desulfovibrio.fairfieldensis | Visit.Name.NormD30 | 0.31883 |
| 222 | -0.08961 | 0.04754 | 68 | -1.88483 | 0.06373 | Lactobacillus.plantarum | Visit.Name.NormD30 | 0.31883 |
| 223 | 0.02378 | 0.01310 | 68 | 1.81561 | 0.07384 | Flavobacteriales.bacterium | Visit.Name.NormD21 | 0.32243 |

| | | | | | | | | |
|---|---|---|---|---|---|---|---|---|
| 224 | 0.02543 | 0.01464 | 68 | 1.73684 | 0.08694 | Klebsiella.oxytoca | Visit.Name.NormD1 | 0.32543 |
| 225 | -0.06715 | 0.03897 | 68 | -1.72301 | 0.08943 | Bacteroides.thetaiotaomicron | Visit.Name.NormD1 | 0.32543 |
| 226 | -0.03423 | 0.01977 | 68 | -1.73124 | 0.08794 | Blautia.sp..MP588D7 | Visit.Name.NormD1 | 0.32543 |
| 227 | -0.03870 | 0.02228 | 68 | -1.73750 | 0.08683 | Absiella.sp..MP391A4 | Visit.Name.NormD1 | 0.32543 |
| 228 | 0.01680 | 0.00962 | 68 | 1.74614 | 0.08530 | Megasphaera.sp..MP182G6 | Visit.Name.NormD1 | 0.32543 |
| 229 | -0.02218 | 0.01228 | 68 | -1.80661 | 0.07525 | Roseburia.faecis | Visit.Name.NormD1 | 0.32543 |
| 230 | -0.02251 | 0.01302 | 68 | -1.72919 | 0.08831 | Ruminococcus.sp..MP226F3 | Visit.Name.NormD1 | 0.32543 |
| 231 | 0.04377 | 0.02515 | 68 | 1.74076 | 0.08625 | Lactobacillus.salivarius | Visit.Name.NormD1 | 0.32543 |
| 232 | 0.03235 | 0.01771 | 68 | 1.82659 | 0.07215 | Parabacteroides.sp..VE800.05 | Visit.Name.NormD30 | 0.32592 |
| 233 | -0.01439 | 0.00786 | 68 | -1.83007 | 0.07162 | Sutterella.sp..AM11.39 | Visit.Name.NormD30 | 0.32592 |
| 234 | 0.02941 | 0.01624 | 68 | 1.81037 | 0.07466 | X.Ruminococcus..torques.CAG.61 | Visit.Name.NormD30 | 0.32600 |
| 235 | -0.08061 | 0.04552 | 68 | -1.77071 | 0.08109 | Lactobacillus.plantarum | Visit.Name.NormD14 | 0.33196 |
| 236 | 0.02562 | 0.01496 | 68 | 1.71293 | 0.09128 | Bifidobacterium.dentium | Visit.Name.NormD30 | 0.33216 |
| 237 | -0.03770 | 0.02196 | 68 | -1.71687 | 0.09055 | Bacteroides.vulgatus | Visit.Name.NormD30 | 0.33216 |
| 238 | -0.01796 | 0.01032 | 68 | -1.73963 | 0.08645 | Lactobacillus.paracasei | Visit.Name.NormD30 | 0.33216 |
| 239 | -0.01937 | 0.01106 | 68 | -1.75020 | 0.08459 | Ruthenibacterium.lactatiformans | Visit.Name.NormD30 | 0.33216 |
| 240 | -0.02770 | 0.01615 | 68 | -1.71578 | 0.09076 | Blautia.sp..MP245E9 | Visit.Name.NormD30 | 0.33216 |
| 241 | -0.02532 | 0.01434 | 68 | -1.76494 | 0.08206 | Enterococcus.faecalis | Visit.Name.NormD30 | 0.33216 |
| 242 | -0.01691 | 0.01126 | 68 | -1.50182 | 0.13777 | Lachnospiraceae.bacterium.5_1_57FAA | Visit.Name.NormD3 | 0.33423 |
| 243 | -0.05687 | 0.03814 | 68 | -1.49138 | 0.14049 | Bacteroides.fragilis | Visit.Name.NormD3 | 0.33461 |
| 244 | -0.01836 | 0.01032 | 68 | -1.77950 | 0.07963 | Lactobacillus.paracasei | Visit.Name.NormD21 | 0.33649 |
| 245 | -0.02119 | 0.01212 | 68 | -1.74851 | 0.08489 | HI22.sp..HI22 | Visit.Name.NormD14 | 0.33698 |
| 246 | -0.03718 | 0.02201 | 68 | -1.68926 | 0.09575 | Lactobacillus.fermentum | Visit.Name.NormD30 | 0.33901 |
| 247 | 0.02528 | 0.01723 | 68 | 1.46735 | 0.14689 | Colibacter.massiliensis | Visit.Name.NormD3 | 0.34362 |
| 248 | 0.02331 | 0.01353 | 68 | 1.72322 | 0.08939 | Bacteroides.sp..MP369D12 | Visit.Name.NormD21 | 0.34443 |
| 249 | -0.01355 | 0.00785 | 68 | -1.72572 | 0.08894 | Sutterella.sp..AM11.39 | Visit.Name.NormD21 | 0.34443 |
| 250 | 0.02304 | 0.01323 | 68 | 1.74206 | 0.08602 | Collinsella.tanakaei | Visit.Name.NormD21 | 0.34443 |
| 251 | 0.02192 | 0.01324 | 68 | 1.65616 | 0.10230 | Blautia.coccoides | Visit.Name.NormD30 | 0.35266 |
| 252 | 0.01754 | 0.01217 | 68 | 1.44136 | 0.15407 | Methanobrevibacter.smithii | Visit.Name.NormD3 | 0.35410 |
| 253 | -0.02108 | 0.01234 | 68 | -1.70785 | 0.09223 | Tyzzerella.sp..MP10F1re | Visit.Name.NormD14 | 0.35534 |
| 254 | -0.03580 | 0.02200 | 68 | -1.62682 | 0.10840 | Lactobacillus.fermentum | Visit.Name.NormD21 | 0.35816 |
| 255 | -0.02063 | 0.01235 | 68 | -1.67010 | 0.09950 | Eggerthella.lenta | Visit.Name.NormD21 | 0.35816 |
| 256 | 0.03863 | 0.02360 | 68 | 1.63672 | 0.10631 | X.Clostridium..clostridioforme | Visit.Name.NormD21 | 0.35816 |
| 257 | 0.03085 | 0.01874 | 68 | 1.64575 | 0.10443 | Alistipes.sp..VE800.08 | Visit.Name.NormD21 | 0.35816 |
| 258 | 0.02578 | 0.01526 | 68 | 1.68861 | 0.09588 | Acidaminococcus.intestini | Visit.Name.NormD21 | 0.35816 |
| 259 | 0.04349 | 0.02681 | 68 | 1.62232 | 0.10936 | Bacteroides.sp..MP487B8 | Visit.Name.NormD21 | 0.35816 |
| 260 | 0.03475 | 0.02324 | 68 | 1.49515 | 0.13950 | Ruthenibacterium.sp..VE800.03 | Visit.Name.NormD7 | 0.35833 |
| 261 | -0.01428 | 0.00952 | 68 | -1.50041 | 0.13814 | X.Clostridium..aldenense | Visit.Name.NormD7 | 0.35833 |
| 262 | 0.02285 | 0.01405 | 68 | 1.62566 | 0.10865 | X.Ruminococcus..sp..MP50A2 | Visit.Name.NormD30 | 0.36494 |
| 263 | -0.04069 | 0.02891 | 68 | -1.40766 | 0.16379 | Alistipes.sp..MP104G4_1 | Visit.Name.NormD3 | 0.36824 |
| 264 | 0.05288 | 0.03796 | 68 | 1.39321 | 0.16809 | Akkermansia.muciniphila | Visit.Name.NormD3 | 0.36824 |
| 265 | 0.06617 | 0.04756 | 68 | 1.39133 | 0.16866 | Lactobacillus.plantarum | Visit.Name.NormD3 | 0.36824 |
| 266 | 0.01322 | 0.00900 | 68 | 1.46888 | 0.14648 | Megasphaera.sp..MP182G6 | Visit.Name.NormD7 | 0.36901 |
| 267 | 0.01676 | 0.01055 | 68 | 1.58889 | 0.11672 | Absiella.sp..MP634G1_ig1 | Visit.Name.NormD21 | 0.37295 |
| 268 | 0.02646 | 0.01834 | 68 | 1.44255 | 0.15374 | Veillonella.atypica | Visit.Name.NormD7 | 0.37999 |
| 269 | -0.02784 | 0.01677 | 68 | -1.65987 | 0.10155 | Bacteroides.xylanisolvens | Visit.Name.NormD14 | 0.38007 |
| 270 | -0.01758 | 0.01126 | 68 | -1.56223 | 0.12288 | Blautia.sp..MP78G6 | Visit.Name.NormD21 | 0.38325 |
| 271 | -0.04495 | 0.03330 | 68 | -1.34982 | 0.18155 | Bacteroides.ovatus | Visit.Name.NormD3 | 0.38360 |
| 272 | -0.01947 | 0.01435 | 68 | -1.35624 | 0.17951 | Enterococcus.faecalis | Visit.Name.NormD3 | 0.38360 |
| 273 | 0.02541 | 0.01769 | 68 | 1.43619 | 0.15553 | Parabacteroides.sp..VE800.05 | Visit.Name.NormD21 | 0.38443 |
| 274 | 0.01630 | 0.01124 | 68 | 1.45080 | 0.15143 | Lachnospiraceae.bacterium.5_1_57FAA | Visit.Name.NormD21 | 0.38443 |
| 275 | 0.01797 | 0.01222 | 68 | 1.47080 | 0.14596 | Blautia.sp..MP351A5 | Visit.Name.NormD21 | 0.38443 |
| 276 | 0.00609 | 0.00423 | 68 | 1.43888 | 0.15477 | Phascolarctobacterium.succinatutens | Visit.Name.NormD21 | 0.38443 |
| 277 | 0.02149 | 0.01469 | 68 | 1.46284 | 0.14812 | Blautia.sp..MP87G3 | Visit.Name.NormD21 | 0.38443 |
| 278 | -0.01719 | 0.01193 | 68 | -1.44071 | 0.15426 | Ruminococcus.sp..MP226F3 | Visit.Name.NormD21 | 0.38443 |
| 279 | 0.01642 | 0.01137 | 68 | 1.44497 | 0.15306 | Pseudomonas.aeruginosa | Visit.Name.NormD21 | 0.38443 |
| 280 | -0.01909 | 0.01287 | 68 | -1.48325 | 0.14263 | Tyzzerella.sp..MP10F1re | Visit.Name.NormD21 | 0.38443 |
| 281 | -0.02646 | 0.01761 | 68 | -1.50301 | 0.13747 | Solobacterium.moorei | Visit.Name.NormD21 | 0.38443 |
| 282 | 0.02305 | 0.01596 | 68 | 1.44395 | 0.15335 | Shigella.dysenteriae | Visit.Name.NormD21 | 0.38443 |
| 283 | -0.01944 | 0.01275 | 68 | -1.52419 | 0.13210 | Enterobacteriaceae.bacterium.9_2_54FAA | Visit.Name.NormD21 | 0.38443 |
| 284 | -0.04825 | 0.03412 | 68 | -1.41387 | 0.16196 | Bacteroides.ovatus | Visit.Name.NormD7 | 0.38576 |
| 285 | -0.02372 | 0.01673 | 68 | -1.41760 | 0.16087 | Blautia.sp..HK34 | Visit.Name.NormD7 | 0.38576 |
| 286 | -0.01920 | 0.01184 | 68 | -1.62210 | 0.10941 | Oscillospiraceae.bacterium.VE202.24 | Visit.Name.NormD1 | 0.38737 |
| 287 | -0.01579 | 0.01129 | 68 | -1.39798 | 0.16666 | X.Clostridium..leptum | Visit.Name.NormD7 | 0.38755 |
| 288 | -0.02394 | 0.01720 | 68 | -1.39144 | 0.16863 | Clostridioides.difficile | Visit.Name.NormD7 | 0.38755 |
| 289 | 0.02387 | 0.01461 | 68 | 1.63362 | 0.10696 | Acidaminococcus.intestini | Visit.Name.NormD14 | 0.38922 |
| 290 | -0.01213 | 0.00750 | 68 | -1.61728 | 0.11045 | Sutterella.sp..AM11.39 | Visit.Name.NormD14 | 0.39105 |
| 291 | -0.06283 | 0.03919 | 68 | -1.60311 | 0.11355 | Parabacteroides.distasonis | Visit.Name.NormD1 | 0.39143 |
| 292 | 0.01522 | 0.01081 | 68 | 1.40783 | 0.16374 | Oscillospiraceae.bacterium.VE202.24 | Visit.Name.NormD21 | 0.39721 |
| 293 | 0.05973 | 0.03812 | 68 | 1.56695 | 0.12177 | Fusobacterium.sp..VE800.01 | Visit.Name.NormD30 | 0.39879 |
| 294 | -0.03348 | 0.02111 | 68 | -1.58596 | 0.11739 | Lactobacillus.fermentum | Visit.Name.NormD14 | 0.40004 |
| 295 | 0.02824 | 0.01829 | 68 | 1.54451 | 0.12711 | Lachnospiraceae.bacterium.7_1_58FAA | Visit.Name.NormD14 | 0.40004 |
| 296 | -0.01537 | 0.00988 | 68 | -1.55638 | 0.12426 | Lactobacillus.paracasei | Visit.Name.NormD14 | 0.40004 |
| 297 | -0.01877 | 0.01214 | 68 | -1.54625 | 0.12669 | Alistipes.sp..MP237E6a | Visit.Name.NormD14 | 0.40004 |
| 298 | -0.02301 | 0.01498 | 68 | -1.53598 | 0.12918 | Eisenbergiella.sp..OF01.20 | Visit.Name.NormD14 | 0.40004 |

| | | | | | | | | |
|---|---|---|---|---|---|---|---|---|
| 299 | 0.02177 | 0.01425 | 68 | 1.52736 | 0.13131 | Atopobium.minutum | Visit.Name.NormD14 | 0.40004 |
| 300 | -0.01884 | 0.01434 | 68 | -1.31427 | 0.19317 | Acidaminococcus.intestini.CAG.325 | Visit.Name.NormD3 | 0.40168 |
| 301 | -0.02097 | 0.01583 | 68 | -1.32531 | 0.18950 | Erysipelotrichaceae.bacterium.6_1_45 | Visit.Name.NormD7 | 0.40712 |
| 302 | -0.01779 | 0.01342 | 68 | -1.32510 | 0.18957 | Barnesiella.intestinihominis | Visit.Name.NormD7 | 0.40712 |
| 303 | 0.00576 | 0.00435 | 68 | 1.32531 | 0.18950 | Phascolarctobacterium.succinatutens | Visit.Name.NormD7 | 0.40712 |
| 304 | 0.01394 | 0.01037 | 68 | 1.34360 | 0.18354 | Succinatimonas.hippei | Visit.Name.NormD7 | 0.40712 |
| 305 | 0.01555 | 0.01033 | 68 | 1.50451 | 0.13708 | Desulfovibrio.fairfieldensis | Visit.Name.NormD14 | 0.40813 |
| 306 | -0.01889 | 0.01442 | 68 | -1.30981 | 0.19467 | X.Ruminococcus..sp..MP50A2 | Visit.Name.NormD7 | 0.41132 |
| 307 | 0.01805 | 0.01314 | 68 | 1.37410 | 0.17392 | Ruminococcus.bicirculans | Visit.Name.NormD21 | 0.41425 |
| 308 | -0.03134 | 0.02112 | 68 | -1.48376 | 0.14249 | Bacteroides.vulgatus | Visit.Name.NormD14 | 0.41482 |
| 309 | 0.04492 | 0.02895 | 68 | 1.55148 | 0.12543 | Raoultella.ornithinolytica | Visit.Name.NormD1 | 0.42131 |
| 310 | -0.02387 | 0.01906 | 68 | -1.25235 | 0.21473 | Bifidobacterium.longum | Visit.Name.NormD3 | 0.43277 |
| 311 | -0.01315 | 0.01043 | 68 | -1.26059 | 0.21176 | Eubacterium.sp..CAG.38 | Visit.Name.NormD3 | 0.43277 |
| 312 | -0.01919 | 0.01276 | 68 | -1.50393 | 0.13723 | Enterobacteriaceae.bacterium.9_2_54FAA | Visit.Name.NormD30 | 0.43847 |
| 313 | -0.03213 | 0.02548 | 68 | -1.26113 | 0.21157 | Alistipes.finegoldii | Visit.Name.NormD7 | 0.43993 |
| 314 | 0.01840 | 0.01473 | 68 | 1.24918 | 0.21588 | Pediococcus.acidilactici | Visit.Name.NormD7 | 0.44189 |
| 315 | 0.03486 | 0.02362 | 68 | 1.47577 | 0.14462 | X.Clostridium..clostridioforme | Visit.Name.NormD30 | 0.44899 |
| 316 | 0.03991 | 0.02723 | 68 | 1.46555 | 0.14738 | Alistipes.sp..AF14.19 | Visit.Name.NormD30 | 0.44899 |
| 317 | 0.04386 | 0.03329 | 68 | 1.31737 | 0.19214 | Bacteroides.ovatus | Visit.Name.NormD21 | 0.44946 |
| 318 | -0.01392 | 0.01152 | 68 | -1.20881 | 0.23092 | X.Clostridium..citroniae | Visit.Name.NormD3 | 0.45150 |
| 319 | 0.01667 | 0.01371 | 68 | 1.21605 | 0.22817 | Desulfovibrio.desulfuricans | Visit.Name.NormD3 | 0.45150 |
| 320 | 0.01397 | 0.01141 | 68 | 1.22401 | 0.22517 | Veillonella.sp..MP19D1_02.09 | Visit.Name.NormD7 | 0.45381 |
| 321 | 0.01482 | 0.01079 | 68 | 1.37379 | 0.17402 | Blautia.sp..VE303.05 | Visit.Name.NormD14 | 0.45593 |
| 322 | -0.01763 | 0.01255 | 68 | -1.40460 | 0.16469 | Barnesiella.intestinihominis | Visit.Name.NormD14 | 0.45593 |
| 323 | 0.01486 | 0.01076 | 68 | 1.38117 | 0.17175 | Absiella.sp..MP355B5 | Visit.Name.NormD14 | 0.45593 |
| 324 | -0.01689 | 0.01222 | 68 | -1.38250 | 0.17134 | Enterobacteriaceae.bacterium.9_2_54FAA | Visit.Name.NormD14 | 0.45593 |
| 325 | 0.01590 | 0.01150 | 68 | 1.38177 | 0.17157 | Gemella.bergeri | Visit.Name.NormD14 | 0.45593 |
| 326 | 0.01332 | 0.00928 | 68 | 1.43583 | 0.15563 | X.Clostridium..aldenense | Visit.Name.NormD30 | 0.46336 |
| 327 | -0.02331 | 0.01570 | 68 | -1.48495 | 0.14218 | Enterococcus.faecalis | Visit.Name.NormD1 | 0.46564 |
| 328 | 0.01772 | 0.01411 | 68 | 1.25564 | 0.21354 | Bacteroides.dorei | Visit.Name.NormD21 | 0.46624 |
| 329 | 0.01338 | 0.01065 | 68 | 1.25656 | 0.21321 | Clostridiales.bacterium | Visit.Name.NormD21 | 0.46624 |
| 330 | -0.01825 | 0.01433 | 68 | -1.27373 | 0.20710 | Enterococcus.faecalis | Visit.Name.NormD21 | 0.46624 |
| 331 | -0.00784 | 0.00623 | 68 | -1.25687 | 0.21310 | Sutterella.sp..54_7 | Visit.Name.NormD21 | 0.46624 |
| 332 | -0.06124 | 0.04179 | 68 | -1.46561 | 0.14736 | Bacteroides.fragilis | Visit.Name.NormD1 | 0.47085 |
| 333 | -0.02826 | 0.02411 | 68 | -1.17203 | 0.24528 | X.Ruminococcus..sp..MP495E3 | Visit.Name.NormD3 | 0.47252 |
| 334 | 0.01819 | 0.01356 | 68 | 1.34140 | 0.18425 | Bacteroides.sp..MP327C3 | Visit.Name.NormD14 | 0.47328 |
| 335 | -0.01991 | 0.01615 | 68 | -1.23299 | 0.22183 | Blautia.sp..MP245E9 | Visit.Name.NormD21 | 0.47638 |
| 336 | -0.01425 | 0.01074 | 68 | -1.32652 | 0.18910 | Lachnospiraceae.bacterium.5_1_57FAA | Visit.Name.NormD14 | 0.47640 |
| 337 | 0.01696 | 0.01496 | 68 | 1.13415 | 0.26071 | Bifidobacterium.dentium | Visit.Name.NormD3 | 0.48103 |
| 338 | -0.01489 | 0.01308 | 68 | -1.13796 | 0.25913 | Roseburia.hominis | Visit.Name.NormD3 | 0.48103 |
| 339 | 0.01175 | 0.01033 | 68 | 1.13758 | 0.25929 | Lactobacillus.paracasei | Visit.Name.NormD3 | 0.48103 |
| 340 | -0.02221 | 0.01545 | 68 | -1.43709 | 0.15528 | Bacteroides.dorei | Visit.Name.NormD1 | 0.48166 |
| 341 | 0.01581 | 0.01108 | 68 | 1.42718 | 0.15810 | Succinatimonas.hippei | Visit.Name.NormD1 | 0.48166 |
| 342 | -0.04477 | 0.03194 | 68 | -1.40181 | 0.16552 | Lachnospiraceae.bacterium.1_1_57FAA | Visit.Name.NormD30 | 0.48185 |
| 343 | -0.01568 | 0.01406 | 68 | -1.11471 | 0.26890 | X.Ruminococcus..sp..MP50A2 | Visit.Name.NormD3 | 0.48925 |
| 344 | 0.01358 | 0.01125 | 68 | 1.20652 | 0.23180 | Blautia.sp..VE303.05 | Visit.Name.NormD21 | 0.48977 |
| 345 | -0.01238 | 0.01069 | 68 | -1.15716 | 0.25125 | Eubacterium.sp..CAG.38 | Visit.Name.NormD7 | 0.49126 |
| 346 | -0.01376 | 0.01180 | 68 | -1.16601 | 0.24769 | X.Clostridium..citroniae | Visit.Name.NormD7 | 0.49126 |
| 347 | -0.03395 | 0.02419 | 68 | -1.40355 | 0.16500 | Eubacterium.sp..CAG.180 | Visit.Name.NormD1 | 0.49126 |
| 348 | -0.01773 | 0.01286 | 68 | -1.37854 | 0.17256 | Alistipes.putredinis | Visit.Name.NormD1 | 0.49141 |
| 349 | -0.02160 | 0.01565 | 68 | -1.37992 | 0.17213 | Acidaminococcus.intestini.CAG.325 | Visit.Name.NormD1 | 0.49141 |
| 350 | -0.01787 | 0.01632 | 68 | -1.09498 | 0.27739 | Blautia.sp..HK34 | Visit.Name.NormD3 | 0.49778 |
| 351 | -0.01674 | 0.01472 | 68 | -1.13762 | 0.25927 | Enterococcus.faecalis | Visit.Name.NormD7 | 0.49948 |
| 352 | 0.02645 | 0.02360 | 68 | 1.12095 | 0.26625 | Lactobacillus.salivarius | Visit.Name.NormD7 | 0.50549 |
| 353 | -0.01619 | 0.01194 | 68 | -1.35647 | 0.17944 | Ruminococcus.sp..MP226F3 | Visit.Name.NormD30 | 0.51101 |
| 354 | 0.01991 | 0.01702 | 68 | 1.16976 | 0.24618 | Bacteroides.sp..MP227D2 | Visit.Name.NormD21 | 0.51190 |
| 355 | 0.02228 | 0.02080 | 68 | 1.07099 | 0.28796 | Cloacibacillus.evryensis | Visit.Name.NormD7 | 0.52003 |
| 356 | -0.01461 | 0.01341 | 68 | -1.08950 | 0.27978 | Roseburia.hominis | Visit.Name.NormD7 | 0.52003 |
| 357 | 0.00753 | 0.00707 | 68 | 1.06452 | 0.29086 | Megasphaera.sp..UBA1696 | Visit.Name.NormD7 | 0.52003 |
| 358 | 0.02592 | 0.02425 | 68 | 1.06864 | 0.28901 | Bacteroides.sp..MP163B7 | Visit.Name.NormD7 | 0.52003 |
| 359 | 0.01734 | 0.01639 | 68 | 1.05809 | 0.29376 | Shigella.dysenteriae | Visit.Name.NormD7 | 0.52003 |
| 360 | 0.04799 | 0.02290 | 68 | 2.09518 | 0.03988 | Phascolarctobacterium.sp..VE800.04 | Visit.Name.NormD58 | 0.52246 |
| 361 | 0.10606 | 0.04808 | 68 | 2.20604 | 0.03076 | Parabacteroides.sp..VE800.07 | Visit.Name.NormD58 | 0.52246 |
| 362 | -0.03558 | 0.01661 | 68 | -2.14214 | 0.03577 | X.Eubacterium..hallii | Visit.Name.NormD58 | 0.52246 |
| 363 | -0.04008 | 0.01890 | 68 | -2.12068 | 0.03760 | Blautia.sp..MP237F5b | Visit.Name.NormD58 | 0.52246 |
| 364 | 0.07160 | 0.03385 | 68 | 2.11527 | 0.03807 | Bacteroides.sp..MP487B8 | Visit.Name.NormD58 | 0.52246 |
| 365 | -0.02089 | 0.01570 | 68 | -1.33086 | 0.18768 | Eisenbergiella.sp..OF01.20 | Visit.Name.NormD30 | 0.52311 |
| 366 | -0.01347 | 0.01078 | 68 | -1.24896 | 0.21596 | Roseburia.faecis | Visit.Name.NormD14 | 0.52391 |
| 367 | 0.03211 | 0.02563 | 68 | 1.25264 | 0.21463 | Bacteroides.sp..MP487B8 | Visit.Name.NormD14 | 0.52391 |
| 368 | 0.01238 | 0.01081 | 68 | 1.14490 | 0.25626 | Desulfovibrio.fairfieldensis | Visit.Name.NormD21 | 0.52454 |
| 369 | 0.01394 | 0.01325 | 68 | 1.05214 | 0.29646 | Collinsella.tanakaei | Visit.Name.NormD3 | 0.52481 |
| 370 | 0.03352 | 0.01713 | 68 | 1.95716 | 0.05443 | Eubacterium.sp..VE800.02 | Visit.Name.NormD58 | 0.52489 |
| 371 | 0.07202 | 0.03819 | 68 | 1.88584 | 0.06359 | Parabacteroides.sp..VE800.10 | Visit.Name.NormD58 | 0.52489 |
| 372 | 0.02597 | 0.01305 | 68 | 1.99049 | 0.05056 | Eubacterium.sp..CAG.38 | Visit.Name.NormD58 | 0.52489 |
| 373 | -0.03062 | 0.01576 | 68 | -1.94244 | 0.05623 | Alistipes.sp..MP237E6a | Visit.Name.NormD58 | 0.52489 |

| | | | | | | | | |
|---|---|---|---|---|---|---|---|---|
| 374 | -0.11739 | 0.05959 | 68 | -1.96995 | 0.05292 | Lactobacillus.plantarum | Visit.Name.NormD58 | 0.52489 |
| 375 | 0.03045 | 0.01618 | 68 | 1.88205 | 0.06411 | Intestinibacter.sp..MP113D11 | Visit.Name.NormD58 | 0.52489 |
| 376 | 0.01449 | 0.01419 | 68 | 1.02109 | 0.31083 | Phascolarctobacterium.sp..MUR_MP6E9_ig1 | Visit.Name.NormD7 | 0.52502 |
| 377 | -0.00837 | 0.00807 | 68 | -1.03653 | 0.30362 | Sutterella.sp..AM11.39 | Visit.Name.NormD7 | 0.52502 |
| 378 | -0.04039 | 0.03911 | 68 | -1.03269 | 0.30541 | Bacteroides.fragilis | Visit.Name.NormD7 | 0.52502 |
| 379 | -0.01245 | 0.01224 | 68 | -1.01732 | 0.31261 | Ruminococcus.sp..MP226F3 | Visit.Name.NormD7 | 0.52502 |
| 380 | 0.01606 | 0.01544 | 68 | 1.04004 | 0.30201 | Erysipelotrichaceae.bacterium.6_1_45 | Visit.Name.NormD3 | 0.52751 |
| 381 | 0.02443 | 0.01876 | 68 | 1.30265 | 0.19709 | Alistipes.sp..VE800.08 | Visit.Name.NormD30 | 0.52786 |
| 382 | 0.01358 | 0.01043 | 68 | 1.30160 | 0.19745 | Eubacterium.sp..CAG.38 | Visit.Name.NormD30 | 0.52786 |
| 383 | 0.05727 | 0.05648 | 68 | 1.01389 | 0.31423 | Escherichia.coli | Visit.Name.NormD3 | 0.53460 |
| 384 | 0.01972 | 0.01940 | 68 | 1.01653 | 0.31298 | Acidaminococcus.sp..MP76F2 | Visit.Name.NormD3 | 0.53460 |
| 385 | -0.01547 | 0.01382 | 68 | -1.11934 | 0.26693 | Phascolarctobacterium.sp..MUR_MP6E9_ig1 | Visit.Name.NormD21 | 0.53797 |
| 386 | 0.01230 | 0.01008 | 68 | 1.21992 | 0.22671 | Absiella.sp..MP634G1_ig1 | Visit.Name.NormD14 | 0.53998 |
| 387 | -0.02699 | 0.02067 | 68 | -1.30574 | 0.19604 | Bifidobacterium.longum | Visit.Name.NormD1 | 0.54641 |
| 388 | 0.01770 | 0.01817 | 68 | 0.97381 | 0.33360 | Parabacteroides.sp..VE800.05 | Visit.Name.NormD7 | 0.55319 |
| 389 | 0.03778 | 0.03886 | 68 | 0.97222 | 0.33439 | Pyramidobacter.piscolens | Visit.Name.NormD3 | 0.56160 |
| 390 | 0.08780 | 0.04819 | 68 | 1.82174 | 0.07289 | Bacteroides.fragilis | Visit.Name.NormD58 | 0.56170 |
| 391 | 0.01905 | 0.01527 | 68 | 1.24683 | 0.21674 | Acidaminococcus.intestini | Visit.Name.NormD30 | 0.56785 |
| 392 | 0.01742 | 0.01481 | 68 | 1.17602 | 0.24369 | Erysipelotrichaceae.bacterium.6_1_45 | Visit.Name.NormD14 | 0.57005 |
| 393 | 0.01185 | 0.01018 | 68 | 1.16456 | 0.24827 | Clostridiales.bacterium | Visit.Name.NormD14 | 0.57058 |
| 394 | 0.02520 | 0.02359 | 68 | 1.06832 | 0.28915 | Bacteroides.sp..MP163B7 | Visit.Name.NormD21 | 0.57392 |
| 395 | 0.01071 | 0.01139 | 68 | 0.94034 | 0.35037 | Pseudomonas.aeruginosa | Visit.Name.NormD3 | 0.57469 |
| 396 | 0.00950 | 0.01012 | 68 | 0.93919 | 0.35096 | Succinatimonas.hippei | Visit.Name.NormD3 | 0.57469 |
| 397 | 0.02137 | 0.01709 | 68 | 1.25099 | 0.21523 | Bilophila.wadsworthia | Visit.Name.NormD1 | 0.57804 |
| 398 | -0.01268 | 0.01016 | 68 | -1.24827 | 0.21621 | X.Clostridium..aldenense | Visit.Name.NormD1 | 0.57804 |
| 399 | -0.02371 | 0.01919 | 68 | -1.23593 | 0.22074 | Bacteroides.xylanisolvens | Visit.Name.NormD1 | 0.57833 |
| 400 | 0.01277 | 0.01064 | 68 | 1.19982 | 0.23437 | Neglecta.timonensis | Visit.Name.NormD1 | 0.60202 |
| 401 | 0.01375 | 0.01232 | 68 | 1.11641 | 0.26818 | Intestinibacter.sp..MP113D11 | Visit.Name.NormD14 | 0.60571 |
| 402 | -0.00745 | 0.00624 | 68 | -1.19387 | 0.23668 | Sutterella.sp..54_7 | Visit.Name.NormD30 | 0.60793 |
| 403 | -0.03312 | 0.03685 | 68 | -0.89866 | 0.37201 | Parabacteroides.distasonis | Visit.Name.NormD7 | 0.60916 |
| 404 | -0.02277 | 0.01930 | 68 | -1.17975 | 0.24221 | Solobacterium.moorei | Visit.Name.NormD1 | 0.61018 |
| 405 | 0.01702 | 0.01444 | 68 | 1.17867 | 0.24264 | X.Clostridium..symbiosum | Visit.Name.NormD30 | 0.61126 |
| 406 | 0.01521 | 0.01383 | 68 | 1.09969 | 0.27534 | X.Clostridium..symbiosum | Visit.Name.NormD14 | 0.61136 |
| 407 | -0.01631 | 0.01402 | 68 | -1.16359 | 0.24866 | Blautia.sp..MP512E3 | Visit.Name.NormD30 | 0.61460 |
| 408 | -0.06182 | 0.03649 | 68 | -1.69428 | 0.09479 | Alistipes.sp..MP104G4_1 | Visit.Name.NormD58 | 0.61644 |
| 409 | 0.03775 | 0.02231 | 68 | 1.69183 | 0.09526 | Parabacteroides.sp..VE800.05 | Visit.Name.NormD58 | 0.61644 |
| 410 | -0.02227 | 0.01296 | 68 | -1.71859 | 0.09024 | Lactobacillus.paracasei | Visit.Name.NormD58 | 0.61644 |
| 411 | -0.02618 | 0.01578 | 68 | -1.65871 | 0.10178 | HI22.sp..HI22 | Visit.Name.NormD58 | 0.61644 |
| 412 | -0.03024 | 0.01833 | 68 | -1.65015 | 0.10352 | Blautia.sp..MP87G3 | Visit.Name.NormD58 | 0.61644 |
| 413 | -0.04322 | 0.02745 | 68 | -1.57449 | 0.12002 | Lactobacillus.fermentum | Visit.Name.NormD58 | 0.61976 |
| 414 | -0.02204 | 0.01390 | 68 | -1.58612 | 0.11735 | Ruthenibacterium.lactatiformans | Visit.Name.NormD58 | 0.61976 |
| 415 | -0.03109 | 0.01991 | 68 | -1.56168 | 0.12301 | Blautia.sp..MP245E9 | Visit.Name.NormD58 | 0.61976 |
| 416 | -0.01277 | 0.00790 | 68 | -1.61738 | 0.11043 | Sutterella.sp..54_7 | Visit.Name.NormD58 | 0.61976 |
| 417 | -0.00562 | 0.00641 | 68 | -0.87654 | 0.38382 | Sutterella.sp..54_7 | Visit.Name.NormD7 | 0.62075 |
| 418 | 0.06790 | 0.04793 | 68 | 1.41657 | 0.16117 | Fusobacterium.sp..VE800.01 | Visit.Name.NormD58 | 0.62099 |
| 419 | -0.03500 | 0.02356 | 68 | -1.48571 | 0.14198 | Bifidobacterium.longum | Visit.Name.NormD58 | 0.62099 |
| 420 | -0.02101 | 0.01482 | 68 | -1.41792 | 0.16078 | Alistipes.putredinis | Visit.Name.NormD58 | 0.62099 |
| 421 | 0.01755 | 0.01170 | 68 | 1.50102 | 0.13798 | X.Clostridium..aldenense | Visit.Name.NormD58 | 0.62099 |
| 422 | 0.02090 | 0.01426 | 68 | 1.46572 | 0.14733 | Dorea.sp..MP50E3 | Visit.Name.NormD58 | 0.62099 |
| 423 | -0.02522 | 0.01741 | 68 | -1.44857 | 0.15205 | Blautia.sp..MP40F7 | Visit.Name.NormD58 | 0.62099 |
| 424 | -0.02528 | 0.01745 | 68 | -1.44889 | 0.15196 | Phascolarctobacterium.sp..MUR_MP6E9_ig1 | Visit.Name.NormD58 | 0.62099 |
| 425 | -0.02386 | 0.01599 | 68 | -1.49194 | 0.14034 | Enterobacteriaceae.bacterium.9_2_54FAA | Visit.Name.NormD58 | 0.62099 |
| 426 | -0.09779 | 0.07092 | 68 | -1.37885 | 0.17246 | Escherichia.coli | Visit.Name.NormD58 | 0.62201 |
| 427 | -0.06175 | 0.04495 | 68 | -1.37365 | 0.17406 | Parabacteroides.distasonis | Visit.Name.NormD58 | 0.62201 |
| 428 | 0.01928 | 0.01409 | 68 | 1.36843 | 0.17568 | Roseburia.faecis | Visit.Name.NormD58 | 0.62201 |
| 429 | 0.01268 | 0.01481 | 68 | 0.85607 | 0.39497 | X.Clostridium..symbiosum | Visit.Name.NormD7 | 0.62338 |
| 430 | 0.01713 | 0.01990 | 68 | 0.86069 | 0.39244 | Acidaminococcus.sp..MP76F2 | Visit.Name.NormD7 | 0.62338 |
| 431 | -0.01289 | 0.01482 | 68 | -0.87031 | 0.38719 | Blautia.sp..HF17 | Visit.Name.NormD3 | 0.62411 |
| 432 | -0.01450 | 0.01678 | 68 | -0.86392 | 0.39067 | Clostridioides.difficile | Visit.Name.NormD3 | 0.62411 |
| 433 | 0.01655 | 0.01450 | 68 | 1.14102 | 0.25786 | Collinsella.tanakaei | Visit.Name.NormD1 | 0.62556 |
| 434 | 0.02109 | 0.01834 | 68 | 1.14985 | 0.25423 | Sutterella.wadsworthensis | Visit.Name.NormD1 | 0.62556 |
| 435 | -0.01416 | 0.01436 | 68 | -0.98624 | 0.32751 | Pediococcus.acidilactici | Visit.Name.NormD21 | 0.62713 |
| 436 | 0.00990 | 0.01009 | 68 | 0.98049 | 0.33032 | Succinatimonas.hippei | Visit.Name.NormD21 | 0.62713 |
| 437 | 0.01278 | 0.01287 | 68 | 0.99296 | 0.32425 | Intestinibacter.sp..MP113D11 | Visit.Name.NormD21 | 0.62713 |
| 438 | 0.05458 | 0.06419 | 68 | 0.85032 | 0.39813 | Bacteroides.sp..VE800.11 | Visit.Name.NormD3 | 0.62838 |
| 439 | 0.02715 | 0.02757 | 68 | 0.98482 | 0.32820 | Alistipes.sp..MP104G4_1 | Visit.Name.NormD14 | 0.62866 |
| 440 | 0.02351 | 0.02367 | 68 | 0.99333 | 0.32407 | Alistipes.finegoldii | Visit.Name.NormD14 | 0.62866 |
| 441 | 0.01338 | 0.01289 | 68 | 1.03822 | 0.30285 | Klebsiella.oxytoca | Visit.Name.NormD14 | 0.62866 |
| 442 | -0.01328 | 0.01350 | 68 | -0.98398 | 0.32861 | Bacteroides.dorei | Visit.Name.NormD14 | 0.62866 |
| 443 | 0.01235 | 0.01256 | 68 | 0.98331 | 0.32894 | Ruminococcus.bicirculans | Visit.Name.NormD14 | 0.62866 |
| 444 | -0.01373 | 0.01369 | 68 | -1.00250 | 0.31966 | Parvimonas.micra | Visit.Name.NormD14 | 0.62866 |
| 445 | -0.01400 | 0.01371 | 68 | -1.02140 | 0.31069 | Enterococcus.faecalis | Visit.Name.NormD14 | 0.62866 |
| 446 | 0.01635 | 0.01527 | 68 | 1.07067 | 0.28811 | Shigella.dysenteriae | Visit.Name.NormD14 | 0.62866 |
| 447 | -0.00583 | 0.00595 | 68 | -0.97936 | 0.33087 | Sutterella.sp..54_7 | Visit.Name.NormD14 | 0.62866 |
| 448 | 0.01193 | 0.01219 | 68 | 0.97884 | 0.33113 | Prevotella.copri | Visit.Name.NormD14 | 0.62866 |

| | | | | | | | | |
|---|---|---|---|---|---|---|---|---|
| 449 | 0.01155 | 0.01082 | 68 | 1.06720 | 0.28965 | Oscillospiraceae.bacterium.VE202.24 | Visit.Name.NormD30 | 0.63241 |
| 450 | -0.01751 | 0.01632 | 68 | -1.07299 | 0.28707 | Blautia.sp..HK34 | Visit.Name.NormD30 | 0.63241 |
| 451 | -0.01518 | 0.01383 | 68 | -1.09713 | 0.27645 | Phascolarctobacterium.sp..MUR_MP6E9_ig1 | Visit.Name.NormD30 | 0.63241 |
| 452 | 0.02615 | 0.02362 | 68 | 1.10701 | 0.27219 | Bacteroides.sp..MP163B7 | Visit.Name.NormD30 | 0.63241 |
| 453 | -0.01942 | 0.01762 | 68 | -1.10219 | 0.27426 | Solobacterium.moorei | Visit.Name.NormD30 | 0.63241 |
| 454 | 0.01593 | 0.01435 | 68 | 1.11028 | 0.27079 | Parvimonas.micra | Visit.Name.NormD30 | 0.63241 |
| 455 | 0.01847 | 0.01703 | 68 | 1.08476 | 0.28186 | Bacteroides.sp..MP227D2 | Visit.Name.NormD30 | 0.63241 |
| 456 | 0.01008 | 0.01209 | 68 | 0.83370 | 0.40737 | Streptococcus.sp..MP111E2 | Visit.Name.NormD7 | 0.63531 |
| 457 | 0.01240 | 0.01295 | 68 | 0.95751 | 0.34170 | Bacteroides.sp..MP369D12 | Visit.Name.NormD14 | 0.63947 |
| 458 | 0.02567 | 0.01921 | 68 | 1.33631 | 0.18590 | Acidaminococcus.intestini | Visit.Name.NormD58 | 0.64087 |
| 459 | -0.01202 | 0.01472 | 68 | -0.81612 | 0.41728 | Parvimonas.micra | Visit.Name.NormD7 | 0.64310 |
| 460 | 0.04987 | 0.06199 | 68 | 0.80443 | 0.42395 | Klebsiella.pneumoniae | Visit.Name.NormD7 | 0.64579 |
| 461 | -0.01167 | 0.01436 | 68 | -0.81310 | 0.41900 | Parvimonas.micra | Visit.Name.NormD3 | 0.65344 |
| 462 | -0.00984 | 0.01252 | 68 | -0.78578 | 0.43473 | Blautia.sp..MP351A5 | Visit.Name.NormD7 | 0.65459 |
| 463 | 0.01744 | 0.01336 | 68 | 1.30556 | 0.19610 | Absiella.sp..MP634G1_ig1 | Visit.Name.NormD58 | 0.65791 |
| 464 | -0.02273 | 0.01760 | 68 | -1.29154 | 0.20089 | Blautia.sp..MP512E3 | Visit.Name.NormD58 | 0.65791 |
| 465 | 0.01312 | 0.01307 | 68 | 1.00375 | 0.31906 | Roseburia.hominis | Visit.Name.NormD30 | 0.66076 |
| 466 | -0.05449 | 0.05646 | 68 | -0.96504 | 0.33795 | Escherichia.coli | Visit.Name.NormD30 | 0.66076 |
| 467 | -0.01172 | 0.01152 | 68 | -1.01795 | 0.31231 | X.Clostridium..citroniae | Visit.Name.NormD30 | 0.66076 |
| 468 | -0.02138 | 0.02214 | 68 | -0.96590 | 0.33752 | Eubacterium.sp..CAG.180 | Visit.Name.NormD30 | 0.66076 |
| 469 | 0.02716 | 0.02683 | 68 | 1.01230 | 0.31498 | Bacteroides.sp..MP487B8 | Visit.Name.NormD30 | 0.66076 |
| 470 | 0.01112 | 0.01138 | 68 | 0.97754 | 0.33177 | Pseudomonas.aeruginosa | Visit.Name.NormD30 | 0.66076 |
| 471 | -0.01429 | 0.01436 | 68 | -0.99503 | 0.32325 | Pediococcus.acidilactici | Visit.Name.NormD30 | 0.66076 |
| 472 | 0.01309 | 0.01417 | 68 | 0.92345 | 0.35904 | Bacteroides.sp..MP327C3 | Visit.Name.NormD21 | 0.66381 |
| 473 | -0.02944 | 0.03193 | 68 | -0.92202 | 0.35978 | Lachnospiraceae.bacterium.1_1_57FAA | Visit.Name.NormD21 | 0.66381 |
| 474 | 0.01029 | 0.01126 | 68 | 0.91362 | 0.36415 | Streptococcus.sp..MP111E2 | Visit.Name.NormD14 | 0.67187 |
| 475 | 0.01153 | 0.01222 | 68 | 0.94337 | 0.34883 | Blautia.sp..MP351A5 | Visit.Name.NormD30 | 0.67201 |
| 476 | 0.00902 | 0.01208 | 68 | 0.74628 | 0.45807 | Parasutterella.excrementihominis | Visit.Name.NormD7 | 0.68190 |
| 477 | -0.01166 | 0.01321 | 68 | -0.88279 | 0.38046 | Phascolarctobacterium.sp..MUR_MP6E9_ig1 | Visit.Name.NormD14 | 0.68275 |
| 478 | -0.01876 | 0.02118 | 68 | -0.88575 | 0.37887 | Eubacterium.sp..CAG.180 | Visit.Name.NormD14 | 0.68275 |
| 479 | 0.01379 | 0.01289 | 68 | 1.06908 | 0.28882 | Parasutterella.excrementihominis | Visit.Name.NormD1 | 0.68791 |
| 480 | -0.02706 | 0.02166 | 68 | -1.24941 | 0.21580 | Colibacter.massiliensis | Visit.Name.NormD58 | 0.68950 |
| 481 | 0.02640 | 0.02888 | 68 | 0.91419 | 0.36385 | Alistipes.sp..MP104G4_1 | Visit.Name.NormD30 | 0.69078 |
| 482 | 0.01401 | 0.01628 | 68 | 0.86050 | 0.39254 | Bacteroides.sp..MP227D2 | Visit.Name.NormD14 | 0.69490 |
| 483 | -0.02698 | 0.02223 | 68 | -1.21350 | 0.22914 | Solobacterium.moorei | Visit.Name.NormD58 | 0.69808 |
| 484 | -0.02103 | 0.01733 | 68 | -1.21349 | 0.22914 | Desulfovibrio.desulfuricans | Visit.Name.NormD58 | 0.69808 |
| 485 | -0.04171 | 0.04762 | 68 | -0.87603 | 0.38410 | Lactobacillus.rhamnosus | Visit.Name.NormD21 | 0.69884 |
| 486 | -0.02533 | 0.02116 | 68 | -1.19698 | 0.23547 | Sutterella.wadsworthensis | Visit.Name.NormD58 | 0.70106 |
| 487 | -0.01184 | 0.01409 | 68 | -0.84017 | 0.40376 | Blautia.sp..MP87G3 | Visit.Name.NormD14 | 0.70523 |
| 488 | -0.01147 | 0.01480 | 68 | -0.77481 | 0.44114 | Blautia.sp..HF17 | Visit.Name.NormD21 | 0.70798 |
| 489 | -0.01572 | 0.01908 | 68 | -0.82377 | 0.41295 | Lachnospiraceae.bacterium.7_1_58FAA | Visit.Name.NormD21 | 0.70798 |
| 490 | 0.01236 | 0.01544 | 68 | 0.80092 | 0.42597 | Erysipelotrichaceae.bacterium.6_1_45 | Visit.Name.NormD21 | 0.70798 |
| 491 | -0.01712 | 0.02196 | 68 | -0.77968 | 0.43828 | Bacteroides.vulgatus | Visit.Name.NormD21 | 0.70798 |
| 492 | -0.04624 | 0.06037 | 68 | -0.76595 | 0.44636 | Klebsiella.pneumoniae | Visit.Name.NormD21 | 0.70798 |
| 493 | -0.01178 | 0.01395 | 68 | -0.84433 | 0.40145 | Blautia.sp..MP40F7 | Visit.Name.NormD21 | 0.70798 |
| 494 | -0.01361 | 0.01767 | 68 | -0.77027 | 0.44381 | Veillonella.parvula | Visit.Name.NormD21 | 0.70798 |
| 495 | 0.00898 | 0.01151 | 68 | 0.77997 | 0.43811 | X.Clostridium..citroniae | Visit.Name.NormD21 | 0.70798 |
| 496 | -0.01195 | 0.01568 | 68 | -0.76221 | 0.44857 | Eisenbergiella.sp..OF01.20 | Visit.Name.NormD21 | 0.70798 |
| 497 | -0.03001 | 0.03805 | 68 | -0.78862 | 0.43308 | Bacteroides.fragilis | Visit.Name.NormD21 | 0.70798 |
| 498 | 0.01506 | 0.01936 | 68 | 0.77772 | 0.43943 | Acidaminococcus.sp..MP76F2 | Visit.Name.NormD21 | 0.70798 |
| 499 | -0.00985 | 0.01324 | 68 | -0.74375 | 0.45959 | Blautia.coccoides | Visit.Name.NormD3 | 0.70831 |
| 500 | -0.01153 | 0.01571 | 68 | -0.73374 | 0.46563 | Eisenbergiella.sp..OF01.20 | Visit.Name.NormD3 | 0.70927 |
| 501 | -0.03757 | 0.03625 | 68 | -1.03627 | 0.30375 | Bacteroides.ovatus | Visit.Name.NormD1 | 0.71055 |
| 502 | -0.01870 | 0.01830 | 68 | -1.02216 | 0.31033 | Clostridioides.difficile | Visit.Name.NormD1 | 0.71321 |
| 503 | -0.00700 | 0.00998 | 68 | -0.70155 | 0.48535 | Neglecta.timonensis | Visit.Name.NormD7 | 0.71439 |
| 504 | -0.01626 | 0.02267 | 68 | -0.71722 | 0.47570 | Ruthenibacterium.sp..VE800.03 | Visit.Name.NormD3 | 0.71628 |
| 505 | 0.00624 | 0.00537 | 68 | 1.16346 | 0.24871 | Phascolarctobacterium.succinatutens | Visit.Name.NormD58 | 0.72402 |
| 506 | -0.05257 | 0.06042 | 68 | -0.87012 | 0.38730 | Klebsiella.pneumoniae | Visit.Name.NormD30 | 0.72480 |
| 507 | -0.01624 | 0.02212 | 68 | -0.73431 | 0.46529 | Eubacterium.sp..CAG.180 | Visit.Name.NormD21 | 0.72563 |
| 508 | -0.00732 | 0.01094 | 68 | -0.66876 | 0.50592 | Clostridiales.bacterium | Visit.Name.NormD7 | 0.73639 |
| 509 | -0.03973 | 0.04764 | 68 | -0.83397 | 0.40722 | Lactobacillus.rhamnosus | Visit.Name.NormD30 | 0.74091 |
| 510 | 0.00920 | 0.01101 | 68 | 0.83550 | 0.40637 | X.Clostridium..leptum | Visit.Name.NormD30 | 0.74091 |
| 511 | -0.02316 | 0.02042 | 68 | -1.13406 | 0.26075 | Blautia.sp..HK34 | Visit.Name.NormD58 | 0.74258 |
| 512 | 0.01084 | 0.01116 | 68 | 0.97171 | 0.33464 | Streptococcus.thermophilus | Visit.Name.NormD1 | 0.74302 |
| 513 | 0.01675 | 0.01720 | 68 | 0.97347 | 0.33377 | Eisenbergiella.sp..OF01.20 | Visit.Name.NormD1 | 0.74302 |
| 514 | -0.02738 | 0.03882 | 68 | -0.70549 | 0.48292 | Pyramidobacter.piscolens | Visit.Name.NormD21 | 0.74426 |
| 515 | -0.03070 | 0.03810 | 68 | -0.80580 | 0.42317 | Bacteroides.fragilis | Visit.Name.NormD30 | 0.74912 |
| 516 | 0.01064 | 0.01311 | 68 | 0.81188 | 0.41969 | Flavobacteriales.bacterium | Visit.Name.NormD30 | 0.74912 |
| 517 | -0.01266 | 0.01329 | 68 | -0.95216 | 0.34439 | Blautia.sp..MP351A5 | Visit.Name.NormD1 | 0.75192 |
| 518 | 0.00794 | 0.01177 | 68 | 0.67489 | 0.50204 | Streptococcus.sp..MP111E2 | Visit.Name.NormD21 | 0.75594 |
| 519 | 0.00820 | 0.01204 | 68 | 0.68131 | 0.49799 | Gemella.bergeri | Visit.Name.NormD21 | 0.75594 |
| 520 | -0.01253 | 0.01888 | 68 | -0.66395 | 0.50897 | Alistipes.sp..HK66 | Visit.Name.NormD3 | 0.75767 |
| 521 | 0.00687 | 0.00886 | 68 | 0.77531 | 0.44084 | X.Clostridium..aldenense | Visit.Name.NormD14 | 0.75987 |
| 522 | -0.03034 | 0.03884 | 68 | -0.78122 | 0.43738 | Pyramidobacter.piscolens | Visit.Name.NormD30 | 0.76396 |
| 523 | 0.01044 | 0.01354 | 68 | 0.77120 | 0.44326 | Bacteroides.sp..MP369D12 | Visit.Name.NormD30 | 0.76404 |

| | | | | | | | | |
|---|---|---|---|---|---|---|---|---|
| 524 | 0.00784 | 0.01214 | 68 | 0.64580 | 0.52058 | Methanobrevibacter.smithii | Visit.Name.NormD21 | 0.76625 |
| 525 | 0.02466 | 0.03787 | 68 | 0.65105 | 0.51721 | Akkermansia.muciniphila | Visit.Name.NormD21 | 0.76625 |
| 526 | -0.00840 | 0.01316 | 68 | -0.63834 | 0.52539 | Ruminococcus.bicirculans | Visit.Name.NormD3 | 0.77333 |
| 527 | -0.04901 | 0.04491 | 68 | -1.09142 | 0.27894 | Bacteroides.thetaiotaomicron | Visit.Name.NormD58 | 0.77748 |
| 528 | -0.00840 | 0.01364 | 68 | -0.61558 | 0.54023 | Bacteroides.ovatus.CAG.22 | Visit.Name.NormD7 | 0.77769 |
| 529 | 0.01330 | 0.02256 | 68 | 0.58945 | 0.55752 | Lactobacillus.fermentum | Visit.Name.NormD7 | 0.78073 |
| 530 | 0.00697 | 0.01248 | 68 | 0.55827 | 0.57849 | Methanobrevibacter.smithii | Visit.Name.NormD7 | 0.78073 |
| 531 | 0.01080 | 0.01956 | 68 | 0.55192 | 0.58281 | Lachnospiraceae.bacterium.7_1_58FAA | Visit.Name.NormD7 | 0.78073 |
| 532 | -0.01605 | 0.02964 | 68 | -0.54138 | 0.59002 | Alistipes.sp..MP104G4_1 | Visit.Name.NormD7 | 0.78073 |
| 533 | -0.00641 | 0.01084 | 68 | -0.59121 | 0.55634 | Absiella.sp..MP634G1_ig1 | Visit.Name.NormD7 | 0.78073 |
| 534 | 0.01024 | 0.01814 | 68 | 0.56473 | 0.57411 | Veillonella.parvula | Visit.Name.NormD7 | 0.78073 |
| 535 | -0.00907 | 0.01611 | 68 | -0.56305 | 0.57525 | Eisenbergiella.sp..OF01.20 | Visit.Name.NormD7 | 0.78073 |
| 536 | 0.03388 | 0.06210 | 68 | 0.54567 | 0.58708 | Klebsiella.michiganensis | Visit.Name.NormD7 | 0.78073 |
| 537 | -0.01261 | 0.01722 | 68 | -0.73230 | 0.46650 | Colibacter.massiliensis | Visit.Name.NormD30 | 0.78349 |
| 538 | -0.01419 | 0.01912 | 68 | -0.74200 | 0.46064 | Bacteroides.sp..MP264A7 | Visit.Name.NormD30 | 0.78349 |
| 539 | -0.00594 | 0.01154 | 68 | -0.51511 | 0.60814 | Blautia.sp..VE303.05 | Visit.Name.NormD7 | 0.78617 |
| 540 | 0.01352 | 0.02793 | 68 | 0.48430 | 0.62973 | Alistipes.sp..AF14.19 | Visit.Name.NormD7 | 0.78617 |
| 541 | -0.00684 | 0.01455 | 68 | -0.47031 | 0.63964 | Bacteroides.sp..MP327C3 | Visit.Name.NormD7 | 0.78617 |
| 542 | -0.00921 | 0.01963 | 68 | -0.46942 | 0.64027 | Bacteroides.sp..MP264A7 | Visit.Name.NormD7 | 0.78617 |
| 543 | -0.00859 | 0.01716 | 68 | -0.50038 | 0.61842 | Sutterella.wadsworthensis | Visit.Name.NormD7 | 0.78617 |
| 544 | -0.02276 | 0.04875 | 68 | -0.46679 | 0.64214 | Lactobacillus.plantarum | Visit.Name.NormD7 | 0.78617 |
| 545 | 0.00705 | 0.01406 | 68 | 0.50105 | 0.61795 | Desulfovibrio.desulfuricans | Visit.Name.NormD7 | 0.78617 |
| 546 | -0.00761 | 0.01533 | 68 | -0.49674 | 0.62097 | Atopobium.minutum | Visit.Name.NormD7 | 0.78617 |
| 547 | 0.00808 | 0.01125 | 68 | 0.71822 | 0.47508 | Lachnospiraceae.bacterium.5_1_57FAA | Visit.Name.NormD30 | 0.78780 |
| 548 | -0.00710 | 0.01567 | 68 | -0.45282 | 0.65212 | Acidaminococcus.intestini | Visit.Name.NormD7 | 0.79100 |
| 549 | 0.00587 | 0.01321 | 68 | 0.44416 | 0.65833 | Intestinibacter.sp..MP113D11 | Visit.Name.NormD7 | 0.79121 |
| 550 | -0.00812 | 0.01332 | 68 | -0.60947 | 0.54424 | Bacteroides.ovatus.CAG.22 | Visit.Name.NormD3 | 0.79218 |
| 551 | 0.01051 | 0.02424 | 68 | 0.43349 | 0.66603 | X.Clostridium..clostridioforme | Visit.Name.NormD7 | 0.79318 |
| 552 | -0.00636 | 0.01067 | 68 | -0.59626 | 0.55298 | Clostridiales.bacterium | Visit.Name.NormD3 | 0.79605 |
| 553 | -0.00717 | 0.01222 | 68 | -0.58692 | 0.55920 | Blautia.sp..MP351A5 | Visit.Name.NormD3 | 0.79626 |
| 554 | 0.00746 | 0.01066 | 68 | 0.69981 | 0.48643 | Clostridiales.bacterium | Visit.Name.NormD30 | 0.79653 |
| 555 | -0.00817 | 0.01951 | 68 | -0.41843 | 0.67695 | Bifidobacterium.longum | Visit.Name.NormD7 | 0.79893 |
| 556 | -0.01482 | 0.01670 | 68 | -0.88758 | 0.37790 | Acidaminococcus.intestini | Visit.Name.NormD1 | 0.80102 |
| 557 | 0.01330 | 0.01503 | 68 | 0.88531 | 0.37911 | Desulfovibrio.desulfuricans | Visit.Name.NormD1 | 0.80102 |
| 558 | 0.03632 | 0.04158 | 68 | 0.87351 | 0.38546 | Akkermansia.muciniphila | Visit.Name.NormD1 | 0.80151 |
| 559 | -0.02591 | 0.02451 | 68 | -1.05683 | 0.29433 | Acidaminococcus.sp..MP76F2 | Visit.Name.NormD58 | 0.80327 |
| 560 | -0.00454 | 0.01168 | 68 | -0.38881 | 0.69863 | Dorea.sp..MP50E3 | Visit.Name.NormD7 | 0.80992 |
| 561 | 0.00980 | 0.02472 | 68 | 0.39627 | 0.69315 | X.Ruminococcus..sp..MP495E3 | Visit.Name.NormD7 | 0.80992 |
| 562 | 0.01654 | 0.02482 | 68 | 0.66623 | 0.50752 | Alistipes.finegoldii | Visit.Name.NormD30 | 0.81079 |
| 563 | 0.00951 | 0.01413 | 68 | 0.67343 | 0.50296 | Bacteroides.dorei | Visit.Name.NormD30 | 0.81079 |
| 564 | -0.01096 | 0.01678 | 68 | -0.65281 | 0.51608 | Clostridioides.difficile | Visit.Name.NormD30 | 0.81211 |
| 565 | -0.01131 | 0.01754 | 68 | -0.64456 | 0.52138 | Bacteroides.xylanisolvens | Visit.Name.NormD30 | 0.81211 |
| 566 | -0.01535 | 0.02414 | 68 | -0.63596 | 0.52694 | Citrobacter.freundii | Visit.Name.NormD30 | 0.81211 |
| 567 | -0.00850 | 0.01395 | 68 | -0.60899 | 0.54456 | Blautia.sp..MP40F7 | Visit.Name.NormD30 | 0.81309 |
| 568 | -0.01094 | 0.01768 | 68 | -0.61867 | 0.53820 | Veillonella.parvula | Visit.Name.NormD30 | 0.81309 |
| 569 | 0.00860 | 0.01418 | 68 | 0.60651 | 0.54620 | Bacteroides.sp..MP327C3 | Visit.Name.NormD30 | 0.81309 |
| 570 | -0.04004 | 0.05775 | 68 | -0.69331 | 0.49048 | Klebsiella.pneumoniae | Visit.Name.NormD14 | 0.81332 |
| 571 | 0.01628 | 0.02307 | 68 | 0.70600 | 0.48260 | X.Ruminococcus..sp..MP495E3 | Visit.Name.NormD14 | 0.81332 |
| 572 | -0.00800 | 0.01144 | 68 | -0.69978 | 0.48645 | Ruminococcus.sp..MP226F3 | Visit.Name.NormD14 | 0.81332 |
| 573 | 0.00612 | 0.01043 | 68 | 0.58700 | 0.55915 | Eubacterium.sp..CAG.38 | Visit.Name.NormD21 | 0.81387 |
| 574 | -0.02287 | 0.02723 | 68 | -0.83985 | 0.40393 | Alistipes.finegoldii | Visit.Name.NormD1 | 0.81457 |
| 575 | -0.01255 | 0.01515 | 68 | -0.82831 | 0.41039 | Phascolarctobacterium.sp..MUR_MP6E9_ig1 | Visit.Name.NormD1 | 0.81457 |
| 576 | 0.01041 | 0.01248 | 68 | 0.83460 | 0.40687 | Pseudomonas.aeruginosa | Visit.Name.NormD1 | 0.81457 |
| 577 | 0.00758 | 0.01288 | 68 | 0.58867 | 0.55803 | Intestinibacter.sp..MP113D11 | Visit.Name.NormD30 | 0.82137 |
| 578 | 0.00377 | 0.00465 | 68 | 0.80987 | 0.42084 | Phascolarctobacterium.succinatutens | Visit.Name.NormD1 | 0.82284 |
| 579 | -0.02332 | 0.02271 | 68 | -1.02681 | 0.30815 | Blautia.sp..MP588D7 | Visit.Name.NormD58 | 0.82383 |
| 580 | -0.00950 | 0.01232 | 68 | -0.77056 | 0.44363 | Lachnospiraceae.bacterium.5_1_57FAA | Visit.Name.NormD1 | 0.82424 |
| 581 | -0.00879 | 0.01137 | 68 | -0.77296 | 0.44222 | Eubacterium.sp..CAG.38 | Visit.Name.NormD1 | 0.82424 |
| 582 | -0.01101 | 0.01439 | 68 | -0.76533 | 0.44673 | Ruminococcus.bicirculans | Visit.Name.NormD1 | 0.82424 |
| 583 | 0.01041 | 0.01321 | 68 | 0.78778 | 0.43356 | Gemella.bergeri | Visit.Name.NormD1 | 0.82424 |
| 584 | -0.01753 | 0.02379 | 68 | -0.73695 | 0.46369 | Bacteroides.vulgatus | Visit.Name.NormD1 | 0.82540 |
| 585 | -0.01152 | 0.01572 | 68 | -0.73271 | 0.46625 | Parvimonas.micra | Visit.Name.NormD1 | 0.82540 |
| 586 | 0.01575 | 0.02126 | 68 | 0.74110 | 0.46119 | Acidaminococcus.sp..MP76F2 | Visit.Name.NormD1 | 0.82540 |
| 587 | 0.01105 | 0.01647 | 68 | 0.67112 | 0.50442 | Colibacter.massiliensis | Visit.Name.NormD14 | 0.82598 |
| 588 | 0.00800 | 0.01128 | 68 | 0.70947 | 0.48046 | Lactobacillus.paracasei | Visit.Name.NormD1 | 0.82727 |
| 589 | -0.00878 | 0.01254 | 68 | -0.70009 | 0.48626 | X.Clostridium..citroniae | Visit.Name.NormD1 | 0.82727 |
| 590 | -0.00996 | 0.01407 | 68 | -0.70790 | 0.48143 | Intestinibacter.sp..MP113D11 | Visit.Name.NormD1 | 0.82727 |
| 591 | 0.00843 | 0.01557 | 68 | 0.54098 | 0.59029 | Bilophila.wadsworthia | Visit.Name.NormD21 | 0.82893 |
| 592 | -0.00996 | 0.01789 | 68 | -0.55705 | 0.57932 | Veillonella.atypica | Visit.Name.NormD21 | 0.82893 |
| 593 | -0.01006 | 0.01883 | 68 | -0.53441 | 0.59480 | Alistipes.sp..HK66 | Visit.Name.NormD21 | 0.82893 |
| 594 | -0.00781 | 0.01432 | 68 | -0.54511 | 0.58746 | Acidaminococcus.intestini.CAG.325 | Visit.Name.NormD21 | 0.82893 |
| 595 | -0.00612 | 0.01126 | 68 | -0.54362 | 0.58848 | Blautia.sp..VE303.05 | Visit.Name.NormD3 | 0.82894 |
| 596 | 0.00810 | 0.01187 | 68 | 0.68211 | 0.49749 | Desulfovibrio.fairfieldensis | Visit.Name.NormD1 | 0.83553 |
| 597 | -0.00981 | 0.01619 | 68 | -0.60566 | 0.54675 | Blautia.sp..HF17 | Visit.Name.NormD1 | 0.83807 |
| 598 | -0.01852 | 0.03165 | 68 | -0.58499 | 0.56049 | Alistipes.sp..MP104G4_1 | Visit.Name.NormD1 | 0.83807 |

| | | | | | | | | |
|---|---|---|---|---|---|---|---|---|
| 599 | -0.00772 | 0.01204 | 68 | -0.64113 | 0.52359 | X.Clostridium..leptum | Visit.Name.NormD1 | 0.83807 |
| 600 | 0.00872 | 0.01576 | 68 | 0.55286 | 0.58217 | X.Clostridium..symbiosum | Visit.Name.NormD1 | 0.83807 |
| 601 | -0.01068 | 0.01780 | 68 | -0.60034 | 0.55028 | Blautia.sp..HK34 | Visit.Name.NormD1 | 0.83807 |
| 602 | 0.00744 | 0.01289 | 68 | 0.57777 | 0.56533 | Streptococcus.sp..MP111E2 | Visit.Name.NormD1 | 0.83807 |
| 603 | -0.00892 | 0.01427 | 68 | -0.62510 | 0.53400 | Barnesiella.intestinihominis | Visit.Name.NormD1 | 0.83807 |
| 604 | -0.01473 | 0.02632 | 68 | -0.55958 | 0.57761 | X.Ruminococcus..sp..MP495E3 | Visit.Name.NormD1 | 0.83807 |
| 605 | -0.00876 | 0.01551 | 68 | -0.56463 | 0.57419 | Bacteroides.sp..MP327C3 | Visit.Name.NormD1 | 0.83807 |
| 606 | 0.01582 | 0.02592 | 68 | 0.61029 | 0.54371 | Bacteroides.sp..MP163B7 | Visit.Name.NormD1 | 0.83807 |
| 607 | -0.00949 | 0.01568 | 68 | -0.60514 | 0.54710 | Pediococcus.acidilactici | Visit.Name.NormD1 | 0.83807 |
| 608 | 0.00995 | 0.01748 | 68 | 0.56926 | 0.57106 | Shigella.dysenteriae | Visit.Name.NormD1 | 0.83807 |
| 609 | 0.00837 | 0.01393 | 68 | 0.60108 | 0.54979 | Enterobacteriaceae.bacterium.9_2_54FAA | Visit.Name.NormD1 | 0.83807 |
| 610 | 0.00820 | 0.01279 | 68 | 0.64121 | 0.52354 | Bacteroides.ovatus.CAG.22 | Visit.Name.NormD14 | 0.84672 |
| 611 | 0.00443 | 0.00876 | 68 | 0.50629 | 0.61429 | Megasphaera.sp..MP182G6 | Visit.Name.NormD21 | 0.84707 |
| 612 | -0.00948 | 0.01910 | 68 | -0.49619 | 0.62136 | Bacteroides.sp..MP264A7 | Visit.Name.NormD21 | 0.84790 |
| 613 | 0.00998 | 0.02028 | 68 | 0.49192 | 0.62436 | Cloacibacillus.evryensis | Visit.Name.NormD3 | 0.85167 |
| 614 | 0.01148 | 0.02364 | 68 | 0.48551 | 0.62888 | X.Clostridium..clostridioforme | Visit.Name.NormD3 | 0.85167 |
| 615 | -0.00384 | 0.00787 | 68 | -0.48753 | 0.62745 | Sutterella.sp..AM11.39 | Visit.Name.NormD3 | 0.85167 |
| 616 | -0.00617 | 0.01276 | 68 | -0.48303 | 0.63063 | Enterobacteriaceae.bacterium.9_2_54FAA | Visit.Name.NormD3 | 0.85167 |
| 617 | -0.00514 | 0.01114 | 68 | -0.46101 | 0.64626 | Veillonella.sp..MP19D1_02.09 | Visit.Name.NormD21 | 0.85262 |
| 618 | -0.01075 | 0.02409 | 68 | -0.44623 | 0.65685 | X.Ruminococcus..sp..MP495E3 | Visit.Name.NormD21 | 0.85262 |
| 619 | -0.00767 | 0.01721 | 68 | -0.44552 | 0.65736 | Colibacter.massiliensis | Visit.Name.NormD21 | 0.85262 |
| 620 | -0.00745 | 0.01670 | 68 | -0.44633 | 0.65677 | Sutterella.wadsworthensis | Visit.Name.NormD21 | 0.85262 |
| 621 | -0.00645 | 0.01433 | 68 | -0.44999 | 0.65415 | Parvimonas.micra | Visit.Name.NormD21 | 0.85262 |
| 622 | 0.01385 | 0.03061 | 68 | 0.45261 | 0.65227 | Parabacteroides.sp..VE800.10 | Visit.Name.NormD3 | 0.85284 |
| 623 | 0.00526 | 0.01179 | 68 | 0.44656 | 0.65661 | Streptococcus.sp..MP111E2 | Visit.Name.NormD3 | 0.85284 |
| 624 | 0.01151 | 0.02658 | 68 | 0.43305 | 0.66635 | Raoultella.ornithinolytica | Visit.Name.NormD3 | 0.85284 |
| 625 | -0.00606 | 0.01419 | 68 | -0.42724 | 0.67055 | Bacteroides.sp..MP327C3 | Visit.Name.NormD3 | 0.85284 |
| 626 | 0.00689 | 0.01598 | 68 | 0.43134 | 0.66758 | Shigella.dysenteriae | Visit.Name.NormD3 | 0.85284 |
| 627 | -0.00663 | 0.01495 | 68 | -0.44324 | 0.65900 | Atopobium.minutum | Visit.Name.NormD3 | 0.85284 |
| 628 | 0.01286 | 0.02410 | 68 | 0.53378 | 0.59523 | X.Ruminococcus..sp..MP495E3 | Visit.Name.NormD30 | 0.85334 |
| 629 | 0.00479 | 0.00877 | 68 | 0.54560 | 0.58713 | Megasphaera.sp..MP182G6 | Visit.Name.NormD30 | 0.85334 |
| 630 | -0.00757 | 0.01433 | 68 | -0.52789 | 0.59929 | Acidaminococcus.intestini.CAG.325 | Visit.Name.NormD30 | 0.85334 |
| 631 | -0.00742 | 0.01431 | 68 | -0.51839 | 0.60587 | Roseburia.hominis | Visit.Name.NormD1 | 0.85343 |
| 632 | 0.00993 | 0.01883 | 68 | 0.52720 | 0.59977 | Colibacter.massiliensis | Visit.Name.NormD1 | 0.85343 |
| 633 | -0.02491 | 0.04909 | 68 | -0.50739 | 0.61353 | Bacteroides.sp..VE800.09 | Visit.Name.NormD1 | 0.85502 |
| 634 | 0.01574 | 0.02607 | 68 | 0.60387 | 0.54794 | Alistipes.sp..AF14.19 | Visit.Name.NormD14 | 0.85548 |
| 635 | 0.00459 | 0.00765 | 68 | 0.60016 | 0.55039 | Megasphaera.massiliensis | Visit.Name.NormD14 | 0.85548 |
| 636 | 0.00644 | 0.01086 | 68 | 0.59310 | 0.55508 | Pseudomonas.aeruginosa | Visit.Name.NormD14 | 0.85548 |
| 637 | -0.02294 | 0.03715 | 68 | -0.61748 | 0.53898 | Pyramidobacter.piscolens | Visit.Name.NormD14 | 0.85548 |
| 638 | -0.02111 | 0.04246 | 68 | -0.49716 | 0.62068 | Pyramidobacter.piscolens | Visit.Name.NormD1 | 0.85588 |
| 639 | -0.00819 | 0.01597 | 68 | -0.51275 | 0.60979 | Shigella.dysenteriae | Visit.Name.NormD30 | 0.85895 |
| 640 | 0.04064 | 0.04152 | 68 | 0.97885 | 0.33113 | Bacteroides.ovatus | Visit.Name.NormD58 | 0.86755 |
| 641 | 0.00612 | 0.01487 | 68 | 0.41169 | 0.68186 | Eubacterium.sp..VE800.02 | Visit.Name.NormD1 | 0.86826 |
| 642 | 0.00737 | 0.01765 | 68 | 0.41760 | 0.67756 | X.Ruminococcus..torques.CAG.61 | Visit.Name.NormD1 | 0.86826 |
| 643 | 0.00625 | 0.01447 | 68 | 0.43219 | 0.66697 | Bacteroides.ovatus.CAG.22 | Visit.Name.NormD1 | 0.86826 |
| 644 | -0.00708 | 0.01537 | 68 | -0.46052 | 0.64662 | X.Ruminococcus..sp..MP50A2 | Visit.Name.NormD1 | 0.86826 |
| 645 | 0.02253 | 0.05189 | 68 | 0.43423 | 0.66549 | Lactobacillus.plantarum | Visit.Name.NormD1 | 0.86826 |
| 646 | -0.00963 | 0.02068 | 68 | -0.46569 | 0.64292 | Alistipes.sp..HK66 | Visit.Name.NormD1 | 0.86826 |
| 647 | 0.00281 | 0.00685 | 68 | 0.41057 | 0.68268 | Sutterella.sp..54_7 | Visit.Name.NormD1 | 0.86826 |
| 648 | 0.00673 | 0.01637 | 68 | 0.41114 | 0.68227 | Atopobium.minutum | Visit.Name.NormD1 | 0.86826 |
| 649 | -0.00932 | 0.01908 | 68 | -0.48847 | 0.62679 | Lachnospiraceae.bacterium.7_1_58FAA | Visit.Name.NormD30 | 0.86894 |
| 650 | -0.00633 | 0.01309 | 68 | -0.48371 | 0.63015 | Barnesiella.intestinihominis | Visit.Name.NormD30 | 0.86894 |
| 651 | -0.00455 | 0.01139 | 68 | -0.39954 | 0.69075 | Dorea.sp..MP50E3 | Visit.Name.NormD3 | 0.87008 |
| 652 | 0.00656 | 0.01544 | 68 | 0.42450 | 0.67254 | Erysipelotrichaceae.bacterium.6_1_45 | Visit.Name.NormD30 | 0.87230 |
| 653 | -0.00539 | 0.01264 | 68 | -0.42661 | 0.67101 | HI22.sp..HI22 | Visit.Name.NormD30 | 0.87230 |
| 654 | 0.00654 | 0.01469 | 68 | 0.44493 | 0.65778 | Blautia.sp..MP87G3 | Visit.Name.NormD30 | 0.87230 |
| 655 | 0.00299 | 0.00689 | 68 | 0.43388 | 0.66575 | Megasphaera.sp..UBA1696 | Visit.Name.NormD30 | 0.87230 |
| 656 | -0.00808 | 0.01886 | 68 | -0.42827 | 0.66981 | Alistipes.sp..HK66 | Visit.Name.NormD30 | 0.87230 |
| 657 | -0.00617 | 0.01370 | 68 | -0.45007 | 0.65409 | Desulfovibrio.desulfuricans | Visit.Name.NormD30 | 0.87230 |
| 658 | -0.00627 | 0.01123 | 68 | -0.55855 | 0.57831 | Alistipes.putredinis | Visit.Name.NormD14 | 0.87253 |
| 659 | 0.01981 | 0.03619 | 68 | 0.54740 | 0.58590 | Akkermansia.muciniphila | Visit.Name.NormD14 | 0.87253 |
| 660 | -0.00750 | 0.01372 | 68 | -0.54706 | 0.58613 | Acidaminococcus.intestini.CAG.325 | Visit.Name.NormD14 | 0.87253 |
| 661 | -0.00855 | 0.01595 | 68 | -0.53632 | 0.59349 | Sutterella.wadsworthensis | Visit.Name.NormD14 | 0.87356 |
| 662 | 0.00483 | 0.01216 | 68 | 0.39707 | 0.69256 | Methanobrevibacter.smithii | Visit.Name.NormD30 | 0.87473 |
| 663 | -0.00714 | 0.01811 | 68 | -0.39451 | 0.69444 | Blautia.sp..MP588D7 | Visit.Name.NormD30 | 0.87473 |
| 664 | -0.00733 | 0.01789 | 68 | -0.40975 | 0.68327 | Veillonella.atypica | Visit.Name.NormD30 | 0.87473 |
| 665 | -0.00483 | 0.01226 | 68 | -0.39368 | 0.69505 | Absiella.sp..MP355B5 | Visit.Name.NormD1 | 0.87549 |
| 666 | 0.01172 | 0.02253 | 68 | 0.52004 | 0.60473 | Bacteroides.sp..MP163B7 | Visit.Name.NormD14 | 0.87839 |
| 667 | -0.00669 | 0.01307 | 68 | -0.51219 | 0.61018 | Desulfovibrio.desulfuricans | Visit.Name.NormD14 | 0.87839 |
| 668 | -0.01365 | 0.03595 | 68 | -0.37981 | 0.70527 | Parabacteroides.distasonis | Visit.Name.NormD30 | 0.87859 |
| 669 | 0.01377 | 0.03792 | 68 | 0.36314 | 0.71763 | Akkermansia.muciniphila | Visit.Name.NormD30 | 0.87859 |
| 670 | 0.00485 | 0.01315 | 68 | 0.36890 | 0.71334 | Ruminococcus.bicirculans | Visit.Name.NormD30 | 0.87859 |
| 671 | 0.00350 | 0.01011 | 68 | 0.34676 | 0.72984 | Succinatimonas.hippei | Visit.Name.NormD30 | 0.88527 |
| 672 | -0.00900 | 0.02477 | 68 | -0.36317 | 0.71760 | Ruthenibacterium.sp..VE800.03 | Visit.Name.NormD1 | 0.88603 |
| 673 | -0.00688 | 0.01938 | 68 | -0.35498 | 0.72370 | Parabacteroides.sp..VE800.05 | Visit.Name.NormD1 | 0.88603 |

| | | | | | | | | |
|---|---|---|---|---|---|---|---|---|
| 674 | -0.00606 | 0.01682 | 68 | -0.36012 | 0.71988 | Erysipelotrichaceae.bacterium.6_1_45 | Visit.Name.NormD1 | 0.88603 |
| 675 | -0.00392 | 0.01057 | 68 | -0.37077 | 0.71196 | Absiella.sp..MP634G1_ig1 | Visit.Name.NormD3 | 0.88826 |
| 676 | 0.00225 | 0.00625 | 68 | 0.35932 | 0.72047 | Sutterella.sp..54_7 | Visit.Name.NormD3 | 0.89039 |
| 677 | 0.00336 | 0.01056 | 68 | 0.31824 | 0.75128 | Absiella.sp..MP634G1_ig1 | Visit.Name.NormD30 | 0.89471 |
| 678 | -0.00536 | 0.01672 | 68 | -0.32047 | 0.74959 | Sutterella.wadsworthensis | Visit.Name.NormD30 | 0.89471 |
| 679 | -0.02165 | 0.04559 | 68 | -0.47481 | 0.63644 | Lactobacillus.rhamnosus | Visit.Name.NormD14 | 0.89844 |
| 680 | 0.00489 | 0.01035 | 68 | 0.47287 | 0.63782 | Oscillospiraceae.bacterium.VE202.24 | Visit.Name.NormD14 | 0.89844 |
| 681 | -0.00336 | 0.01114 | 68 | -0.30133 | 0.76408 | Veillonella.sp..MP19D1_02.09 | Visit.Name.NormD30 | 0.90176 |
| 682 | -0.00775 | 0.01693 | 68 | -0.45779 | 0.64856 | Veillonella.parvula | Visit.Name.NormD14 | 0.90384 |
| 683 | -0.05600 | 0.05983 | 68 | -0.93599 | 0.35259 | Lactobacillus.rhamnosus | Visit.Name.NormD58 | 0.90568 |
| 684 | 0.01856 | 0.02016 | 68 | 0.92095 | 0.36033 | X.Ruminococcus..torques.CAG.61 | Visit.Name.NormD58 | 0.90776 |
| 685 | -0.00683 | 0.01810 | 68 | -0.37748 | 0.70699 | Blautia.sp..MP588D7 | Visit.Name.NormD21 | 0.90800 |
| 686 | 0.00492 | 0.01444 | 68 | 0.34096 | 0.73419 | X.Clostridium..symbiosum | Visit.Name.NormD21 | 0.91037 |
| 687 | -0.00627 | 0.01753 | 68 | -0.35777 | 0.72162 | Bacteroides.xylanisolvens | Visit.Name.NormD21 | 0.91037 |
| 688 | 0.00551 | 0.01631 | 68 | 0.33769 | 0.73663 | Blautia.sp..HK34 | Visit.Name.NormD21 | 0.91037 |
| 689 | 0.00240 | 0.00688 | 68 | 0.34945 | 0.72783 | Megasphaera.sp..UBA1696 | Visit.Name.NormD21 | 0.91037 |
| 690 | 0.00384 | 0.01176 | 68 | 0.32695 | 0.74471 | Parasutterella.excrementihominis | Visit.Name.NormD21 | 0.91174 |
| 691 | -0.00438 | 0.01354 | 68 | -0.32331 | 0.74745 | Bacteroides.sp..MP369D12 | Visit.Name.NormD3 | 0.91511 |
| 692 | -0.00465 | 0.01069 | 68 | -0.43521 | 0.66479 | Veillonella.sp..MP19D1_02.09 | Visit.Name.NormD14 | 0.91671 |
| 693 | 0.01484 | 0.05791 | 68 | 0.25623 | 0.79855 | Escherichia.coli | Visit.Name.NormD7 | 0.91763 |
| 694 | 0.00375 | 0.01623 | 68 | 0.23109 | 0.81794 | Bifidobacterium.dentium | Visit.Name.NormD1 | 0.91929 |
| 695 | -0.00880 | 0.04168 | 68 | -0.21126 | 0.83332 | Fusobacterium.sp..VE800.01 | Visit.Name.NormD1 | 0.91929 |
| 696 | -0.01548 | 0.07016 | 68 | -0.22062 | 0.82605 | Bacteroides.sp..VE800.11 | Visit.Name.NormD1 | 0.91929 |
| 697 | -0.00505 | 0.02396 | 68 | -0.21093 | 0.83358 | Lactobacillus.fermentum | Visit.Name.NormD1 | 0.91929 |
| 698 | -0.00438 | 0.02081 | 68 | -0.21028 | 0.83408 | Lachnospiraceae.bacterium.7_1_58FAA | Visit.Name.NormD1 | 0.91929 |
| 699 | 0.00263 | 0.01448 | 68 | 0.18199 | 0.85613 | Blautia.coccoides | Visit.Name.NormD1 | 0.91929 |
| 700 | -0.00225 | 0.01227 | 68 | -0.18338 | 0.85505 | Blautia.sp..VE303.05 | Visit.Name.NormD1 | 0.91929 |
| 701 | 0.00242 | 0.01243 | 68 | 0.19470 | 0.84621 | Dorea.sp..MP50E3 | Visit.Name.NormD1 | 0.91929 |
| 702 | 0.00600 | 0.02973 | 68 | 0.20177 | 0.84070 | Alistipes.sp..AF14.19 | Visit.Name.NormD1 | 0.91929 |
| 703 | -0.00204 | 0.00863 | 68 | -0.23611 | 0.81405 | Sutterella.sp..AM11.39 | Visit.Name.NormD1 | 0.91929 |
| 704 | 0.00216 | 0.01168 | 68 | 0.18463 | 0.85407 | Clostridiales.bacterium | Visit.Name.NormD1 | 0.91929 |
| 705 | 0.00890 | 0.02938 | 68 | 0.30292 | 0.76288 | Bacteroides.sp..MP487B8 | Visit.Name.NormD1 | 0.91929 |
| 706 | -0.00407 | 0.01862 | 68 | -0.21843 | 0.82775 | Bacteroides.sp..MP227D2 | Visit.Name.NormD1 | 0.91929 |
| 707 | 0.00325 | 0.01647 | 68 | 0.19739 | 0.84411 | Bacteroides.sp..MP244D10 | Visit.Name.NormD1 | 0.91929 |
| 708 | 0.00371 | 0.01396 | 68 | 0.26587 | 0.79115 | Prevotella.copri | Visit.Name.NormD1 | 0.91929 |
| 709 | -0.00339 | 0.01276 | 68 | -0.26597 | 0.79107 | Prevotella.copri | Visit.Name.NormD30 | 0.92527 |
| 710 | -0.00295 | 0.00973 | 68 | -0.30284 | 0.76293 | Neglecta.timonensis | Visit.Name.NormD3 | 0.92541 |
| 711 | -0.00311 | 0.01308 | 68 | -0.23799 | 0.81260 | Enterobacteriaceae.bacterium.9_2_54FAA | Visit.Name.NormD7 | 0.92566 |
| 712 | 0.00477 | 0.01173 | 68 | 0.40627 | 0.68582 | Blautia.sp..MP351A5 | Visit.Name.NormD14 | 0.92809 |
| 713 | 0.00338 | 0.00837 | 68 | 0.40436 | 0.68721 | Megasphaera.sp..MP182G6 | Visit.Name.NormD14 | 0.92809 |
| 714 | -0.01298 | 0.03398 | 68 | -0.38204 | 0.70362 | Bacteroides.thetaiotaomicron | Visit.Name.NormD14 | 0.93121 |
| 715 | -0.00597 | 0.01564 | 68 | -0.38188 | 0.70374 | Blautia.sp..HK34 | Visit.Name.NormD14 | 0.93121 |
| 716 | -0.00511 | 0.01375 | 68 | -0.37122 | 0.71163 | Pediococcus.acidilactici | Visit.Name.NormD14 | 0.93224 |
| 717 | -0.00482 | 0.03333 | 68 | -0.14446 | 0.88556 | Parabacteroides.sp..VE800.10 | Visit.Name.NormD1 | 0.93555 |
| 718 | -0.00174 | 0.01158 | 68 | -0.15035 | 0.88093 | Absiella.sp..MP634G1_ig1 | Visit.Name.NormD1 | 0.93555 |
| 719 | 0.00567 | 0.01933 | 68 | 0.29330 | 0.77019 | Cloacibacillus.evryensis | Visit.Name.NormD14 | 0.93567 |
| 720 | 0.00435 | 0.01489 | 68 | 0.29201 | 0.77117 | Bilophila.wadsworthia | Visit.Name.NormD14 | 0.93567 |
| 721 | 0.00338 | 0.01160 | 68 | 0.29171 | 0.77140 | Methanobrevibacter.smithii | Visit.Name.NormD14 | 0.93567 |
| 722 | -0.01213 | 0.03445 | 68 | -0.35206 | 0.72588 | Parabacteroides.distasonis | Visit.Name.NormD14 | 0.93567 |
| 723 | 0.00440 | 0.01264 | 68 | 0.34781 | 0.72906 | Collinsella.tanakaei | Visit.Name.NormD14 | 0.93567 |
| 724 | -0.00507 | 0.01714 | 68 | -0.29597 | 0.76816 | Veillonella.atypica | Visit.Name.NormD14 | 0.93567 |
| 725 | 0.00550 | 0.01799 | 68 | 0.30557 | 0.76087 | Alistipes.sp..HK66 | Visit.Name.NormD14 | 0.93567 |
| 726 | -0.01759 | 0.05792 | 68 | -0.30364 | 0.76233 | Klebsiella.michiganensis | Visit.Name.NormD14 | 0.93567 |
| 727 | -0.02022 | 0.02387 | 68 | -0.84700 | 0.39997 | Lachnospiraceae.bacterium.7_1_58FAA | Visit.Name.NormD58 | 0.93948 |
| 728 | -0.00864 | 0.00995 | 68 | -0.86829 | 0.38829 | Sutterella.sp..AM11.39 | Visit.Name.NormD58 | 0.93948 |
| 729 | 0.00856 | 0.01014 | 68 | 0.84404 | 0.40161 | Megasphaera.massiliensis | Visit.Name.NormD58 | 0.93948 |
| 730 | -0.01536 | 0.01807 | 68 | -0.84983 | 0.39840 | Enterococcus.faecalis | Visit.Name.NormD58 | 0.93948 |
| 731 | 0.00918 | 0.03816 | 68 | 0.24060 | 0.81059 | Parabacteroides.sp..VE800.07 | Visit.Name.NormD3 | 0.93971 |
| 732 | -0.00383 | 0.01384 | 68 | -0.27703 | 0.78260 | Phascolarctobacterium.sp..MUR_MP6E9_ig1 | Visit.Name.NormD3 | 0.93971 |
| 733 | -0.00291 | 0.01124 | 68 | -0.25911 | 0.79634 | Absiella.sp..MP355B5 | Visit.Name.NormD3 | 0.93971 |
| 734 | 0.00342 | 0.01276 | 68 | 0.26784 | 0.78963 | Prevotella.copri | Visit.Name.NormD3 | 0.93971 |
| 735 | 0.00300 | 0.01206 | 68 | 0.24876 | 0.80430 | Gemella.bergeri | Visit.Name.NormD3 | 0.93971 |
| 736 | -0.00690 | 0.02413 | 68 | -0.28578 | 0.77591 | Citrobacter.freundii | Visit.Name.NormD21 | 0.94115 |
| 737 | -0.00878 | 0.03194 | 68 | -0.27488 | 0.78424 | Bacteroides.ovatus | Visit.Name.NormD14 | 0.94253 |
| 738 | -0.00254 | 0.01358 | 68 | -0.18671 | 0.85244 | Blautia.coccoides | Visit.Name.NormD7 | 0.95444 |
| 739 | -0.00262 | 0.01344 | 68 | -0.19480 | 0.84613 | Flavobacteriales.bacterium | Visit.Name.NormD7 | 0.95444 |
| 740 | 0.00327 | 0.01528 | 68 | 0.21382 | 0.83133 | Acidaminococcus.intestini | Visit.Name.NormD3 | 0.95530 |
| 741 | -0.00225 | 0.01249 | 68 | -0.18008 | 0.85762 | Roseburia.hominis | Visit.Name.NormD14 | 0.96025 |
| 742 | 0.00278 | 0.01560 | 68 | 0.17843 | 0.85892 | X.Ruminococcus..torques.CAG.61 | Visit.Name.NormD14 | 0.96025 |
| 743 | 0.00150 | 0.01053 | 68 | 0.14270 | 0.88695 | X.Clostridium..leptum | Visit.Name.NormD14 | 0.96025 |
| 744 | 0.00271 | 0.01608 | 68 | 0.16874 | 0.86651 | Clostridioides.difficile | Visit.Name.NormD14 | 0.96025 |
| 745 | 0.00196 | 0.00979 | 68 | 0.20004 | 0.84205 | Streptococcus.thermophilus | Visit.Name.NormD14 | 0.96025 |
| 746 | -0.00224 | 0.01091 | 68 | -0.20540 | 0.83787 | Dorea.sp..MP50E3 | Visit.Name.NormD14 | 0.96025 |
| 747 | 0.00200 | 0.01105 | 68 | 0.18139 | 0.85660 | X.Clostridium..citroniae | Visit.Name.NormD14 | 0.96025 |
| 748 | -0.00412 | 0.01825 | 68 | -0.22564 | 0.82215 | Bacteroides.sp..MP264A7 | Visit.Name.NormD14 | 0.96025 |

| | | | | | | | | |
|---|---|---|---|---|---|---|---|---|
| 749 | 0.00733 | 0.03635 | 68 | 0.20178 | 0.84069 | Bacteroides.fragilis | Visit.Name.NormD14 | 0.96025 |
| 750 | 0.00332 | 0.02200 | 68 | 0.15095 | 0.88046 | Lactobacillus.salivarius | Visit.Name.NormD14 | 0.96025 |
| 751 | -0.00205 | 0.01254 | 68 | -0.16375 | 0.87042 | Flavobacteriales.bacterium | Visit.Name.NormD14 | 0.96025 |
| 752 | -0.00364 | 0.02317 | 68 | -0.15720 | 0.87556 | Citrobacter.freundii | Visit.Name.NormD14 | 0.96025 |
| 753 | -0.00193 | 0.01388 | 68 | -0.13908 | 0.88980 | Bacteroides.sp..MP369D12 | Visit.Name.NormD7 | 0.96116 |
| 754 | 0.00164 | 0.01359 | 68 | 0.12098 | 0.90406 | Collinsella.tanakaei | Visit.Name.NormD7 | 0.96116 |
| 755 | -0.00345 | 0.02754 | 68 | -0.12520 | 0.90074 | Bacteroides.sp..MP487B8 | Visit.Name.NormD7 | 0.96116 |
| 756 | 0.00141 | 0.01349 | 68 | 0.10443 | 0.91714 | Ruminococcus.bicirculans | Visit.Name.NormD7 | 0.96116 |
| 757 | -0.00233 | 0.01747 | 68 | -0.13339 | 0.89428 | Bacteroides.sp..MP227D2 | Visit.Name.NormD7 | 0.96116 |
| 758 | -0.00163 | 0.01556 | 68 | -0.10491 | 0.91675 | Bacteroides.sp..MP244D10 | Visit.Name.NormD7 | 0.96116 |
| 759 | 0.00158 | 0.01470 | 68 | 0.10732 | 0.91485 | Acidaminococcus.intestini.CAG.325 | Visit.Name.NormD7 | 0.96116 |
| 760 | -0.00541 | 0.03985 | 68 | -0.13584 | 0.89235 | Pyramidobacter.piscolens | Visit.Name.NormD7 | 0.96116 |
| 761 | 0.00291 | 0.02256 | 68 | 0.12917 | 0.89760 | X.Clostridium..clostridioforme | Visit.Name.NormD14 | 0.96182 |
| 762 | -0.00104 | 0.01000 | 68 | -0.10363 | 0.91777 | Eubacterium.sp..CAG.38 | Visit.Name.NormD14 | 0.96182 |
| 763 | 0.00198 | 0.01850 | 68 | 0.10721 | 0.91494 | Acidaminococcus.sp..MP76F2 | Visit.Name.NormD14 | 0.96182 |
| 764 | -0.00163 | 0.01463 | 68 | -0.11159 | 0.91148 | Bacteroides.sp..MP244D10 | Visit.Name.NormD14 | 0.96182 |
| 765 | -0.00418 | 0.01939 | 68 | -0.21545 | 0.83006 | Acidaminococcus.sp..MP76F2 | Visit.Name.NormD30 | 0.96228 |
| 766 | 0.00214 | 0.02024 | 68 | 0.10594 | 0.91595 | Cloacibacillus.evryensis | Visit.Name.NormD21 | 0.96317 |
| 767 | -0.00212 | 0.01174 | 68 | -0.18083 | 0.85703 | Alistipes.putredinis | Visit.Name.NormD21 | 0.96317 |
| 768 | -0.00617 | 0.02885 | 68 | -0.21390 | 0.83126 | Alistipes.sp..MP104G4_1 | Visit.Name.NormD21 | 0.96317 |
| 769 | -0.00156 | 0.01022 | 68 | -0.15282 | 0.87900 | Streptococcus.thermophilus | Visit.Name.NormD21 | 0.96317 |
| 770 | -0.00874 | 0.05643 | 68 | -0.15497 | 0.87731 | Escherichia.coli | Visit.Name.NormD21 | 0.96317 |
| 771 | 0.00190 | 0.01344 | 68 | 0.14138 | 0.88799 | Klebsiella.oxytoca | Visit.Name.NormD21 | 0.96317 |
| 772 | -0.00825 | 0.03554 | 68 | -0.23199 | 0.81724 | Bacteroides.thetaiotaomicron | Visit.Name.NormD21 | 0.96317 |
| 773 | -0.00238 | 0.01332 | 68 | -0.17836 | 0.85897 | Bacteroides.ovatus.CAG.22 | Visit.Name.NormD21 | 0.96317 |
| 774 | -0.00206 | 0.01309 | 68 | -0.15718 | 0.87557 | Barnesiella.intestinihominis | Visit.Name.NormD21 | 0.96317 |
| 775 | 0.00270 | 0.01404 | 68 | 0.19237 | 0.84803 | X.Ruminococcus..sp..MP50A2 | Visit.Name.NormD21 | 0.96317 |
| 776 | -0.00650 | 0.03594 | 68 | -0.18083 | 0.85704 | Parabacteroides.distasonis | Visit.Name.NormD21 | 0.96317 |
| 777 | 0.00185 | 0.01125 | 68 | 0.16472 | 0.86965 | Roseburia.faecis | Visit.Name.NormD21 | 0.96317 |
| 778 | 0.00235 | 0.02299 | 68 | 0.10242 | 0.91873 | Lactobacillus.salivarius | Visit.Name.NormD21 | 0.96317 |
| 779 | -0.00142 | 0.01368 | 68 | -0.10391 | 0.91754 | Desulfovibrio.desulfuricans | Visit.Name.NormD21 | 0.96317 |
| 780 | -0.00730 | 0.06050 | 68 | -0.12066 | 0.90432 | Klebsiella.michiganensis | Visit.Name.NormD21 | 0.96317 |
| 781 | -0.00203 | 0.01274 | 68 | -0.15894 | 0.87418 | Prevotella.copri | Visit.Name.NormD21 | 0.96317 |
| 782 | 0.00152 | 0.01492 | 68 | 0.10201 | 0.91905 | Atopobium.minutum | Visit.Name.NormD21 | 0.96317 |
| 783 | 0.00115 | 0.01237 | 68 | 0.09264 | 0.92646 | Gemella.bergeri | Visit.Name.NormD7 | 0.96322 |
| 784 | 0.00146 | 0.01678 | 68 | 0.08684 | 0.93105 | Clostridioides.difficile | Visit.Name.NormD21 | 0.96396 |
| 785 | 0.00125 | 0.01520 | 68 | 0.08246 | 0.93452 | Bacteroides.sp..MP244D10 | Visit.Name.NormD21 | 0.96396 |
| 786 | -0.00140 | 0.01432 | 68 | -0.09746 | 0.92265 | Flavobacteriales.bacterium | Visit.Name.NormD1 | 0.96693 |
| 787 | 0.00078 | 0.01306 | 68 | 0.05974 | 0.95254 | Roseburia.hominis | Visit.Name.NormD21 | 0.96769 |
| 788 | -0.00112 | 0.02478 | 68 | -0.04519 | 0.96409 | Alistipes.finegoldii | Visit.Name.NormD21 | 0.96769 |
| 789 | -0.00108 | 0.02657 | 68 | -0.04066 | 0.96769 | Raoultella.ornithinolytica | Visit.Name.NormD21 | 0.96769 |
| 790 | 0.00047 | 0.00801 | 68 | 0.05866 | 0.95339 | Megasphaera.massiliensis | Visit.Name.NormD21 | 0.96769 |
| 791 | -0.00332 | 0.01876 | 68 | -0.17693 | 0.86009 | Alistipes.sp..VE800.08 | Visit.Name.NormD3 | 0.97130 |
| 792 | -0.00424 | 0.02365 | 68 | -0.17910 | 0.85839 | Bacteroides.sp..MP163B7 | Visit.Name.NormD3 | 0.97130 |
| 793 | 0.00124 | 0.01988 | 68 | 0.06236 | 0.95046 | Phascolarctobacterium.sp..VE800.04 | Visit.Name.NormD1 | 0.97274 |
| 794 | -0.00284 | 0.04175 | 68 | -0.06804 | 0.94595 | Parabacteroides.sp..VE800.07 | Visit.Name.NormD1 | 0.97274 |
| 795 | -0.00180 | 0.02587 | 68 | -0.06955 | 0.94475 | X.Clostridium..clostridioforme | Visit.Name.NormD1 | 0.97274 |
| 796 | -0.00075 | 0.01048 | 68 | -0.07155 | 0.94317 | Streptococcus.thermophilus | Visit.Name.NormD7 | 0.97287 |
| 797 | 0.00359 | 0.02201 | 68 | 0.16297 | 0.87103 | Lactobacillus.fermentum | Visit.Name.NormD3 | 0.97525 |
| 798 | 0.00071 | 0.02222 | 68 | 0.03206 | 0.97452 | Cloacibacillus.evryensis | Visit.Name.NormD1 | 0.97563 |
| 799 | 0.00067 | 0.01479 | 68 | 0.04503 | 0.96421 | Bacteroides.sp..MP369D12 | Visit.Name.NormD1 | 0.97563 |
| 800 | -0.00063 | 0.02049 | 68 | -0.03066 | 0.97563 | Alistipes.sp..VE800.08 | Visit.Name.NormD1 | 0.97563 |
| 801 | -0.00195 | 0.01310 | 68 | -0.14859 | 0.88232 | Barnesiella.intestinihominis | Visit.Name.NormD3 | 0.97952 |
| 802 | 0.00204 | 0.01123 | 68 | 0.18132 | 0.85665 | Absiella.sp..MP355B5 | Visit.Name.NormD30 | 0.98299 |
| 803 | -0.01049 | 0.06054 | 68 | -0.17330 | 0.86293 | Klebsiella.michiganensis | Visit.Name.NormD30 | 0.98299 |
| 804 | -0.01390 | 0.01863 | 68 | -0.74606 | 0.45820 | Blautia.sp..HF17 | Visit.Name.NormD58 | 0.98400 |
| 805 | 0.01254 | 0.01671 | 68 | 0.75043 | 0.45558 | Collinsella.tanakaei | Visit.Name.NormD58 | 0.98400 |
| 806 | 0.01123 | 0.01494 | 68 | 0.75159 | 0.45489 | Ruminococcus.sp..MP226F3 | Visit.Name.NormD58 | 0.98400 |
| 807 | 0.01076 | 0.01438 | 68 | 0.74791 | 0.45709 | Pseudomonas.aeruginosa | Visit.Name.NormD58 | 0.98400 |
| 808 | 0.01227 | 0.01606 | 68 | 0.76383 | 0.44761 | Tyzzerella.sp..MP10F1re | Visit.Name.NormD58 | 0.98400 |
| 809 | -0.01626 | 0.02218 | 68 | -0.73313 | 0.46600 | Veillonella.parvula | Visit.Name.NormD58 | 0.98462 |
| 810 | 0.00638 | 0.01851 | 68 | 0.34445 | 0.73157 | Bifidobacterium.dentium | Visit.Name.NormD58 | 0.98501 |
| 811 | -0.00604 | 0.02849 | 68 | -0.21199 | 0.83275 | Ruthenibacterium.sp..VE800.03 | Visit.Name.NormD58 | 0.98501 |
| 812 | -0.00139 | 0.02563 | 68 | -0.05424 | 0.95691 | Cloacibacillus.evryensis | Visit.Name.NormD58 | 0.98501 |
| 813 | 0.00405 | 0.01969 | 68 | 0.20553 | 0.83777 | Bilophila.wadsworthia | Visit.Name.NormD58 | 0.98501 |
| 814 | -0.00405 | 0.01226 | 68 | -0.33007 | 0.74236 | Neglecta.timonensis | Visit.Name.NormD58 | 0.98501 |
| 815 | 0.00828 | 0.01648 | 68 | 0.50223 | 0.61713 | Roseburia.hominis | Visit.Name.NormD58 | 0.98501 |
| 816 | -0.00916 | 0.01536 | 68 | -0.59650 | 0.55282 | Methanobrevibacter.smithii | Visit.Name.NormD58 | 0.98501 |
| 817 | 0.00277 | 0.01699 | 68 | 0.16273 | 0.87121 | Bacteroides.sp..MP369D12 | Visit.Name.NormD58 | 0.98501 |
| 818 | -0.00687 | 0.01666 | 68 | -0.41263 | 0.68118 | Blautia.coccoides | Visit.Name.NormD58 | 0.98501 |
| 819 | 0.00063 | 0.01385 | 68 | 0.04584 | 0.96357 | X.Clostridium..leptum | Visit.Name.NormD58 | 0.98501 |
| 820 | -0.00659 | 0.01547 | 68 | -0.42617 | 0.67133 | Eggerthella.lenta | Visit.Name.NormD58 | 0.98501 |
| 821 | 0.00633 | 0.02980 | 68 | 0.21233 | 0.83249 | X.Clostridium..clostridioforme | Visit.Name.NormD58 | 0.98501 |
| 822 | 0.00401 | 0.02098 | 68 | 0.19098 | 0.84911 | Clostridioides.difficile | Visit.Name.NormD58 | 0.98501 |
| 823 | 0.00415 | 0.01486 | 68 | 0.27935 | 0.78082 | Parasutterella.excrementihominis | Visit.Name.NormD58 | 0.98501 |

| | | | | | | | | |
|---|---|---|---|---|---|---|---|---|
| 824 | -0.00123 | 0.01420 | 68 | -0.08668 | 0.93118 | Lachnospiraceae.bacterium.5_1_57FAA | Visit.Name.NormD58 | 0.98501 |
| 825 | 0.00898 | 0.01281 | 68 | 0.70111 | 0.48562 | Streptococcus.thermophilus | Visit.Name.NormD58 | 0.98501 |
| 826 | 0.00866 | 0.01408 | 68 | 0.61508 | 0.54055 | Blautia.sp..VE303.05 | Visit.Name.NormD58 | 0.98501 |
| 827 | -0.00275 | 0.03141 | 68 | -0.08770 | 0.93037 | Alistipes.finegoldii | Visit.Name.NormD58 | 0.98501 |
| 828 | -0.00194 | 0.01810 | 68 | -0.10695 | 0.91514 | X.Clostridium..symbiosum | Visit.Name.NormD58 | 0.98501 |
| 829 | 0.00389 | 0.01926 | 68 | 0.20177 | 0.84070 | Erysipelotrichaceae.bacterium.6_1_45 | Visit.Name.NormD58 | 0.98501 |
| 830 | -0.00788 | 0.01362 | 68 | -0.57879 | 0.56464 | Oscillospiraceae.bacterium.VE202.24 | Visit.Name.NormD58 | 0.98501 |
| 831 | -0.00460 | 0.02207 | 68 | -0.20850 | 0.83546 | Bacteroides.xylanisolvens | Visit.Name.NormD58 | 0.98501 |
| 832 | 0.00557 | 0.01521 | 68 | 0.36620 | 0.71535 | Blautia.sp..MP351A5 | Visit.Name.NormD58 | 0.98501 |
| 833 | -0.00966 | 0.02709 | 68 | -0.35663 | 0.72248 | Bacteroides.vulgatus | Visit.Name.NormD58 | 0.98501 |
| 834 | -0.00682 | 0.01483 | 68 | -0.45982 | 0.64711 | Streptococcus.sp..MP111E2 | Visit.Name.NormD58 | 0.98501 |
| 835 | -0.00659 | 0.03416 | 68 | -0.19285 | 0.84765 | Alistipes.sp..AF14.19 | Visit.Name.NormD58 | 0.98501 |
| 836 | 0.00778 | 0.01653 | 68 | 0.47082 | 0.63928 | Bacteroides.ovatus.CAG.22 | Visit.Name.NormD58 | 0.98501 |
| 837 | -0.00993 | 0.02760 | 68 | -0.35984 | 0.72008 | Faecalibacterium.prausnitzii | Visit.Name.NormD58 | 0.98501 |
| 838 | 0.02655 | 0.07610 | 68 | 0.34891 | 0.72823 | Klebsiella.pneumoniae | Visit.Name.NormD58 | 0.98501 |
| 839 | -0.00894 | 0.01637 | 68 | -0.54641 | 0.58657 | Barnesiella.intestinihominis | Visit.Name.NormD58 | 0.98501 |
| 840 | 0.01531 | 0.02355 | 68 | 0.65029 | 0.51769 | Alistipes.sp..VE800.08 | Visit.Name.NormD58 | 0.98501 |
| 841 | -0.00929 | 0.01860 | 68 | -0.49933 | 0.61916 | Roseburia.intestinalis | Visit.Name.NormD58 | 0.98501 |
| 842 | -0.00075 | 0.01407 | 68 | -0.05361 | 0.95740 | Blautia.sp..MP78G6 | Visit.Name.NormD58 | 0.98501 |
| 843 | -0.00657 | 0.03318 | 68 | -0.19790 | 0.84371 | Raoultella.ornithinolytica | Visit.Name.NormD58 | 0.98501 |
| 844 | 0.00249 | 0.01769 | 68 | 0.14091 | 0.88835 | X.Ruminococcus..sp..MP50A2 | Visit.Name.NormD58 | 0.98501 |
| 845 | -0.00457 | 0.01779 | 68 | -0.25667 | 0.79821 | Bacteroides.dorei | Visit.Name.NormD58 | 0.98501 |
| 846 | -0.00636 | 0.01386 | 68 | -0.45847 | 0.64807 | Veillonella.sp..MP19D1_02.09 | Visit.Name.NormD58 | 0.98501 |
| 847 | 0.02142 | 0.03025 | 68 | 0.70787 | 0.48144 | X.Ruminococcus..sp..MP495E3 | Visit.Name.NormD58 | 0.98501 |
| 848 | -0.00116 | 0.01369 | 68 | -0.08453 | 0.93288 | Desulfovibrio.fairfieldensis | Visit.Name.NormD58 | 0.98501 |
| 849 | -0.01500 | 0.02238 | 68 | -0.67007 | 0.50508 | Veillonella.atypica | Visit.Name.NormD58 | 0.98501 |
| 850 | -0.00139 | 0.01436 | 68 | -0.09702 | 0.92300 | X.Clostridium..citroniae | Visit.Name.NormD58 | 0.98501 |
| 851 | -0.00898 | 0.02781 | 68 | -0.32280 | 0.74784 | Eubacterium.sp..CAG.180 | Visit.Name.NormD58 | 0.98501 |
| 852 | 0.00168 | 0.01346 | 68 | 0.12480 | 0.90105 | Clostridiales.bacterium | Visit.Name.NormD58 | 0.98501 |
| 853 | -0.00133 | 0.01408 | 68 | -0.09448 | 0.92501 | Absiella.sp..MP355B5 | Visit.Name.NormD58 | 0.98501 |
| 854 | 0.00471 | 0.01110 | 68 | 0.42464 | 0.67244 | Megasphaera.sp..MP182G6 | Visit.Name.NormD58 | 0.98501 |
| 855 | 0.00043 | 0.00872 | 68 | 0.04884 | 0.96119 | Megasphaera.sp..UBA1696 | Visit.Name.NormD58 | 0.98501 |
| 856 | -0.01255 | 0.03992 | 68 | -0.31438 | 0.75420 | Lachnospiraceae.bacterium.1_1_57FAA | Visit.Name.NormD58 | 0.98501 |
| 857 | 0.01863 | 0.04794 | 68 | 0.38866 | 0.69874 | Akkermansia.muciniphila | Visit.Name.NormD58 | 0.98501 |
| 858 | -0.01125 | 0.01983 | 68 | -0.56723 | 0.57243 | Eisenbergiella.sp..OF01.20 | Visit.Name.NormD58 | 0.98501 |
| 859 | 0.01285 | 0.02991 | 68 | 0.42955 | 0.66888 | Bacteroides.sp..MP163B7 | Visit.Name.NormD58 | 0.98501 |
| 860 | 0.00496 | 0.02416 | 68 | 0.20527 | 0.83797 | Bacteroides.sp..MP264A7 | Visit.Name.NormD58 | 0.98501 |
| 861 | 0.00290 | 0.01657 | 68 | 0.17475 | 0.86179 | Ruminococcus.bicirculans | Visit.Name.NormD58 | 0.98501 |
| 862 | 0.00715 | 0.02892 | 68 | 0.24730 | 0.80542 | Lactobacillus.salivarius | Visit.Name.NormD58 | 0.98501 |
| 863 | -0.00661 | 0.01813 | 68 | -0.36464 | 0.71651 | Parvimonas.micra | Visit.Name.NormD58 | 0.98501 |
| 864 | -0.01083 | 0.01800 | 68 | -0.60195 | 0.54921 | Pediococcus.acidilactici | Visit.Name.NormD58 | 0.98501 |
| 865 | -0.00446 | 0.01277 | 68 | -0.34937 | 0.72789 | Succinatimonas.hippei | Visit.Name.NormD58 | 0.98501 |
| 866 | 0.00670 | 0.01647 | 68 | 0.40674 | 0.68547 | Flavobacteriales.bacterium | Visit.Name.NormD58 | 0.98501 |
| 867 | -0.01395 | 0.02386 | 68 | -0.58477 | 0.56064 | Alistipes.sp..HK66 | Visit.Name.NormD58 | 0.98501 |
| 868 | 0.00071 | 0.01875 | 68 | 0.03778 | 0.96998 | Bacteroides.sp..MP244D10 | Visit.Name.NormD58 | 0.98501 |
| 869 | -0.02361 | 0.07605 | 68 | -0.31042 | 0.75719 | Klebsiella.michiganensis | Visit.Name.NormD58 | 0.98501 |
| 870 | -0.00971 | 0.03001 | 68 | -0.32369 | 0.74716 | Citrobacter.freundii | Visit.Name.NormD58 | 0.98501 |
| 871 | -0.01390 | 0.02012 | 68 | -0.69078 | 0.49206 | Shigella.dysenteriae | Visit.Name.NormD58 | 0.98501 |
| 872 | 0.00270 | 0.01799 | 68 | 0.15018 | 0.88106 | Acidaminococcus.intestini.CAG.325 | Visit.Name.NormD58 | 0.98501 |
| 873 | -0.02428 | 0.04883 | 68 | -0.49723 | 0.62063 | Pyramidobacter.piscolens | Visit.Name.NormD58 | 0.98501 |
| 874 | 0.00308 | 0.01608 | 68 | 0.19175 | 0.84851 | Prevotella.copri | Visit.Name.NormD58 | 0.98501 |
| 875 | 0.00134 | 0.01888 | 68 | 0.07100 | 0.94360 | Atopobium.minutum | Visit.Name.NormD58 | 0.98501 |
| 876 | 0.00840 | 0.01523 | 68 | 0.55166 | 0.58299 | Gemella.bergeri | Visit.Name.NormD58 | 0.98501 |
| 877 | -0.00082 | 0.01359 | 68 | -0.06068 | 0.95179 | Eubacterium.sp..VE800.02 | Visit.Name.NormD3 | 0.98611 |
| 878 | -0.00046 | 0.01625 | 68 | -0.02815 | 0.97763 | X.Ruminococcus..torques.CAG.61 | Visit.Name.NormD3 | 0.98611 |
| 879 | 0.00158 | 0.01909 | 68 | 0.08293 | 0.93415 | Lachnospiraceae.bacterium.7_1_58FAA | Visit.Name.NormD3 | 0.98611 |
| 880 | -0.00033 | 0.01772 | 68 | -0.01880 | 0.98505 | Parabacteroides.sp..VE800.05 | Visit.Name.NormD3 | 0.98611 |
| 881 | 0.00048 | 0.02724 | 68 | 0.01747 | 0.98611 | Alistipes.sp..AF14.19 | Visit.Name.NormD3 | 0.98611 |
| 882 | 0.00081 | 0.03596 | 68 | 0.02261 | 0.98203 | Parabacteroides.distasonis | Visit.Name.NormD3 | 0.98611 |
| 883 | -0.00569 | 0.04489 | 68 | -0.12684 | 0.89944 | Bacteroides.sp..VE800.09 | Visit.Name.NormD3 | 0.98611 |
| 884 | 0.00070 | 0.02685 | 68 | 0.02615 | 0.97921 | Bacteroides.sp..MP487B8 | Visit.Name.NormD3 | 0.98611 |
| 885 | 0.00050 | 0.01437 | 68 | 0.03510 | 0.97210 | Pediococcus.acidilactici | Visit.Name.NormD3 | 0.98611 |
| 886 | -0.00099 | 0.01704 | 68 | -0.05795 | 0.95396 | Bacteroides.sp..MP227D2 | Visit.Name.NormD3 | 0.98611 |
| 887 | 0.00102 | 0.01311 | 68 | 0.07759 | 0.93838 | Flavobacteriales.bacterium | Visit.Name.NormD3 | 0.98611 |
| 888 | -0.00095 | 0.01520 | 68 | -0.06247 | 0.95037 | Bacteroides.sp..MP244D10 | Visit.Name.NormD3 | 0.98611 |
| 889 | -0.00032 | 0.01288 | 68 | -0.02487 | 0.98023 | Intestinibacter.sp..MP113D11 | Visit.Name.NormD3 | 0.98611 |
| 890 | 0.00164 | 0.03651 | 68 | 0.04489 | 0.96433 | Bacteroides.thetaiotaomicron | Visit.Name.NormD7 | 0.98693 |
| 891 | 0.00040 | 0.01785 | 68 | 0.02218 | 0.98237 | Bacteroides.sp..MP327C3 | Visit.Name.NormD58 | 0.98993 |
| 892 | 0.00036 | 0.01152 | 68 | 0.03086 | 0.97547 | Absiella.sp..MP355B5 | Visit.Name.NormD7 | 0.99059 |
| 893 | -0.00156 | 0.01022 | 68 | -0.15286 | 0.87896 | Streptococcus.thermophilus | Visit.Name.NormD30 | 0.99114 |
| 894 | 0.00171 | 0.01178 | 68 | 0.14490 | 0.88522 | Streptococcus.sp..MP111E2 | Visit.Name.NormD30 | 0.99114 |
| 895 | 0.00016 | 0.01664 | 68 | 0.00974 | 0.99226 | X.Ruminococcus..torques.CAG.61 | Visit.Name.NormD7 | 0.99226 |
| 896 | -0.00011 | 0.01059 | 68 | -0.01017 | 0.99191 | Lactobacillus.paracasei | Visit.Name.NormD7 | 0.99226 |
| 897 | -0.00144 | 0.01175 | 68 | -0.12246 | 0.90289 | Alistipes.putredinis | Visit.Name.NormD30 | 0.99306 |
| 898 | 0.00126 | 0.01481 | 68 | 0.08512 | 0.93241 | Blautia.sp..HF17 | Visit.Name.NormD30 | 0.99306 |

| | | | | | | | | |
|---|---|---|---|---|---|---|---|---|
| 899 | 0.00108 | 0.01177 | 68 | 0.09145 | 0.92740 | Parasutterella.excrementihominis | Visit.Name.NormD30 | 0.99306 |
| 900 | 0.00133 | 0.01332 | 68 | 0.10006 | 0.92059 | Bacteroides.ovatus.CAG.22 | Visit.Name.NormD30 | 0.99306 |
| 901 | 0.00075 | 0.00802 | 68 | 0.09297 | 0.92620 | Megasphaera.massiliensis | Visit.Name.NormD30 | 0.99306 |
| 902 | -0.00138 | 0.01125 | 68 | -0.12266 | 0.90274 | Roseburia.faecis | Visit.Name.NormD30 | 0.99306 |
| 903 | -0.00130 | 0.02026 | 68 | -0.06391 | 0.94923 | Cloacibacillus.evryensis | Visit.Name.NormD30 | 0.99332 |
| 904 | -0.00181 | 0.03330 | 68 | -0.05447 | 0.95672 | Bacteroides.ovatus | Visit.Name.NormD30 | 0.99332 |
| 905 | 0.00218 | 0.03558 | 68 | 0.06131 | 0.95129 | Bacteroides.thetaiotaomicron | Visit.Name.NormD30 | 0.99332 |
| 906 | -0.00107 | 0.02300 | 68 | -0.04657 | 0.96299 | Lactobacillus.salivarius | Visit.Name.NormD30 | 0.99332 |
| 907 | -0.00022 | 0.01559 | 68 | -0.01407 | 0.98881 | Bilophila.wadsworthia | Visit.Name.NormD30 | 0.99642 |
| 908 | -0.00079 | 0.02657 | 68 | -0.02980 | 0.97631 | Raoultella.ornithinolytica | Visit.Name.NormD30 | 0.99642 |
| 909 | -0.00028 | 0.01493 | 68 | -0.01841 | 0.98536 | Atopobium.minutum | Visit.Name.NormD30 | 0.99642 |
| 910 | -0.00011 | 0.00929 | 68 | -0.01162 | 0.99076 | Neglecta.timonensis | Visit.Name.NormD14 | 0.99677 |
| 911 | 0.00052 | 0.01124 | 68 | 0.04626 | 0.96324 | Parasutterella.excrementihominis | Visit.Name.NormD14 | 0.99677 |
| 912 | 0.00060 | 0.05402 | 68 | 0.01103 | 0.99123 | Escherichia.coli | Visit.Name.NormD14 | 0.99677 |
| 913 | 0.00026 | 0.02548 | 68 | 0.01021 | 0.99189 | Raoultella.ornithinolytica | Visit.Name.NormD14 | 0.99677 |
| 914 | -0.00008 | 0.01344 | 68 | -0.00614 | 0.99512 | X.Ruminococcus..sp..MP50A2 | Visit.Name.NormD14 | 0.99677 |
| 915 | 0.00003 | 0.00657 | 68 | 0.00406 | 0.99677 | Megasphaera.sp..UBA1696 | Visit.Name.NormD14 | 0.99677 |
| 916 | 0.00005 | 0.01344 | 68 | 0.00380 | 0.99698 | Klebsiella.oxytoca | Visit.Name.NormD30 | 0.99698 |
| 917 | 0.00003 | 0.01677 | 68 | 0.00170 | 0.99865 | Klebsiella.oxytoca | Visit.Name.NormD58 | 0.99865 |

**Supplementary Table S14: Ordinal Analysis of Baseline Factors Impacting Colonization in Melanoma Cohort**

| | Predictors | coeff | dataset | Species | coeff_logic | Time |
|---|---|---|---|---|---|---|
| 1 | Phylum_Firmicutes | 1.3162639 | Mic_Phylum | VE800-01 | 1 | Baseline |
| 2 | Order_Clostridiales | 1.1094642 | Mic_Order | VE800-01 | 1 | Baseline |
| 3 | Order_uncl_Order_of_Proteobacteria | 7.0443603 | Mic_Order | VE800-01 | 1 | Baseline |
| 4 | Order_Burkholderiales | -12.0495527 | Mic_Order | VE800-01 | -1 | Baseline |
| 5 | Order_uncl_Order_of_Firmicutes | -9.2456784 | Mic_Order | VE800-01 | -1 | Baseline |
| 6 | CD8_TNF_Cytokines | -0.0142537 | Immune_Cells | VE800-01 | -1 | Baseline |
| 7 | X2.Methylbutyric.acid | -0.6844564 | SCFA | VE800-02 | -1 | Baseline |
| 8 | Taurodeoxycholic.Acid | 0.3892978 | BA | VE800-02 | 1 | Baseline |
| 9 | Oscillibacter.sp..CAG.241_62_21 | 15.3305776 | Mic_Species | VE800-02 | 1 | Baseline |
| 10 | Live.Cells_CD3._NonMAIT_CD4._TF | -0.0165729 | Immune_Cells | VE800-02 | -1 | Baseline |
| 11 | CD8_IFNg_Cytokines | -0.0027507 | Immune_Cells | VE800-02 | -1 | Baseline |
| 12 | NK_CD94_CD107a_Cytokines | -0.0314622 | Immune_Cells | VE800-02 | -1 | Baseline |
| 13 | Isobutyric.acid | -2.4214889 | SCFA | VE800-03 | -1 | Baseline |
| 14 | Phylum_Actinobacteria | 4.2234283 | Mic_Phylum | VE800-03 | 1 | Baseline |
| 15 | Class_Actinobacteria | 2.3337037 | Mic_Class | VE800-03 | 1 | Baseline |
| 16 | Order_Bifidobacteriales | 2.3337037 | Mic_Order | VE800-03 | 1 | Baseline |
| 17 | CD8_IFNg_Cytokines | -0.0053180 | Immune_Cells | VE800-03 | -1 | Baseline |
| 18 | Isovaleric.acid | -0.5792901 | SCFA | VE800-04 | -1 | Baseline |
| 19 | Deoxycholic.Acid | -0.4839097 | BA | VE800-04 | -1 | Baseline |
| 20 | Glycolithocholic.Acid | -0.1396161 | BA | VE800-04 | -1 | Baseline |
| 21 | Phylum_Bacteroidetes | 0.6036193 | Mic_Phylum | VE800-04 | 1 | Baseline |
| 22 | Class_Erysipelotrichia | 113.0093396 | Mic_Class | VE800-04 | 1 | Baseline |
| 23 | Order_Bacteroidales | 0.8459270 | Mic_Order | VE800-04 | 1 | Baseline |
| 24 | Order_Bifidobacteriales | 2.7044983 | Mic_Order | VE800-04 | 1 | Baseline |
| 25 | Order_Erysipelotrichales | 235.0487414 | Mic_Order | VE800-04 | 1 | Baseline |
| 26 | Order_Acidaminococcales | -16.2931231 | Mic_Order | VE800-04 | -1 | Baseline |
| 27 | Genus_Klebsiella | -8.7459589 | Mic_Genus | VE800-04 | -1 | Baseline |
| 28 | Genus_Subdoligranulum | -263.7348726 | Mic_Genus | VE800-04 | -1 | Baseline |
| 29 | Genus_uncl_Genus_of_Erysipelotrichale | 10.1413862 | Mic_Genus | VE800-04 | 1 | Baseline |
| 30 | Absiella.sp..MP391A4 | -1.6275147 | Mic_Species | VE800-04 | -1 | Baseline |
| 31 | Subdoligranulum.sp..MP158A7 | -18.8504211 | Mic_Species | VE800-04 | -1 | Baseline |
| 32 | Live.Cells_CD3._MAIT_TF | -0.0852210 | Immune_Cells | VE800-04 | -1 | Baseline |
| 33 | NK_CD94_IFNg_Cytokines | -0.0136499 | Immune_Cells | VE800-04 | -1 | Baseline |
| 34 | HI22.sp..HI22 | 18.8499293 | Mic_Species | VE800-05 | 1 | Baseline |
| 35 | Live.Cells_CD3.ClassMono.MDSC_TF | 0.0747047 | Immune_Cells | VE800-05 | 1 | Baseline |
| 36 | Valeric.acid | -1.7938917 | SCFA | VE800-07 | -1 | Baseline |
| 37 | Phylum_Proteobacteria | -5.6835434 | Mic_Phylum | VE800-07 | -1 | Baseline |
| 38 | Phylum_Verrucomicrobia | -0.5259962 | Mic_Phylum | VE800-07 | -1 | Baseline |
| 39 | Class_Bacteroidia | 0.0580399 | Mic_Class | VE800-07 | 1 | Baseline |
| 40 | Class_Coriobacteriia | -10.2885438 | Mic_Class | VE800-07 | -1 | Baseline |
| 41 | Class_Deltaproteobacteria | -108.8840355 | Mic_Class | VE800-07 | -1 | Baseline |
| 42 | Order_Desulfovibrionales | -83.7592013 | Mic_Order | VE800-07 | -1 | Baseline |
| 43 | Genus_Lachnoclostridium | -27.6479866 | Mic_Genus | VE800-07 | -1 | Baseline |
| 44 | Genus_Bilophila | -38.0355673 | Mic_Genus | VE800-07 | -1 | Baseline |
| 45 | X.Clostridium..glycyrrhizinilyticum | -36.5033089 | Mic_Species | VE800-07 | -1 | Baseline |
| 46 | HI22.sp..HI22 | 39.3006683 | Mic_Species | VE800-07 | 1 | Baseline |
| 47 | Bilophila.wadsworthia | -46.1505511 | Mic_Species | VE800-07 | -1 | Baseline |
| 48 | X.Ruminococcus..sp..MP10A9 | -8.4900761 | Mic_Species | VE800-07 | -1 | Baseline |
| 49 | Phylum_Actinobacteria | 3.5475678 | Mic_Phylum | VE800-08 | 1 | Baseline |
| 50 | Class_Actinobacteria | 2.1786664 | Mic_Class | VE800-08 | 1 | Baseline |
| 51 | Live.Cells_CD3.ClassMono.MDSC_TF | 0.0217676 | Immune_Cells | VE800-08 | 1 | Baseline |
| 52 | Valeric.acid | -1.8793492 | SCFA | VE800-09 | -1 | Baseline |
| 53 | Chenodeoxycholic.Acid | -0.0239468 | BA | VE800-09 | -1 | Baseline |
| 54 | Cholic.Acid | -0.0539501 | BA | VE800-09 | -1 | Baseline |
| 55 | Tauroursodeoxycholic.Acid | -0.5879278 | BA | VE800-09 | -1 | Baseline |
| 56 | Phylum_Proteobacteria | -6.2780524 | Mic_Phylum | VE800-09 | -1 | Baseline |
| 57 | Phylum_Verrucomicrobia | -0.7005082 | Mic_Phylum | VE800-09 | -1 | Baseline |
| 58 | Class_uncl_Class_of_Proteobacteria | -21.0439086 | Mic_Class | VE800-09 | -1 | Baseline |
| 59 | Class_Deltaproteobacteria | -25.3181680 | Mic_Class | VE800-09 | -1 | Baseline |
| 60 | Order_uncl_Order_of_Proteobacteria | -15.1043069 | Mic_Order | VE800-09 | -1 | Baseline |
| 61 | Order_Desulfovibrionales | -8.1621729 | Mic_Order | VE800-09 | -1 | Baseline |
| 62 | Genus_uncl_Genus_of_Erysipelotrichale | 26.8728828 | Mic_Genus | VE800-09 | 1 | Baseline |
| 63 | Bacteroides.vulgatus | 2.2013503 | Mic_Species | VE800-09 | 1 | Baseline |
| 64 | HI22.sp..HI22 | 85.6874983 | Mic_Species | VE800-09 | 1 | Baseline |
| 65 | Clostridium.sp..AT4 | -87.8206142 | Mic_Species | VE800-09 | -1 | Baseline |
| 66 | Erysipelotrichaceae.bacterium.6_1_45 | 31.9812028 | Mic_Species | VE800-09 | 1 | Baseline |
| 67 | Live.Cells_CD3.ClassMono.MDSC. | 0.0230898 | Immune_Cells | VE800-09 | 1 | Baseline |
| 68 | Butyric.acid | -0.1719479 | SCFA | VE800-10 | -1 | Baseline |
| 69 | Isobutyric.acid | -1.1871593 | SCFA | VE800-10 | -1 | Baseline |
| 70 | Isovaleric.acid | -0.4114441 | SCFA | VE800-10 | -1 | Baseline |
| 71 | Cholic.Acid | -0.0738484 | BA | VE800-10 | -1 | Baseline |
| 72 | Phylum_Proteobacteria | -2.8368296 | Mic_Phylum | VE800-10 | -1 | Baseline |
| 73 | Class_Deltaproteobacteria | -12.9711583 | Mic_Class | VE800-10 | -1 | Baseline |

| | | | | | | |
|---|---|---|---|---|---|---|
| 74 | Genus_Lachnoclostridium | -8.9739444 | Mic_Genus | VE800-10 | -1 | Baseline |
| 75 | X.Eubacterium..rectale | 13.3151464 | Mic_Species | VE800-10 | 1 | Baseline |
| 76 | Butyric.acid | -0.3990630 | SCFA | VE800-11 | -1 | Baseline |
| 77 | Chenodeoxycholic.Acid | -0.2851765 | BA | VE800-11 | -1 | Baseline |
| 78 | Phylum_Proteobacteria | -6.4893061 | Mic_Phylum | VE800-11 | -1 | Baseline |
| 79 | Phylum_Verrucomicrobia | -0.2572074 | Mic_Phylum | VE800-11 | -1 | Baseline |
| 80 | Class_uncl_Class_of_Proteobacteria | -18.8238016 | Mic_Class | VE800-11 | -1 | Baseline |
| 81 | Order_uncl_Order_of_Proteobacteria | -8.9956388 | Mic_Order | VE800-11 | -1 | Baseline |
| 82 | Genus_uncl_Genus_of_Erysipelotrichale | 37.3852724 | Mic_Genus | VE800-11 | 1 | Baseline |
| 83 | Genus_Gemmiger | 176.7416127 | Mic_Genus | VE800-11 | 1 | Baseline |
| 84 | HI22.sp..HI22 | 20.3972646 | Mic_Species | VE800-11 | 1 | Baseline |
| 85 | Clostridium.sp..AT4 | -40.1302254 | Mic_Species | VE800-11 | -1 | Baseline |
| 86 | Erysipelotrichaceae.bacterium.6_1_45 | 2.3740947 | Mic_Species | VE800-11 | 1 | Baseline |
| 87 | Gemmiger.formicilis | 126.1511706 | Mic_Species | VE800-11 | 1 | Baseline |
| 88 | Chenodeoxycholic.Acid | 0.6895750 | BA | VE800-01 | 1 | D14 |
| 89 | Lithocholic.Acid | -1.0939378 | BA | VE800-01 | -1 | D14 |
| 90 | Taurocholic.Acid | 0.5950136 | BA | VE800-01 | 1 | D14 |
| 91 | Phylum_Fusobacteria | 34.5032747 | Mic_Phylum | VE800-01 | 1 | D14 |
| 92 | Class_Negativicutes | -2.6429525 | Mic_Class | VE800-01 | -1 | D14 |
| 93 | Class_uncl_Class_of_Proteobacteria | 19.4053274 | Mic_Class | VE800-01 | 1 | D14 |
| 94 | Order_Veillonellales | -1.3208484 | Mic_Order | VE800-01 | -1 | D14 |
| 95 | Order_uncl_Order_of_Proteobacteria | 10.8206525 | Mic_Order | VE800-01 | 1 | D14 |
| 96 | Blautia.sp..VE303.05 | -97.5410295 | Mic_Species | VE800-01 | -1 | D14 |
| 97 | Acetic.acid | 0.7235153 | SCFA | VE800-02 | 1 | D14 |
| 98 | Succinic.acid | -1.9915965 | SCFA | VE800-02 | -1 | D14 |
| 99 | Class_Bacilli | 0.3565686 | Mic_Class | VE800-02 | 1 | D14 |
| 100 | Live.Cells_CD3._NonMAIT_CD4._Tbet. | 0.0224109 | Immune_Cells | VE800-02 | 1 | D14 |
| 101 | Live.Cells_CD3._IntMono_TF | -0.0229906 | Immune_Cells | VE800-02 | -1 | D14 |
| 102 | Deoxycholic.Acid | 0.2720081 | BA | VE800-03 | 1 | D14 |
| 103 | Taurochenodeoxycholic.Acid | -0.2550654 | BA | VE800-03 | -1 | D14 |
| 104 | Taurocholic.Acid | -0.1810416 | BA | VE800-03 | -1 | D14 |
| 105 | Phylum_Fusobacteria | -179.5539781 | Mic_Phylum | VE800-03 | -1 | D14 |
| 106 | Genus_uncl_Genus_of_Clostridiales | -4.3688098 | Mic_Genus | VE800-03 | -1 | D14 |
| 107 | Genus_Clostridioides | -34.8884227 | Mic_Genus | VE800-03 | -1 | D14 |
| 108 | Genus_Fusobacterium | -147.4830820 | Mic_Genus | VE800-03 | -1 | D14 |
| 109 | X.Ruminococcus..torques.CAG.61 | -35.0135853 | Mic_Species | VE800-03 | -1 | D14 |
| 110 | Fusobacterium.nucleatum | -68.4096354 | Mic_Species | VE800-03 | -1 | D14 |
| 111 | Valeric.acid | -2.4528065 | SCFA | VE800-04 | -1 | D14 |
| 112 | Cholic.Acid | -0.1258587 | BA | VE800-04 | -1 | D14 |
| 113 | Class_Actinobacteria | -1.8372229 | Mic_Class | VE800-04 | -1 | D14 |
| 114 | Order_Bifidobacteriales | -4.4648114 | Mic_Order | VE800-04 | -1 | D14 |
| 115 | Genus_Bifidobacterium | -2.7640307 | Mic_Genus | VE800-04 | -1 | D14 |
| 116 | Genus_Megasphaera | -23.7332034 | Mic_Genus | VE800-04 | -1 | D14 |
| 117 | Live.Cells_CD3._MAIT_TF | -0.3323039 | Immune_Cells | VE800-04 | -1 | D14 |
| 118 | NK_CD94_CD107a_Cytokines | -0.0275495 | Immune_Cells | VE800-04 | -1 | D14 |
| 119 | Phylum_Firmicutes | -2.2954885 | Mic_Phylum | VE800-05 | -1 | D14 |
| 120 | Phylum_Bacteroidetes | -0.8993309 | Mic_Phylum | VE800-05 | -1 | D14 |
| 121 | Genus_Roseburia | 4.7178751 | Mic_Genus | VE800-05 | 1 | D14 |
| 122 | Genus_Parasutterella | 20.1994820 | Mic_Genus | VE800-05 | 1 | D14 |
| 123 | Genus_Oscillibacter | 23.2483128 | Mic_Genus | VE800-05 | 1 | D14 |
| 124 | Live.Cells_CD3._NonMAIT_CD4. | 0.1534334 | Immune_Cells | VE800-05 | 1 | D14 |
| 125 | Live.Cells_CD3._CD19._TF | 0.0133487 | Immune_Cells | VE800-05 | 1 | D14 |
| 126 | CD4_IL17_Cytokines | 0.0342989 | Immune_Cells | VE800-05 | 1 | D14 |
| 127 | Succinic.acid | 1.7699717 | SCFA | VE800-07 | 1 | D14 |
| 128 | Ursodeoxycholic.Acid | -0.3755578 | BA | VE800-07 | -1 | D14 |
| 129 | Phylum_Actinobacteria | -2.9184215 | Mic_Phylum | VE800-07 | -1 | D14 |
| 130 | Class_Actinobacteria | -8.7044958 | Mic_Class | VE800-07 | -1 | D14 |
| 131 | Class_Gammaproteobacteria | -7.0238298 | Mic_Class | VE800-07 | -1 | D14 |
| 132 | Class_Betaproteobacteria | 15.3703428 | Mic_Class | VE800-07 | 1 | D14 |
| 133 | Class_Deltaproteobacteria | -2.5448595 | Mic_Class | VE800-07 | -1 | D14 |
| 134 | Order_Bifidobacteriales | -9.4950733 | Mic_Order | VE800-07 | -1 | D14 |
| 135 | Order_Enterobacterales | -8.3259981 | Mic_Order | VE800-07 | -1 | D14 |
| 136 | Order_Veillonellales | -0.9454855 | Mic_Order | VE800-07 | -1 | D14 |
| 137 | Order_Burkholderiales | 18.3818290 | Mic_Order | VE800-07 | 1 | D14 |
| 138 | Order_Desulfovibrionales | -7.9263011 | Mic_Order | VE800-07 | -1 | D14 |
| 139 | Streptococcus.thermophilus | -6.3641926 | Mic_Species | VE800-07 | -1 | D14 |
| 140 | Lactobacillus.plantarum | -2.5728846 | Mic_Species | VE800-07 | -1 | D14 |
| 141 | Live.Cells_CD3.ClassMono.MDSC. | -0.0319453 | Immune_Cells | VE800-07 | -1 | D14 |
| 142 | X2.Methylbutyric.acid | 0.2318423 | SCFA | VE800-08 | 1 | D14 |
| 143 | Genus_uncl_Genus_of_Clostridia | 25.5488544 | Mic_Genus | VE800-08 | 1 | D14 |
| 144 | Propionic.acid | -0.1608577 | SCFA | VE800-09 | -1 | D14 |
| 145 | Succinic.acid | 0.8568948 | SCFA | VE800-09 | 1 | D14 |
| 146 | Phylum_Actinobacteria | -2.1799847 | Mic_Phylum | VE800-09 | -1 | D14 |
| 147 | Class_Bacilli | -0.7523875 | Mic_Class | VE800-09 | -1 | D14 |
| 148 | Class_Actinobacteria | -7.5547630 | Mic_Class | VE800-09 | -1 | D14 |

| | | | | | | |
|---|---|---|---|---|---|---|
| 149 | Class_Betaproteobacteria | 31.8621575 | Mic_Class | VE800-09 | 1 | D14 |
| 150 | Class_uncl_Class_of_Proteobacteria | -102.1481750 | Mic_Class | VE800-09 | -1 | D14 |
| 151 | Order_Bifidobacteriales | -1.6731688 | Mic_Order | VE800-09 | -1 | D14 |
| 152 | Order_Burkholderiales | 18.7967071 | Mic_Order | VE800-09 | 1 | D14 |
| 153 | Order_uncl_Order_of_Proteobacteria | -59.1432861 | Mic_Order | VE800-09 | -1 | D14 |
| 154 | Live.Cells_CD3. | 0.0003434 | Immune_Cells | VE800-09 | 1 | D14 |
| 155 | CD4_TNF_Cytokines | 0.0061740 | Immune_Cells | VE800-09 | 1 | D14 |
| 156 | Propionic.acid | -1.6908870 | SCFA | VE800-10 | -1 | D14 |
| 157 | Valeric.acid | -7.3305659 | SCFA | VE800-10 | -1 | D14 |
| 158 | Phylum_Firmicutes | -0.3311927 | Mic_Phylum | VE800-11 | -1 | D14 |
| 159 | Class_uncl_Class_of_Proteobacteria | -60.7113891 | Mic_Class | VE800-11 | -1 | D14 |
| 160 | Order_uncl_Order_of_Proteobacteria | -60.7113891 | Mic_Order | VE800-11 | -1 | D14 |
| 161 | Proteobacteria.bacterium.CAG.139 | -39.7834251 | Mic_Species | VE800-11 | -1 | D14 |
| 162 | CD3._Cytokines | 0.0034865 | Immune_Cells | VE800-11 | 1 | D14 |
| 163 | Chenodeoxycholic.Acid | 1.2097499 | BA | VE800-01 | 1 | D30 |
| 164 | Live.Cells_CD3.ClassMono.MDSC. | 0.0508417 | Immune_Cells | VE800-01 | 1 | D30 |
| 165 | Succinic.acid | -3.4611125 | SCFA | VE800-02 | -1 | D30 |
| 166 | Glycoursodeoxycholic.Acid | 0.2705054 | BA | VE800-02 | 1 | D30 |
| 167 | Taurodeoxycholic.Acid | 0.5659376 | BA | VE800-02 | 1 | D30 |
| 168 | Ursodeoxycholic.Acid | 2.9736282 | BA | VE800-02 | 1 | D30 |
| 169 | Phylum_Actinobacteria | 0.0695439 | Mic_Phylum | VE800-02 | 1 | D30 |
| 170 | Phylum_Fusobacteria | -13.3995031 | Mic_Phylum | VE800-02 | -1 | D30 |
| 171 | Class_Erysipelotrichia | 5.0697385 | Mic_Class | VE800-02 | 1 | D30 |
| 172 | Class_Deltaproteobacteria | -1.1084389 | Mic_Class | VE800-02 | -1 | D30 |
| 173 | Order_Erysipelotrichales | 5.0697385 | Mic_Order | VE800-02 | 1 | D30 |
| 174 | Order_Desulfovibrionales | -1.1084389 | Mic_Order | VE800-02 | -1 | D30 |
| 175 | Glycolithocholic.Acid | 0.2069606 | BA | VE800-03 | 1 | D30 |
| 176 | Phylum_Bacteroidetes | -2.2674215 | Mic_Phylum | VE800-03 | -1 | D30 |
| 177 | Class_Bacteroidia | -1.1393549 | Mic_Class | VE800-03 | -1 | D30 |
| 178 | Class_Erysipelotrichia | 1.6450286 | Mic_Class | VE800-03 | 1 | D30 |
| 179 | Genus_Flavonifractor | 5.4898524 | Mic_Genus | VE800-03 | 1 | D30 |
| 180 | Genus_uncl_Genus_of_Erysipelotrichale | 36.5368752 | Mic_Genus | VE800-03 | 1 | D30 |
| 181 | Blautia.sp..HF29 | 58.2451042 | Mic_Species | VE800-03 | 1 | D30 |
| 182 | Isovaleric.acid | -0.5454388 | SCFA | VE800-04 | -1 | D30 |
| 183 | Valeric.acid | -0.9442886 | SCFA | VE800-04 | -1 | D30 |
| 184 | Cholic.Acid | -0.5112912 | BA | VE800-04 | -1 | D30 |
| 185 | Taurodeoxycholic.Acid | 0.5183046 | BA | VE800-04 | 1 | D30 |
| 186 | Class_Negativicutes | -18.8693544 | Mic_Class | VE800-04 | -1 | D30 |
| 187 | Class_Deltaproteobacteria | -1.1910164 | Mic_Class | VE800-04 | -1 | D30 |
| 188 | Order_Veillonellales | -15.9383997 | Mic_Order | VE800-04 | -1 | D30 |
| 189 | Order_Desulfovibrionales | -47.3964213 | Mic_Order | VE800-04 | -1 | D30 |
| 190 | Order_Acidaminococcales | -61.9730576 | Mic_Order | VE800-04 | -1 | D30 |
| 191 | Genus_Parabacteroides | -3.1230659 | Mic_Genus | VE800-04 | -1 | D30 |
| 192 | Genus_Megasphaera | -10.7459177 | Mic_Genus | VE800-04 | -1 | D30 |
| 193 | Genus_Bilophila | -42.2182883 | Mic_Genus | VE800-04 | -1 | D30 |
| 194 | Genus_Phascolarctobacterium | -56.2385277 | Mic_Genus | VE800-04 | -1 | D30 |
| 195 | Live.Cells_CD3._MAIT_TF | -0.0587303 | Immune_Cells | VE800-04 | -1 | D30 |
| 196 | Phylum_Actinobacteria | 2.1105293 | Mic_Phylum | VE800-05 | 1 | D30 |
| 197 | Phylum_Verrucomicrobia | 6.5883307 | Mic_Phylum | VE800-05 | 1 | D30 |
| 198 | Live.Cells_CD3._NonMAIT_CD4. | 0.0116574 | Immune_Cells | VE800-05 | 1 | D30 |
| 199 | All_PBMCs_Stim_Cytokines | -0.0803077 | Immune_Cells | VE800-05 | -1 | D30 |
| 200 | Valeric.acid | -0.6655751 | SCFA | VE800-07 | -1 | D30 |
| 201 | Class_Clostridia | 0.3011555 | Mic_Class | VE800-07 | 1 | D30 |
| 202 | Class_Actinobacteria | -2.9291417 | Mic_Class | VE800-07 | -1 | D30 |
| 203 | Class_Deltaproteobacteria | -60.5979816 | Mic_Class | VE800-07 | -1 | D30 |
| 204 | Genus_Flavonifractor | 27.9913398 | Mic_Genus | VE800-07 | 1 | D30 |
| 205 | Genus_Parasutterella | 113.2815469 | Mic_Genus | VE800-07 | 1 | D30 |
| 206 | Glycocholic.Acid | -1.6469290 | BA | VE800-08 | -1 | D30 |
| 207 | Phylum_Verrucomicrobia | 4.0845936 | Mic_Phylum | VE800-08 | 1 | D30 |
| 208 | Live.Cells_CD3._NonMAIT_CD4. | 0.0125699 | Immune_Cells | VE800-08 | 1 | D30 |
| 209 | Valeric.acid | -3.2860095 | SCFA | VE800-09 | -1 | D30 |
| 210 | Phylum_Bacteroidetes | -0.2353777 | Mic_Phylum | VE800-09 | -1 | D30 |
| 211 | Phylum_Fusobacteria | -12.7774074 | Mic_Phylum | VE800-09 | -1 | D30 |
| 212 | Phylum_Bacteroidetes | -0.6832097 | Mic_Phylum | VE800-10 | -1 | D30 |
| 213 | Class_Coriobacteriia | 11.6799630 | Mic_Class | VE800-10 | 1 | D30 |
| 214 | Genus_uncl_Genus_of_Erysipelotrichale | 12.3124975 | Mic_Genus | VE800-10 | 1 | D30 |
| 215 | Genus_Fusicatenibacter | -7.4766422 | Mic_Genus | VE800-10 | -1 | D30 |
| 216 | Valeric.acid | -3.2860095 | SCFA | VE800-11 | -1 | D30 |
| 217 | Class_Bacteroidia | -0.0940513 | Mic_Class | VE800-11 | -1 | D30 |
| 218 | Class_uncl_Class_of_Proteobacteria | -9.0553709 | Mic_Class | VE800-11 | -1 | D30 |

#### Supplementary Table S15: Serum SCFA Changes in Melanoma Cohort

| | contrast | Response_Wk8 | estimate | SE | df | t.ratio | p.value | Analyte | p_adj |
|---|---|---|---|---|---|---|---|---|---|
| 1 | Baseline - D1 | SD | -0.03139507 | 0.10398180 | 40 | -0.30192851 | 0.99029162 | 2-Methylbutyrate | 0.993170484 |
| 2 | Baseline - D14 | SD | -0.08211533 | 0.10398180 | 40 | -0.78970864 | 0.85868715 | 2-Methylbutyrate | 0.999675891 |
| 3 | Baseline - D30 | SD | 0.03705211 | 0.10398180 | 40 | 0.35633266 | 0.98426721 | 2-Methylbutyrate | 0.999763662 |
| 4 | Baseline - D1 | PD | 0.07399739 | 0.09803498 | 40 | 0.75480595 | 0.87407080 | 2-Methylbutyrate | 0.988682031 |
| 5 | Baseline - D14 | PD | 0.16384566 | 0.10160910 | 40 | 1.61250966 | 0.38332125 | 2-Methylbutyrate | 0.969732218 |
| 6 | Baseline - D30 | PD | 0.16457133 | 0.11906969 | 40 | 1.38214291 | 0.51764817 | 2-Methylbutyrate | 0.999997504 |
| 7 | Baseline - D1 | SD | -0.03337801 | 0.12462319 | 40 | -0.26783142 | 0.99317048 | Acetate | 0.993170484 |
| 8 | Baseline - D14 | SD | 0.05980136 | 0.12462319 | 40 | 0.47985740 | 0.96311565 | Acetate | 0.999675891 |
| 9 | Baseline - D30 | SD | 0.01078389 | 0.12462319 | 40 | 0.08653196 | 0.99976366 | Acetate | 0.999763662 |
| 10 | Baseline - D1 | PD | -0.06125270 | 0.11749587 | 40 | -0.52131786 | 0.95346890 | Acetate | 0.988682031 |
| 11 | Baseline - D14 | PD | -0.11513008 | 0.12166388 | 40 | -0.94629633 | 0.78017359 | Acetate | 0.999998293 |
| 12 | Baseline - D30 | PD | -0.08138280 | 0.14207366 | 40 | -0.57282115 | 0.93961267 | Acetate | 0.999997504 |
| 13 | Baseline - D1 | SD | -0.04384898 | 0.05262363 | 40 | -0.83325648 | 0.83833829 | Butyrate | 0.993170484 |
| 14 | Baseline - D14 | SD | 0.00506033 | 0.05262363 | 40 | 0.09616082 | 0.99967589 | Butyrate | 0.999675891 |
| 15 | Baseline - D30 | SD | -0.06369990 | 0.05262363 | 40 | -1.21048092 | 0.62393571 | Butyrate | 0.999763662 |
| 16 | Baseline - D1 | PD | 0.04443084 | 0.04961403 | 40 | 0.89552962 | 0.80718086 | Butyrate | 0.988682031 |
| 17 | Baseline - D14 | PD | -0.11759467 | 0.05142132 | 40 | -2.28688551 | 0.11821969 | Butyrate | 0.945757523 |
| 18 | Baseline - D30 | PD | -0.00552130 | 0.06025105 | 40 | -0.09163821 | 0.99971941 | Butyrate | 0.999997504 |
| 19 | Baseline - D1 | SD | -0.06744955 | 0.05398044 | 40 | -1.24951845 | 0.59969820 | Isobutyrate | 0.993170484 |
| 20 | Baseline - D14 | SD | -0.05843944 | 0.05398044 | 40 | -1.08260399 | 0.70201535 | Isobutyrate | 0.999675891 |
| 21 | Baseline - D30 | SD | -0.06324839 | 0.05398044 | 40 | -1.17169091 | 0.64790287 | Isobutyrate | 0.999763662 |
| 22 | Baseline - D1 | PD | 0.05195232 | 0.05089324 | 40 | 1.02080981 | 0.73832338 | Isobutyrate | 0.988682031 |
| 23 | Baseline - D14 | PD | 0.03299224 | 0.05282167 | 40 | 0.62459672 | 0.92357197 | Isobutyrate | 0.999998293 |
| 24 | Baseline - D30 | PD | 0.01118859 | 0.06221065 | 40 | 0.17985010 | 0.99789915 | Isobutyrate | 0.999997504 |
| 25 | Baseline - D1 | SD | -0.09396359 | 0.08545421 | 40 | -1.09957813 | 0.69184072 | Isovalerate | 0.993170484 |
| 26 | Baseline - D14 | SD | -0.04566506 | 0.08545421 | 40 | -0.53438047 | 0.95015226 | Isovalerate | 0.999675891 |
| 27 | Baseline - D30 | SD | -0.06158511 | 0.08545421 | 40 | -0.72067965 | 0.88827473 | Isovalerate | 0.999763662 |
| 28 | Baseline - D1 | PD | 0.04235299 | 0.08056701 | 40 | 0.52568657 | 0.95237456 | Isovalerate | 0.988682031 |
| 29 | Baseline - D14 | PD | 0.12013005 | 0.08364572 | 40 | 1.43617679 | 0.48486611 | Isovalerate | 0.969732218 |
| 30 | Baseline - D30 | PD | -0.00187058 | 0.09862381 | 40 | -0.01896680 | 0.99999750 | Isovalerate | 0.999997504 |
| 31 | Baseline - D1 | SD | -0.04507296 | 0.05011703 | 40 | -0.89935419 | 0.80519390 | Propionate | 0.993170484 |
| 32 | Baseline - D14 | SD | -0.03869238 | 0.05011703 | 40 | -0.77204064 | 0.86658045 | Propionate | 0.999675891 |
| 33 | Baseline - D30 | SD | -0.05103343 | 0.05011703 | 40 | -1.01828516 | 0.73977889 | Propionate | 0.999763662 |
| 34 | Baseline - D1 | PD | 0.04987756 | 0.04725079 | 40 | 1.05559220 | 0.71803874 | Propionate | 0.988682031 |
| 35 | Baseline - D14 | PD | -0.04079381 | 0.04904356 | 40 | -0.83178725 | 0.83904490 | Propionate | 0.999998293 |
| 36 | Baseline - D30 | PD | 0.00418341 | 0.05777105 | 40 | 0.07241361 | 0.99986137 | Propionate | 0.999997504 |
| 37 | Baseline - D1 | SD | -0.01913013 | 0.04241371 | 40 | -0.45103656 | 0.96904522 | Succinate | 0.993170484 |
| 38 | Baseline - D14 | SD | 0.02278798 | 0.04241371 | 40 | 0.53727856 | 0.94939827 | Succinate | 0.999675891 |
| 39 | Baseline - D30 | SD | 0.04158340 | 0.04241371 | 40 | 0.98042347 | 0.76130406 | Succinate | 0.999763662 |
| 40 | Baseline - D1 | PD | -0.01272401 | 0.03998803 | 40 | -0.31819541 | 0.98868203 | Succinate | 0.988682031 |
| 41 | Baseline - D14 | PD | 0.00069412 | 0.04153599 | 40 | 0.01671123 | 0.99999829 | Succinate | 0.999998293 |
| 42 | Baseline - D30 | PD | 0.05024024 | 0.04905809 | 40 | 1.02409686 | 0.73642477 | Succinate | 0.999997504 |
| 43 | Baseline - D1 | SD | -0.01461571 | 0.02513570 | 40 | -0.58147205 | 0.93707986 | Valerate | 0.993170484 |
| 44 | Baseline - D14 | SD | 0.02726665 | 0.02513570 | 40 | 1.08477779 | 0.70071663 | Valerate | 0.999675891 |
| 45 | Baseline - D30 | SD | -0.00958608 | 0.02513570 | 40 | -0.38137300 | 0.98085292 | Valerate | 0.999763662 |
| 46 | Baseline - D1 | PD | 0.01373445 | 0.02369817 | 40 | 0.57955763 | 0.93764547 | Valerate | 0.988682031 |
| 47 | Baseline - D14 | PD | -0.04044900 | 0.02460796 | 40 | -1.64373661 | 0.36646476 | Valerate | 0.969732218 |
| 48 | Baseline - D30 | PD | -0.00766801 | 0.02903227 | 40 | -0.26412024 | 0.99344529 | Valerate | 0.999997504 |

#### Supplementary Table S16: Serum Bile Acid Changes in Melanoma Cohort

| | contrast | Response_ Wk8 | estimate | SE | df | t.ratio | p.value | Analyte | p_adj |
|---|---|---|---|---|---|---|---|---|---|
| 17 | Baseline - D14 | PD | 0.97907022 | 0.19470796 | 40 | 5.0284037 | 0.0000621 | Deoxycholic Acid | 0.0008697 |
| 35 | Baseline - D14 | PD | 0.77983521 | 0.19624037 | 40 | 3.9738775 | 0.0015792 | Glycodeoxycholic Acid | 0.0110541 |
| 16 | Baseline - D1 | PD | 0.76326315 | 0.18783454 | 40 | 4.0634867 | 0.0012119 | Deoxycholic Acid | 0.0169660 |
| 40 | Baseline - D1 | PD | 0.46298187 | 0.12708451 | 40 | 3.6431022 | 0.0041010 | Glycolithocholic Acid | 0.0287071 |
| 41 | Baseline - D14 | PD | 0.43067208 | 0.13099868 | 40 | 3.2876062 | 0.0109055 | Glycolithocholic Acid | 0.0416811 |
| 71 | Baseline - D14 | PD | 0.49348253 | 0.15162800 | 40 | 3.2545607 | 0.0119089 | Taurodeoxycholic Acid | 0.0416811 |
| 47 | Baseline - D14 | PD | -0.49297100 | 0.16687419 | 40 | -2.9541476 | 0.0258326 | Glycoursodeoxycholic Acid | 0.0723313 |
| 23 | Baseline - D14 | PD | -0.29007968 | 0.12504310 | 40 | -2.3198375 | 0.1104906 | Glycochenodeoxycholic Acid | 0.2578114 |
| 83 | Baseline - D14 | PD | -0.37356408 | 0.18618376 | 40 | -2.0064268 | 0.2027103 | Ursodeoxycholic Acid | 0.4054205 |
| 82 | Baseline - D1 | PD | 0.41486771 | 0.17929471 | 40 | 2.3138871 | 0.1118550 | Ursodeoxycholic Acid | 0.5219901 |
| 77 | Baseline - D14 | PD | -0.23699646 | 0.13679830 | 40 | -1.7324518 | 0.3207899 | Tauroursodeoxycholic Acid | 0.5613823 |
| 58 | Baseline - D1 | PD | -0.32022532 | 0.15434871 | 40 | -2.0746873 | 0.1788793 | Taurochenodeoxycholic Acid | 0.6260777 |
| 22 | Baseline - D1 | PD | -0.21393633 | 0.12035103 | 40 | -1.7776028 | 0.2988819 | Glycochenodeoxycholic Acid | 0.7551253 |
| 52 | Baseline - D1 | PD | 0.16731680 | 0.09689726 | 40 | 1.7267444 | 0.3236251 | Lithocholic Acid | 0.7551253 |
| 5 | Baseline - D14 | PD | -0.31606940 | 0.22817242 | 40 | -1.3852218 | 0.5157657 | Chenodeoxycholic Acid | 0.8023021 |
| 12 | Baseline - D30 | PD | -0.59651814 | 0.28128865 | 40 | -2.1206619 | 0.1640526 | Cholic Acid | 0.8342372 |
| 18 | Baseline - D30 | PD | 0.43760929 | 0.22827575 | 40 | 1.9170205 | 0.2372880 | Deoxycholic Acid | 0.8342372 |
| 36 | Baseline - D30 | PD | 0.44128752 | 0.23050774 | 40 | 1.9144151 | 0.2383535 | Glycodeoxycholic Acid | 0.8342372 |
| 72 | Baseline - D30 | PD | 0.38502657 | 0.17861849 | 40 | 2.1555806 | 0.1534328 | Taurodeoxycholic Acid | 0.8342372 |
| 84 | Baseline - D30 | PD | -0.38837251 | 0.21968275 | 40 | -1.7678790 | 0.3035210 | Ursodeoxycholic Acid | 0.8498589 |
| 6 | Baseline - D30 | PD | -0.36598403 | 0.26741668 | 40 | -1.3685909 | 0.5259523 | Chenodeoxycholic Acid | 0.8777936 |
| 42 | Baseline - D30 | PD | 0.23159685 | 0.15038496 | 40 | 1.5400267 | 0.4238604 | Glycolithocholic Acid | 0.8777936 |
| 48 | Baseline - D30 | PD | -0.25777323 | 0.19730150 | 40 | -1.3064940 | 0.5642959 | Glycoursodeoxycholic Acid | 0.8777936 |
| 60 | Baseline - D30 | PD | 0.27736188 | 0.18845471 | 40 | 1.4717694 | 0.4636225 | Taurochenodeoxycholic Acid | 0.8777936 |
| 66 | Baseline - D30 | PD | 0.23907666 | 0.20115785 | 40 | 1.1885028 | 0.6375353 | Taurocholic Acid | 0.8925495 |
| 53 | Baseline - D14 | PD | 0.10586412 | 0.10008374 | 40 | 1.0577554 | 0.7167637 | Lithocholic Acid | 0.9122447 |
| 59 | Baseline - D14 | PD | -0.17114716 | 0.16015725 | 40 | -1.0686195 | 0.7103382 | Taurochenodeoxycholic Acid | 0.9122447 |
| 10 | Baseline - D1 | PD | 0.27088312 | 0.23069736 | 40 | 1.1741925 | 0.6463625 | Cholic Acid | 0.9621089 |
| 34 | Baseline - D1 | PD | 0.23328534 | 0.18921475 | 40 | 1.2329131 | 0.6100167 | Glycodeoxycholic Acid | 0.9621089 |
| 46 | Baseline - D1 | PD | 0.17783265 | 0.16060816 | 40 | 1.1072454 | 0.6872207 | Glycoursodeoxycholic Acid | 0.9621089 |
| 64 | Baseline - D1 | PD | -0.22211079 | 0.16453712 | 40 | -1.3499129 | 0.5374406 | Taurocholic Acid | 0.9621089 |
| 24 | Baseline - D30 | PD | -0.07775108 | 0.14782887 | 40 | -0.5259533 | 0.9523073 | Glycochenodeoxycholic Acid | 0.9941260 |
| 30 | Baseline - D30 | PD | 0.04331183 | 0.17020299 | 40 | 0.2544716 | 0.9941260 | Glycocholic Acid | 0.9941260 |
| 54 | Baseline - D30 | PD | 0.09782705 | 0.11578520 | 40 | 0.8449012 | 0.8326899 | Lithocholic Acid | 0.9941260 |
| 78 | Baseline - D30 | PD | 0.07005571 | 0.16053385 | 40 | 0.4363921 | 0.9718194 | Tauroursodeoxycholic Acid | 0.9941260 |
| 4 | Baseline - D1 | PD | -0.07109293 | 0.22013855 | 40 | -0.3229463 | 0.9881818 | Chenodeoxycholic Acid | 0.9965679 |
| 28 | Baseline - D1 | PD | -0.11053822 | 0.13875712 | 40 | -0.7966310 | 0.8555364 | Glycocholic Acid | 0.9965679 |
| 70 | Baseline - D1 | PD | 0.03099600 | 0.14608353 | 40 | 0.2121800 | 0.9965679 | Taurodeoxycholic Acid | 0.9965679 |
| 76 | Baseline - D1 | PD | 0.04257014 | 0.13193506 | 40 | 0.3226598 | 0.9882124 | Tauroursodeoxycholic Acid | 0.9965679 |
| 11 | Baseline - D14 | PD | -0.05046388 | 0.23930830 | 40 | -0.2108739 | 0.9966301 | Cholic Acid | 0.9989754 |
| 29 | Baseline - D14 | PD | -0.06891377 | 0.14412352 | 40 | -0.4781577 | 0.9634828 | Glycocholic Acid | 0.9989754 |
| 65 | Baseline - D14 | PD | -0.02413664 | 0.17077753 | 40 | -0.1413338 | 0.9989754 | Taurocholic Acid | 0.9989754 |
| 14 | Baseline - D14 | SD | 0.70581655 | 0.19922862 | 40 | 3.5427468 | 0.0054346 | Deoxycholic Acid | 0.0289994 |
| 32 | Baseline - D14 | SD | 0.74907171 | 0.20069255 | 40 | 3.7324340 | 0.0031814 | Glycodeoxycholic Acid | 0.0289994 |
| 68 | Baseline - D14 | SD | 0.54144690 | 0.15494499 | 40 | 3.4944461 | 0.0062142 | Taurodeoxycholic Acid | 0.0289994 |
| 49 | Baseline - D1 | SD | 0.38292461 | 0.10277506 | 40 | 3.7258514 | 0.0032418 | Lithocholic Acid | 0.0453853 |
| 37 | Baseline - D1 | SD | 0.44691321 | 0.13479348 | 40 | 3.3155402 | 0.0101194 | Glycolithocholic Acid | 0.0708361 |
| 13 | Baseline - D1 | SD | 0.62671012 | 0.19922862 | 40 | 3.1456832 | 0.0158543 | Deoxycholic Acid | 0.0739868 |
| 79 | Baseline - D1 | SD | 0.53709220 | 0.19017076 | 40 | 2.8242628 | 0.0355441 | Ursodeoxycholic Acid | 0.1244044 |
| 38 | Baseline - D14 | SD | 0.35773023 | 0.13479348 | 40 | 2.6539134 | 0.0531669 | Glycolithocholic Acid | 0.1860840 |

| | | | | | | | | | |
|---|---|---|---|---|---|---|---|---|---|
| 44 | Baseline - D14 | SD | -0.40096677 | 0.17035068 | 40 | -2.3537726 | 0.1029670 | Glycoursodeoxycholic Acid | 0.2697555 |
| 50 | Baseline - D14 | SD | 0.23615786 | 0.10277506 | 40 | 2.2978129 | 0.1156095 | Lithocholic Acid | 0.2697555 |
| 3 | Baseline - D30 | SD | -0.58135946 | 0.23349219 | 40 | -2.4898454 | 0.0769245 | Chenodeoxycholic Acid | 0.4179828 |
| 9 | Baseline - D30 | SD | -0.56667770 | 0.24469150 | 40 | -2.3158863 | 0.1113951 | Cholic Acid | 0.4179828 |
| 21 | Baseline - D30 | SD | -0.27697140 | 0.12765155 | 40 | -2.1697457 | 0.1492796 | Glycochenodeoxycholic Acid | 0.4179828 |
| 51 | Baseline - D30 | SD | 0.26921076 | 0.10277506 | 40 | 2.6194171 | 0.0575499 | Lithocholic Acid | 0.4179828 |
| 69 | Baseline - D30 | SD | 0.34246515 | 0.15494499 | 40 | 2.2102371 | 0.1378898 | Taurodeoxycholic Acid | 0.4179828 |
| 20 | Baseline - D14 | SD | -0.25259185 | 0.12765155 | 40 | -1.9787606 | 0.2129992 | Glycochenodeoxycholic Acid | 0.4259984 |
| 15 | Baseline - D30 | SD | 0.40197816 | 0.19922862 | 40 | 2.0176728 | 0.1986325 | Deoxycholic Acid | 0.4634758 |
| 55 | Baseline - D1 | SD | -0.32775514 | 0.16371153 | 40 | -2.0020284 | 0.2043215 | Taurochenodeoxycholic Acid | 0.4767502 |
| 61 | Baseline - D1 | SD | -0.35067172 | 0.17451797 | 40 | -2.0093732 | 0.2016361 | Taurocholic Acid | 0.4767502 |
| 25 | Baseline - D1 | SD | -0.26770370 | 0.14717415 | 40 | -1.8189587 | 0.2796448 | Glycocholic Acid | 0.5592897 |
| 1 | Baseline - D1 | SD | -0.38555891 | 0.23349219 | 40 | -1.6512711 | 0.3624562 | Chenodeoxycholic Acid | 0.5638207 |
| 19 | Baseline - D1 | SD | -0.21832070 | 0.12765155 | 40 | -1.7102864 | 0.3318822 | Glycochenodeoxycholic Acid | 0.5638207 |
| 33 | Baseline - D30 | SD | 0.27039630 | 0.20069255 | 40 | 1.3473161 | 0.5390414 | Glycodeoxycholic Acid | 0.7546580 |
| 39 | Baseline - D30 | SD | 0.20100471 | 0.13479348 | 40 | 1.4912050 | 0.4521611 | Glycolithocholic Acid | 0.7546580 |
| 45 | Baseline - D30 | SD | -0.26251011 | 0.17035068 | 40 | -1.5409983 | 0.4233049 | Glycoursodeoxycholic Acid | 0.7546580 |
| 57 | Baseline - D30 | SD | -0.23478966 | 0.16371153 | 40 | -1.4341669 | 0.4860748 | Taurochenodeoxycholic Acid | 0.7546580 |
| 27 | Baseline - D30 | SD | -0.17677454 | 0.14717415 | 40 | -1.2011249 | 0.6297305 | Glycocholic Acid | 0.8014752 |
| 2 | Baseline - D14 | SD | -0.34482791 | 0.23349219 | 40 | -1.4768284 | 0.4606292 | Chenodeoxycholic Acid | 0.8061011 |
| 63 | Baseline - D30 | SD | -0.12641628 | 0.17451797 | 40 | -0.7243740 | 0.8867781 | Taurocholic Acid | 0.9107773 |
| 75 | Baseline - D30 | SD | -0.13197713 | 0.13993826 | 40 | -0.9431097 | 0.7819076 | Tauroursodeoxycholic Acid | 0.9107773 |
| 81 | Baseline - D30 | SD | -0.12583850 | 0.19017076 | 40 | -0.6617132 | 0.9107773 | Ursodeoxycholic Acid | 0.9107773 |
| 7 | Baseline - D1 | SD | -0.19897702 | 0.24469150 | 40 | -0.8131750 | 0.8478765 | Cholic Acid | 0.9316600 |
| 31 | Baseline - D1 | SD | 0.19021255 | 0.20069255 | 40 | 0.9477808 | 0.7793641 | Glycodeoxycholic Acid | 0.9316600 |
| 43 | Baseline - D1 | SD | 0.13208351 | 0.17035068 | 40 | 0.7753624 | 0.8651129 | Glycoursodeoxycholic Acid | 0.9316600 |
| 73 | Baseline - D1 | SD | 0.11502998 | 0.13993826 | 40 | 0.8220052 | 0.8437144 | Tauroursodeoxycholic Acid | 0.9316600 |
| 8 | Baseline - D14 | SD | -0.13599012 | 0.24469150 | 40 | -0.5557615 | 0.9444340 | Cholic Acid | 0.9444340 |
| 26 | Baseline - D14 | SD | -0.15436012 | 0.14717415 | 40 | -1.0488263 | 0.7220169 | Glycocholic Acid | 0.9444340 |
| 56 | Baseline - D14 | SD | -0.16588722 | 0.16371153 | 40 | -1.0132898 | 0.7426517 | Taurochenodeoxycholic Acid | 0.9444340 |
| 62 | Baseline - D14 | SD | -0.12770285 | 0.17451797 | 40 | -0.7317461 | 0.8837615 | Taurocholic Acid | 0.9444340 |
| 74 | Baseline - D14 | SD | -0.10948033 | 0.13993826 | 40 | -0.7823473 | 0.8620019 | Tauroursodeoxycholic Acid | 0.9444340 |
| 80 | Baseline - D14 | SD | -0.11171144 | 0.19017076 | 40 | -0.5874270 | 0.9353020 | Ursodeoxycholic Acid | 0.9444340 |
| 67 | Baseline - D1 | SD | 0.02706684 | 0.15494499 | 40 | 0.1746868 | 0.9980735 | Taurodeoxycholic Acid | 0.9980735 |

#### Supplementary Table S17: Serum Cytokine LME in Melanoma Cohort

| | contrast | Time | estimate | SE | df | t.ratio | p.value | Cytokines | p_adj |
|---|---|---|---|---|---|---|---|---|---|
| 48 | HC - LC | D30 | 1.9750165 | 0.6173977 | 14 | 3.198937199 | 0.006434126 | IL-21 | 0.215317238 |
| 192 | HC - LC | D30 | 1.0422433 | 0.3426195 | 14 | 3.041985107 | 0.008788459 | TRAIL | 0.215317238 |
| 64 | HC - LC | D30 | 0.7847390 | 0.2870051 | 14 | 2.734233157 | 0.016136701 | IL-17F | 0.263566113 |
| 7 | HC - LC | D14 | 0.5624485 | 0.2304214 | 14 | 2.44095635 | 0.028535839 | Eotaxin | 0.453976956 |
| 47 | HC - LC | D14 | 1.3291254 | 0.5780533 | 14 | 2.299312979 | 0.03739896 | IL-21 | 0.453976956 |
| 88 | HC - LC | D30 | 0.3972051 | 0.1744241 | 14 | 2.277236824 | 0.038995592 | IL-9 | 0.355801605 |
| 16 | HC - LC | D30 | 0.5823164 | 0.2563880 | 14 | 2.271230912 | 0.039440944 | FLT-3L | 0.355801605 |
| 54 | HC - LC | D1 | 0.8545335 | 0.3820454 | 14 | 2.236732832 | 0.042093071 | IL-17A | 0.888145567 |
| 148 | HC - LC | D30 | 0.4600318 | 0.2106129 | 14 | 2.184252962 | 0.046450004 | MDC | 0.355801605 |
| 171 | HC - LC | D14 | 0.6281018 | 0.2897104 | 14 | 2.168033321 | 0.047879552 | RANTES | 0.453976956 |
| 63 | HC - LC | D14 | 0.5769713 | 0.2741249 | 14 | 2.104775601 | 0.053855064 | IL-17F | 0.453976956 |
| 191 | HC - LC | D14 | 0.6332054 | 0.3008938 | 14 | 2.104414557 | 0.053891071 | TRAIL | 0.453976956 |
| 75 | HC - LC | D14 | 0.4132673 | 0.1979593 | 14 | 2.08763769 | 0.055589015 | IL-12 (p40) | 0.453976956 |
| 132 | HC - LC | D30 | -1.1124265 | 0.5345502 | 14 | -2.081051629 | 0.05626898 | IP-10 | 0.355801605 |
| 164 | HC - LC | D30 | 0.5791130 | 0.2806096 | 14 | 2.063767964 | 0.058090058 | PDGF-AA | 0.355801605 |
| 151 | HC - LC | D14 | 0.3877321 | 0.1966250 | 14 | 1.971936332 | 0.06870713 | MIG | 0.476537786 |
| 87 | HC - LC | D14 | 0.2947139 | 0.1556845 | 14 | 1.893020552 | 0.079212809 | IL-9 | 0.476537786 |
| 154 | HC - LC | D1 | 0.6652421 | 0.3586521 | 14 | 1.854839463 | 0.084798902 | MIP-1a | 0.888145567 |
| 147 | HC - LC | D14 | 0.3424608 | 0.1864234 | 14 | 1.837004918 | 0.087527349 | MDC | 0.476537786 |
| 14 | HC - LC | D1 | 0.3899164 | 0.2195158 | 14 | 1.77625657 | 0.097419026 | FLT-3L | 0.888145567 |
| 143 | HC - LC | D14 | 0.4473417 | 0.2537927 | 14 | 1.762626221 | 0.099770331 | MCP-1 | 0.488874624 |
| 28 | HC - LC | D30 | 0.5368792 | 0.3051378 | 14 | 1.759464926 | 0.100322793 | GROa | 0.518071763 |
| 188 | HC - LC | D30 | 0.2989982 | 0.1728997 | 14 | 1.729315316 | 0.105728931 | TNFb | 0.518071763 |
| 56 | HC - LC | D30 | 0.6954172 | 0.4209944 | 14 | 1.651844455 | 0.120809398 | IL-17A | 0.534548436 |
| 106 | HC - LC | D1 | -0.3500560 | 0.2123557 | 14 | -1.648441758 | 0.121512671 | IL-4 | 0.888145567 |
| 179 | HC - LC | D14 | 0.5669754 | 0.3505814 | 14 | 1.617243224 | 0.128127881 | TGFa | 0.570751471 |
| 120 | HC - LC | D30 | 0.4790123 | 0.3038953 | 14 | 1.576241226 | 0.13729215 | IL-1a | 0.534548436 |
| 44 | HC - LC | D30 | -1.2526456 | 0.8203804 | 14 | -1.526908358 | 0.149056096 | IL-22 | 0.534548436 |
| 180 | HC - LC | D30 | 0.5879374 | 0.3887995 | 14 | 1.512186613 | 0.152728125 | TGFa | 0.534548436 |
| 114 | HC - LC | D1 | 0.7752508 | 0.5263700 | 14 | 1.472824734 | 0.16292249 | IL-2 | 0.888145567 |
| 4 | HC - LC | D30 | 0.8739250 | 0.5993140 | 14 | 1.458208996 | 0.166850195 | EGF | 0.545043969 |
| 15 | HC - LC | D14 | 0.3240855 | 0.2282250 | 14 | 1.42002615 | 0.177484655 | FLT-3L | 0.708134907 |
| 83 | HC - LC | D14 | -0.9334256 | 0.6741908 | 14 | -1.384512466 | 0.187872526 | IL-10 | 0.708134907 |
| 144 | HC - LC | D30 | 0.3697712 | 0.2773416 | 14 | 1.333270119 | 0.203732666 | MCP-1 | 0.620842514 |
| 190 | HC - LC | D1 | 0.3932459 | 0.3009401 | 14 | 1.306724986 | 0.212364767 | TRAIL | 0.888145567 |
| 98 | HC - LC | D1 | 0.6132932 | 0.4711454 | 14 | 1.301706878 | 0.214029105 | IL-6 | 0.888145567 |
| 176 | HC - LC | D30 | 0.9997461 | 0.7704496 | 14 | 1.297613848 | 0.215394342 | sCD40l | 0.620842514 |
| 95 | HC - LC | D14 | 0.2131412 | 0.1646425 | 14 | 1.294569288 | 0.216414368 | IL-7 | 0.753930278 |
| 170 | HC - LC | D1 | 0.3510347 | 0.2797104 | 14 | 1.254993507 | 0.230026863 | RANTES | 0.888145567 |
| 142 | HC - LC | D1 | 0.3090521 | 0.2466519 | 14 | 1.252989147 | 0.230733898 | MCP-1 | 0.888145567 |
| 103 | HC - LC | D14 | 0.3944797 | 0.3160220 | 14 | 1.248266553 | 0.232406562 | IL-5 | 0.753930278 |
| 122 | HC - LC | D1 | 0.4191603 | 0.3393456 | 14 | 1.23520165 | 0.237083679 | IL-1B | 0.888145567 |
| 36 | HC - LC | D30 | 0.4352736 | 0.3611403 | 14 | 1.205275841 | 0.248074643 | IFNy | 0.654604792 |
| 159 | HC - LC | D14 | -0.1872270 | 0.1566262 | 14 | -1.195374907 | 0.251796926 | MIP-1B | 0.753930278 |
| 110 | HC - LC | D1 | -1.1184478 | 0.9511131 | 14 | -1.175935671 | 0.259230662 | IL-3 | 0.888145567 |
| 166 | HC - LC | D1 | 0.1787450 | 0.1570776 | 14 | 1.137940822 | 0.274245329 | PDGF-AB/BB | 0.888145567 |
| 131 | HC - LC | D14 | -0.5311416 | 0.4688980 | 14 | -1.132744322 | 0.2763492 | IP-10 | 0.753930278 |
| 43 | HC - LC | D14 | -0.8560947 | 0.7567648 | 14 | -1.131256077 | 0.27695398 | IL-22 | 0.753930278 |
| 92 | HC - LC | D30 | 0.2542166 | 0.2264036 | 14 | 1.12284699 | 0.280390002 | IL-8 | 0.654604792 |
| 2 | HC - LC | D1 | 0.5795943 | 0.5244005 | 14 | 1.105251339 | 0.287683494 | EGF | 0.888145567 |
| 102 | HC - LC | D1 | 0.3295352 | 0.3021748 | 14 | 1.090544769 | 0.293887679 | IL-5 | 0.888145567 |
| 174 | HC - LC | D1 | 0.7501211 | 0.6878652 | 14 | 1.09050592 | 0.2939042 | sCD40l | 0.888145567 |
| 111 | HC - LC | D14 | -1.1019374 | 1.0118558 | 14 | -1.089026128 | 0.294533975 | IL-3 | 0.75958762 |
| 8 | HC - LC | D30 | 0.2641598 | 0.2430536 | 14 | 1.086837549 | 0.295467238 | Eotaxin | 0.654604792 |

| | | | | | | | | | |
|---|---|---|---|---|---|---|---|---|---|
| 40 | HC - LC | D30 | -0.1579130 | 0.1463383 | 14 | -1.07909579 | 0.298786145 | IL-27 | 0.654604792 |
| 20 | HC - LC | D30 | 0.1816243 | 0.1687948 | 14 | 1.076006578 | 0.300118179 | Fractalkine | 0.654604792 |
| 162 | HC - LC | D1 | 0.2600636 | 0.2434737 | 14 | 1.068138613 | 0.303530592 | PDGF-AA | 0.888145567 |
| 91 | HC - LC | D14 | 0.2141458 | 0.2034851 | 14 | 1.05239033 | 0.310446535 | IL-8 | 0.760594011 |
| 146 | HC - LC | D1 | 0.1827962 | 0.1789125 | 14 | 1.02170728 | 0.324251085 | MDC | 0.888145567 |
| 90 | HC - LC | D1 | 0.1998503 | 0.1964465 | 14 | 1.017326886 | 0.326257555 | IL-8 | 0.888145567 |
| 55 | HC - LC | D14 | 0.3927407 | 0.3864197 | 14 | 1.016357849 | 0.326702637 | IL-17A | 0.762306152 |
| 76 | HC - LC | D30 | 0.2232946 | 0.2233271 | 14 | 0.999854517 | 0.334349912 | IL-12 (p40) | 0.654604792 |
| 24 | HC - LC | D30 | -0.5374361 | 0.5554280 | 14 | -0.967607259 | 0.349659994 | G-CSF | 0.654604792 |
| 108 | HC - LC | D30 | 0.2425239 | 0.2514608 | 14 | 0.964459914 | 0.351180348 | IL-4 | 0.654604792 |
| 68 | HC - LC | D30 | 0.2047173 | 0.2126089 | 14 | 0.96288183 | 0.351944404 | IL-15 | 0.654604792 |
| 60 | HC - LC | D30 | 0.1447961 | 0.1532290 | 14 | 0.944965893 | 0.3607006 | IL-17E/IL-25 | 0.654604792 |
| 51 | HC - LC | D14 | 0.2361547 | 0.2526100 | 14 | 0.93485863 | 0.365706837 | IL-18 | 0.814528863 |
| 140 | HC - LC | D30 | 0.4738143 | 0.5250680 | 14 | 0.902386527 | 0.38211494 | MCP-3 | 0.668701144 |
| 74 | HC - LC | D1 | 0.1715224 | 0.1900905 | 14 | 0.902319361 | 0.382149391 | IL-12 (p40) | 0.911904869 |
| 59 | HC - LC | D14 | 0.1207355 | 0.1375463 | 14 | 0.877780683 | 0.394877341 | IL-17E/IL-25 | 0.827264317 |
| 42 | HC - LC | D1 | -0.6469538 | 0.7375818 | 14 | -0.877128161 | 0.395219643 | IL-22 | 0.911904869 |
| 139 | HC - LC | D14 | 0.4130378 | 0.4897499 | 14 | 0.843364755 | 0.413202487 | MCP-3 | 0.827264317 |
| 104 | HC - LC | D30 | 0.3037249 | 0.3603205 | 14 | 0.842929896 | 0.413437561 | IL-5 | 0.676358889 |
| 100 | HC - LC | D30 | 0.4587990 | 0.5450794 | 14 | 0.841710368 | 0.414097279 | IL-6 | 0.676358889 |
| 123 | HC - LC | D14 | 0.3085009 | 0.3730067 | 14 | 0.827065278 | 0.422073631 | IL-1B | 0.827264317 |
| 22 | HC - LC | D1 | -0.4139854 | 0.5033266 | 14 | -0.822498495 | 0.424581228 | G-CSF | 0.911904869 |
| 26 | HC - LC | D1 | 0.2060917 | 0.2627055 | 14 | 0.784497143 | 0.445820433 | GROa | 0.911904869 |
| 79 | HC - LC | D14 | 0.2600154 | 0.3375050 | 14 | 0.770404569 | 0.453865068 | IL-12 (p70) | 0.844528234 |
| 6 | HC - LC | D1 | 0.1612204 | 0.2147035 | 14 | 0.750898126 | 0.46514901 | Eotaxin | 0.911904869 |
| 138 | HC - LC | D1 | 0.3550335 | 0.4791811 | 14 | 0.7409171 | 0.47098919 | MCP-3 | 0.911904869 |
| 196 | HC - LC | D30 | 0.4123291 | 0.5576950 | 14 | 0.739345287 | 0.471912981 | VEGF-A | 0.74592697 |
| 167 | HC - LC | D14 | -0.1184629 | 0.1631430 | 14 | -0.726129496 | 0.479723936 | PDGF-AB/BB | 0.844528234 |
| 31 | HC - LC | D14 | 0.1764207 | 0.2445813 | 14 | 0.721317101 | 0.482587562 | IFNa2 | 0.844528234 |
| 50 | HC - LC | D1 | 0.1654777 | 0.2317188 | 14 | 0.714131499 | 0.486882509 | IL-18 | 0.911904869 |
| 84 | HC - LC | D30 | -0.5232153 | 0.7536623 | 14 | -0.694230403 | 0.498896663 | IL-10 | 0.750309437 |
| 12 | HC - LC | D30 | 0.1493774 | 0.2332141 | 14 | 0.640516398 | 0.532181348 | FGF-2 | 0.750309437 |
| 70 | HC - LC | D1 | -0.1028850 | 0.1648511 | 14 | -0.624108713 | 0.542592971 | IL-13 | 0.911904869 |
| 168 | HC - LC | D30 | 0.1139223 | 0.1827937 | 14 | 0.623228735 | 0.543154539 | PDGF-AB/BB | 0.750309437 |
| 52 | HC - LC | D30 | -0.1648452 | 0.2695650 | 14 | -0.611523074 | 0.550655093 | IL-18 | 0.750309437 |
| 3 | HC - LC | D14 | 0.3282193 | 0.5419435 | 14 | 0.60563368 | 0.554450102 | EGF | 0.936829483 |
| 124 | HC - LC | D30 | 0.2349269 | 0.3891315 | 14 | 0.603721247 | 0.555685485 | IL-1B | 0.750309437 |
| 158 | HC - LC | D1 | 0.0871756 | 0.1453423 | 14 | 0.599795467 | 0.558226106 | MIP-1B | 0.911904869 |
| 112 | HC - LC | D30 | 0.6546656 | 1.1153027 | 14 | 0.58698466 | 0.566560187 | IL-3 | 0.750309437 |
| 175 | HC - LC | D14 | 0.4054987 | 0.7070514 | 14 | 0.573506713 | 0.575399281 | sCD40l | 0.939818826 |
| 72 | HC - LC | D30 | -0.1018226 | 0.1851375 | 14 | -0.549983799 | 0.590997468 | IL-13 | 0.762075683 |
| 134 | HC - LC | D1 | -0.2820546 | 0.5143260 | 14 | -0.548396576 | 0.592057705 | M-CSF | 0.911904869 |
| 18 | HC - LC | D1 | 0.0791106 | 0.1495056 | 14 | 0.529147802 | 0.604992083 | Fractalkine | 0.911904869 |
| 118 | HC - LC | D1 | 0.1286360 | 0.2632577 | 14 | 0.488631434 | 0.632667852 | IL-1a | 0.911904869 |
| 78 | HC - LC | D1 | -0.1330279 | 0.2750452 | 14 | -0.483658242 | 0.636105802 | IL-12 (p70) | 0.911904869 |
| 194 | HC - LC | D1 | 0.2333350 | 0.4996097 | 14 | 0.467034645 | 0.647660596 | VEGF-A | 0.911904869 |
| 182 | HC - LC | D1 | 0.0618120 | 0.1343398 | 14 | 0.460116704 | 0.652497291 | TNFa | 0.911904869 |
| 130 | HC - LC | D1 | -0.2034724 | 0.4483781 | 14 | -0.45379643 | 0.656930358 | IP-10 | 0.911904869 |
| 19 | HC - LC | D14 | 0.0702410 | 0.1625048 | 14 | 0.432239347 | 0.672150865 | Fractalkine | 0.97164965 |
| 39 | HC - LC | D14 | -0.0545099 | 0.1310495 | 14 | -0.415949141 | 0.683752938 | IL-27 | 0.97164965 |
| 126 | HC - LC | D1 | 0.1070261 | 0.2663112 | 14 | 0.401883512 | 0.69383791 | IL-1RA | 0.911904869 |
| 82 | HC - LC | D1 | 0.2576900 | 0.6497016 | 14 | 0.39662814 | 0.697621613 | IL-10 | 0.911904869 |
| 96 | HC - LC | D30 | 0.0703675 | 0.1836712 | 14 | 0.383116735 | 0.707387586 | IL-7 | 0.871958272 |
| 94 | HC - LC | D1 | 0.0595576 | 0.1587875 | 14 | 0.375077132 | 0.713224174 | IL-7 | 0.911904869 |
| 186 | HC - LC | D1 | -0.0570844 | 0.1568124 | 14 | -0.364030167 | 0.721274479 | TNFb | 0.911904869 |
| 136 | HC - LC | D30 | 0.2116312 | 0.5910235 | 14 | 0.358075705 | 0.725628039 | M-CSF | 0.871958272 |

| | | | | | | | | | |
|---|---|---|---|---|---|---|---|---|---|
| 46 | HC - LC | D1 | 0.1861901 | 0.5295206 | 14 | 0.351620109 | 0.730359142 | IL-21 | 0.911904869 |
| 38 | HC - LC | D1 | 0.0420131 | 0.1263419 | 14 | 0.332534969 | 0.744412138 | IL-27 | 0.911904869 |
| 128 | HC - LC | D30 | -0.1027681 | 0.3111936 | 14 | -0.330238574 | 0.746109552 | IL-1RA | 0.871958272 |
| 71 | HC - LC | D14 | -0.0567183 | 0.1722402 | 14 | -0.329297472 | 0.746805577 | IL-13 | 0.97164965 |
| 160 | HC - LC | D30 | 0.0536742 | 0.1656830 | 14 | 0.323957016 | 0.750759627 | MIP-1B | 0.871958272 |
| 58 | HC - LC | D1 | -0.0389668 | 0.1327256 | 14 | -0.293588956 | 0.773379289 | IL-17E/IL-25 | 0.924282565 |
| 163 | HC - LC | D14 | -0.0737261 | 0.2521991 | 14 | -0.292333092 | 0.774319503 | PDGF-AA | 0.97164965 |
| 152 | HC - LC | D30 | 0.0646102 | 0.2241709 | 14 | 0.288218529 | 0.777402477 | MIG | 0.871958272 |
| 80 | HC - LC | D30 | -0.0856135 | 0.3049088 | 14 | -0.280783939 | 0.782982938 | IL-12 (p70) | 0.871958272 |
| 27 | HC - LC | D14 | -0.0663069 | 0.2727061 | 14 | -0.243144124 | 0.811420122 | GROa | 0.97164965 |
| 195 | HC - LC | D14 | 0.1150806 | 0.5130861 | 14 | 0.224290993 | 0.8257712 | VEGF-A | 0.97164965 |
| 119 | HC - LC | D14 | 0.0602631 | 0.2782053 | 14 | 0.21661369 | 0.831634087 | IL-1a | 0.97164965 |
| 62 | HC - LC | D1 | 0.0544644 | 0.2571271 | 14 | 0.211819168 | 0.835300804 | IL-17F | 0.943044032 |
| 107 | HC - LC | D14 | 0.0506065 | 0.2389777 | 14 | 0.211762241 | 0.835344364 | IL-4 | 0.97164965 |
| 99 | HC - LC | D14 | 0.1067978 | 0.5119074 | 14 | 0.208627211 | 0.837744135 | IL-6 | 0.97164965 |
| 23 | HC - LC | D14 | 0.0955400 | 0.4918979 | 14 | 0.19422741 | 0.848787923 | G-CSF | 0.97164965 |
| 156 | HC - LC | D30 | 0.0799314 | 0.4190361 | 14 | 0.190750714 | 0.851459384 | MIP-1a | 0.927144663 |
| 35 | HC - LC | D14 | -0.0654948 | 0.3446696 | 14 | -0.190021932 | 0.852019614 | IFNy | 0.97164965 |
| 150 | HC - LC | D1 | -0.0345023 | 0.1880150 | 14 | -0.183508253 | 0.857030467 | MIG | 0.943044032 |
| 29 | HC - LC | Baseline | -0.0383785 | 0.2230512 | 14 | -0.172061445 | 0.865851672 | IFNa2 | 1 |
| 115 | HC - LC | D14 | 0.0885658 | 0.5626364 | 14 | 0.157412218 | 0.877167705 | IL-2 | 0.97164965 |
| 135 | HC - LC | D14 | 0.0833833 | 0.5323257 | 14 | 0.156639697 | 0.87776525 | M-CSF | 0.97164965 |
| 32 | HC - LC | D30 | -0.0390243 | 0.2503267 | 14 | -0.155893532 | 0.878342481 | IFNa2 | 0.934263156 |
| 155 | HC - LC | D14 | -0.0560222 | 0.3809612 | 14 | -0.147054836 | 0.885185445 | MIP-1a | 0.97164965 |
| 127 | HC - LC | D14 | 0.0401261 | 0.2911109 | 14 | 0.137837985 | 0.892331311 | IL-1RA | 0.97164965 |
| 66 | HC - LC | D1 | -0.0245569 | 0.1827026 | 14 | -0.134409383 | 0.894992037 | IL-15 | 0.943044032 |
| 172 | HC - LC | D30 | -0.0428445 | 0.3222762 | 14 | -0.132943569 | 0.896129966 | RANTES | 0.934263156 |
| 121 | HC - LC | Baseline | -0.0454898 | 0.3461086 | 14 | -0.131432075 | 0.897303606 | IL-1B | 1 |
| 34 | HC - LC | D1 | -0.0380166 | 0.3222710 | 14 | -0.11796459 | 0.907771484 | IFNy | 0.943044032 |
| 30 | HC - LC | D1 | -0.0208144 | 0.2139531 | 14 | -0.097284758 | 0.923879292 | IFNa2 | 0.943044032 |
| 178 | HC - LC | D1 | -0.0270478 | 0.3388712 | 14 | -0.079817445 | 0.937512177 | TGFa | 0.943044032 |
| 86 | HC - LC | D1 | -0.0120390 | 0.1558684 | 14 | -0.077238356 | 0.939526935 | IL-9 | 0.943044032 |
| 10 | HC - LC | D1 | 0.0155657 | 0.2139990 | 14 | 0.072737436 | 0.943044032 | FGF-2 | 0.943044032 |
| 11 | HC - LC | D14 | -0.0101347 | 0.2216093 | 14 | -0.04573241 | 0.964169466 | FGF-2 | 0.974426063 |
| 67 | HC - LC | D14 | -0.0082459 | 0.1897562 | 14 | -0.043455191 | 0.965952393 | IL-15 | 0.974426063 |
| 183 | HC - LC | D14 | -0.0059852 | 0.1389101 | 14 | -0.043086905 | 0.966240756 | TNFa | 0.974426063 |
| 187 | HC - LC | D14 | 0.0051097 | 0.1565703 | 14 | 0.032635391 | 0.974426063 | TNFb | 0.974426063 |
| 184 | HC - LC | D30 | 0.0025157 | 0.1538406 | 14 | 0.016352369 | 0.987184035 | TNFa | 0.996070902 |
| 116 | HC - LC | D30 | -0.0029571 | 0.5898848 | 14 | -0.005013067 | 0.996070902 | IL-2 | 0.996070902 |
| 189 | HC - LC | Baseline | 0.0000000 | 0.2878773 | 14 | 2.56E-15 | 1 | TRAIL | 1 |
| 17 | HC - LC | Baseline | 0.0000000 | 0.1495056 | 14 | -8.13E-16 | 1 | Fractalkine | 1 |
| 21 | HC - LC | Baseline | 0.0000000 | 0.4722974 | 14 | -8.23E-16 | 1 | G-CSF | 1 |
| 37 | HC - LC | Baseline | 0.0000000 | 0.1263419 | 14 | 9.86E-16 | 1 | IL-27 | 1 |
| 57 | HC - LC | Baseline | 0.0000000 | 0.1327256 | 14 | 1.28E-15 | 1 | IL-17E/IL-25 | 1 |
| 101 | HC - LC | Baseline | 0.0000000 | 0.3021748 | 14 | -1.23E-15 | 1 | IL-5 | 1 |
| 105 | HC - LC | Baseline | 0.0000000 | 0.2123557 | 14 | -8.19E-16 | 1 | IL-4 | 1 |
| 113 | HC - LC | Baseline | 0.0000000 | 0.5062530 | 14 | 7.91E-16 | 1 | IL-2 | 1 |
| 125 | HC - LC | Baseline | 0.0000000 | 0.2663112 | 14 | 1.09E-15 | 1 | IL-1RA | 1 |
| 129 | HC - LC | Baseline | 0.0000000 | 0.4483781 | 14 | 1.74E-15 | 1 | IP-10 | 1 |
| 145 | HC - LC | Baseline | 0.0000000 | 0.1789125 | 14 | 1.32E-15 | 1 | MDC | 1 |
| 157 | HC - LC | Baseline | 0.0000000 | 0.1453423 | 14 | -8.50E-16 | 1 | MIP-1B | 1 |
| 161 | HC - LC | Baseline | 0.0000000 | 0.2434737 | 14 | -1.09E-15 | 1 | PDGF-AA | 1 |
| 165 | HC - LC | Baseline | 0.0000000 | 0.1570776 | 14 | 8.21E-16 | 1 | PDGF-AB/BB | 1 |
| 1 | HC - LC | Baseline | 0.0000000 | 0.5244005 | 14 | -1.16E-17 | 1 | EGF | 1 |
| 5 | HC - LC | Baseline | 0.0000000 | 0.2147035 | 14 | -4.02E-16 | 1 | Eotaxin | 1 |
| 9 | HC - LC | Baseline | 0.0000000 | 0.2078898 | 14 | 1.07E-16 | 1 | FGF-2 | 1 |

| | | | | | | | | | |
|---|---|---|---|---|---|---|---|---|---|
| 13 | HC - LC | Baseline | 0.0000000 | 0.2195158 | 14 | -1.87E-16 | 1 | FLT-3L | 1 |
| 25 | HC - LC | Baseline | 0.0000000 | 0.2627055 | 14 | 1.47E-16 | 1 | GROa | 1 |
| 33 | HC - LC | Baseline | 0.0000000 | 0.3099431 | 14 | 4.57E-17 | 1 | IFNy | 1 |
| 41 | HC - LC | Baseline | 0.0000000 | 0.7375818 | 14 | 1.70E-16 | 1 | IL-22 | 1 |
| 45 | HC - LC | Baseline | 0.0000000 | 0.5295206 | 14 | -6.70E-16 | 1 | IL-21 | 1 |
| 49 | HC - LC | Baseline | 0.0000000 | 0.2317188 | 14 | 5.88E-16 | 1 | IL-18 | 1 |
| 53 | HC - LC | Baseline | 0.0000000 | 0.3759576 | 14 | -2.74E-17 | 1 | IL-17A | 1 |
| 61 | HC - LC | Baseline | 0.0000000 | 0.2476510 | 14 | -5.28E-16 | 1 | IL-17F | 1 |
| 65 | HC - LC | Baseline | 0.0000000 | 0.1827026 | 14 | 4.73E-16 | 1 | IL-15 | 1 |
| 69 | HC - LC | Baseline | 0.0000000 | 0.1648511 | 14 | -1.12E-17 | 1 | IL-13 | 1 |
| 73 | HC - LC | Baseline | 0.0000000 | 0.1900905 | 14 | -8.10E-18 | 1 | IL-12 (p40) | 1 |
| 77 | HC - LC | Baseline | 0.0000000 | 0.2600538 | 14 | 2.92E-16 | 1 | IL-12 (p70) | 1 |
| 81 | HC - LC | Baseline | 0.0000000 | 0.6497016 | 14 | 4.47E-16 | 1 | IL-10 | 1 |
| 85 | HC - LC | Baseline | 0.0000000 | 0.1499552 | 14 | -5.00E-16 | 1 | IL-9 | 1 |
| 89 | HC - LC | Baseline | 0.0000000 | 0.1964465 | 14 | 2.72E-16 | 1 | IL-8 | 1 |
| 93 | HC - LC | Baseline | 0.0000000 | 0.1587875 | 14 | 2.17E-16 | 1 | IL-7 | 1 |
| 97 | HC - LC | Baseline | 0.0000000 | 0.4711454 | 14 | 3.83E-16 | 1 | IL-6 | 1 |
| 109 | HC - LC | Baseline | 0.0000000 | 0.9511131 | 14 | -4.44E-16 | 1 | IL-3 | 1 |
| 117 | HC - LC | Baseline | 0.0000000 | 0.2632577 | 14 | 1.57E-16 | 1 | IL-1a | 1 |
| 133 | HC - LC | Baseline | 0.0000000 | 0.5143260 | 14 | 2.47E-16 | 1 | M-CSF | 1 |
| 137 | HC - LC | Baseline | 0.0000000 | 0.4791811 | 14 | 5.76E-16 | 1 | MCP-3 | 1 |
| 141 | HC - LC | Baseline | 0.0000000 | 0.2466519 | 14 | 5.36E-16 | 1 | MCP-1 | 1 |
| 149 | HC - LC | Baseline | 0.0000000 | 0.1880150 | 14 | -1.71E-16 | 1 | MIG | 1 |
| 153 | HC - LC | Baseline | 0.0000000 | 0.3586521 | 14 | 4.69E-16 | 1 | MIP-1a | 1 |
| 169 | HC - LC | Baseline | 0.0000000 | 0.2797104 | 14 | 3.93E-16 | 1 | RANTES | 1 |
| 173 | HC - LC | Baseline | 0.0000000 | 0.6878652 | 14 | -5.34E-16 | 1 | sCD40l | 1 |
| 177 | HC - LC | Baseline | 0.0000000 | 0.3388712 | 14 | 4.09E-16 | 1 | TGFa | 1 |
| 181 | HC - LC | Baseline | 0.0000000 | 0.1343398 | 14 | -9.05E-17 | 1 | TNFa | 1 |
| 185 | HC - LC | Baseline | 0.0000000 | 0.1516535 | 14 | -2.58E-16 | 1 | TNFb | 1 |
| 193 | HC - LC | Baseline | 0.0000000 | 0.4996097 | 14 | -1.73E-16 | 1 | VEGF-A | 1 |

**Supplementary Table S18: Summary of Sample Availability by Clinical Responses**

| | STOOL SAMPLES | | | SERUM SAMPLES | | PBMC SAMPLES | |
|---|---|---|---|---|---|---|---|
| Timepoint | CRC-MSS (n samples) | Gastric (n samples) | Melanoma (n samples) | Melanoma Stable Disease (n samples) | Melanoma Progressive Disease (n samples) | Melanoma Stable Disease (n samples) | Melanoma Progressive Disease (n samples) |
| Baseline | 13 | 16 | 19 | 8 | 8 | 11 | 11 |
| D1 | 9 | 17 | 19 | 8 | 8 | 8 | 11 |
| D3 | 13 | 17 | 18 | na | na | na | na |
| D7 | 13 | 19 | 17 | na | na | na | na |
| D14 | 13 | 15 | 17 | 8 | 7 | 8 | 10 |
| D21 | 12 | 14 | 16 | na | na | na | na |
| D30 | 12 | 12 | 17 | 8 | 5 | 8 | 10 |
| D56 | 6 | 3 | 11 | 7 | 0 | 8 | 4 |
| D86 | 5 | 2 | 9 | 5 | 1 | 8 | 1 |
| D114 | 3 | 2 | 7 | na | na | na | na |
| D142 | 3 | 2 | 4 | na | na | na | na |
| D170 | 3 | 2 | 3 | na | na | na | na |
| EOT | 6 | 6 | 4 | na | na | na | na |

**Supplementary Table S19: Flow Cytometry Antibodies**

| Marker | Fluorescent Marker | Supplier | Catalogue | Lot |
|---|---|---|---|---|
| Viability | Zombie Near Infrared | Biolegend | 423106 | B388457 |
| CD11b | Brilliant Violet 570 | Biolegend | 301325 | B401013 |
| HLA-DR | cFluor® V505 | Cytek Biosciences | R7-20254 | F-062723-01 |
| CD14 | Spark Blue 550 | Biolegend | 367148 | B361330 |
| CD16 | PE-Fire 640 | Biolegend | 302068 | B356212 |
| CD19 | Alexa Fluor 488 | Biolegend | 302219 | B397267 |
| CD3 | APC-Fire 810 | Biolegend | 344858 | B363363 |
| CD33 | PE-Cy5 | Biolegend | 303406 | B402606 |
| CD56 | cFluor® BYG750 | Cytek Biosciences | R7-20283 | F-051823-01 |
| CD158 | eFluor450 | Thermo Fisher Scientific | 48-1589-41 | 2497973 |
| CD8 | Brilliant Violet 480 | BD Biosciences | 566121 | 3178581 |
| CD4 | Alexa Fluor 660 | Biolegend | 344676 | B377325 |
| TCRva7.2 | Brilliant Violet 711 | Biolegend | 351732 | B393151 |
| CD40 | PE-Fire 810 | Biolegend | 334355 | B403224 |
| CD274 (PD-L1) | Brilliant Violet 650 | BD Biosciences | 568321 | 3171430 |
| TIGIT | Brilliant Violet 605 | BD Biosciences | 747841 | 3261620 |
| CD278 (ICOS) | APC-Cy7 | Biolegend | 313529 | B397517 |
| CD134 (OX40) | PE-Cy5 | Biolegend | 350009 | B395925 |
| CD366 (TIM3) | Super Bright 780 | Thermo Fisher Scientific | 78-3109-42 | 2312466 |
| CD279 (PD-1) | PE-Dazzle594 | Biolegend | 329940 | B367981 |
| Eomes | PE | Thermo Fisher Scientific | 12-4877-42 | 2633379 |
| RORγT | APC | Thermo Fisher Scientific | 17-6988-82 | 2548779 |
| Gata3 | Alexa Fluor 700 | Thermo Fisher Scientific | 56-9966-42 | 2736763 |
| Foxp3 | PE-Cy7 | Thermo Fisher Scientific | 25-4776-42 | 2627903 |
| T-bet | Brilliant Violet 421 | Biolegend | 644816 | B363145 |
| TCF1/TCF7 | Alexa Fluor 647 | Biolegend | 655203 | B364130 |
| CD94 | Brilliant Violet 510 | BD Biosciences | 743949 | 3261622 |
| CD107a | APC-Cy7 | Biolegend | 328630 | B362202 |
| IL-10 | PE | Biolegend | 506804 | B357934 |
| IL-17A | Brilliant Violet 605 | Biolegend | 512326 | B376461 |
| IL-4 | PE-Cy7 | Thermo Fisher Scientific | 25-7049-82 | 2630364 |
| IFN-γ | Brilliant Violet 570 | Biolegend | 502534 | B370493 |
| TNF-α | APC | Biolegend | 502912 | B351608 |
| TL1A | PerCP-eFluor710 | Thermo Fisher Scientific | 46-7911-82 | 2686541 |
